# A systematic review of the noma evidence landscape: current knowledge and gaps

**DOI:** 10.1101/2025.02.07.24315593

**Authors:** Brittany J. Maguire, Rujan Shrestha, Prabin Dahal, Roland Ngu, Lionel Nizigama, Sumayyah Rashan, Poojan Shrestha, Elinor Harriss, Paul N. Newton, Yuka Makino, Benoit Varenne, Philippe J. Guérin

**Author notes:** **Corresponding authors:** Brittany J. Maguire and Philippe J. Guérin.

## Abstract

**Background:** Noma (cancrum oris) is a severe gangrenous disease of the mouth and oro-facial structures. Noma often presents in young children alongside extreme poverty, malnutrition and poor environmental sanitation. Gaps however remain in understanding its aetiology, pathogenesis, preventative and treatment efficacy.

**Methods and findings:** We systematically searched databases to find all primary research studies reporting patients of any age diagnosed with noma (including clinical trials, cohort studies, case-control, cross-sectional, other observational studies, case studies and case series) up to 7 December 2022. The 366 publications included in our review describe 15,082 noma patients, from manuscripts published between 1839 and 2022. While several cohort (n=53) and cross-sectional (n=29) studies were identified, accounting for a total of 13,489 enrolled noma patients, only 6 interventional studies enrolling a total of 101 patients (range 7 to 26 patients) were identified, with only one interventional study from the last decade. Over 380 different treatment modalities were described for noma management. Disease aetiology reports identified 117 different microorganisms across 113 publications, but none were more widespread or uniquely related to noma development. Since 2000, most (91.2%) cases have been reported in Sub-Saharan Africa, but not solely in the historical ‘noma belt’. Noma was also observed in Asia and the Americas. There were 212 different possible noma risk factors presented in 269 (73.5%) publications, with substantial heterogeneity. The definition and harmonisation of noma progression staging are poorly standardised and reported.

**Conclusion:** The current literature provides very weak evidence to guide policy. Our thorough review also identified substantial knowledge gaps, and highlights the lack of prospective high-quality studies on the physiopathology of the disease that can guide therapeutic and preventive policies. Urgent research investment is therefore essential to improve the situation, especially as noma is now duly recognised as a Neglected Tropical Disease (NTD) by the World Health Organization (WHO).

**Research in context:** *What is already known on this topic:* No exhaustive synthesis of the noma evidence landscape existed at the inception of this review in 2018, particularly any relating to evidence-based risk factors, microbiology, prevention, treatment, or the burden of disease. Since commencing this work, high-quality systematic and scoping reviews on noma have been published, using existing data collected from clinical trials, longitudinal patient observational studies and retrospective studies as their sources. This systematic review was designed to encompass a wider evidence landscape, including case series and case reports as well. We hypothesised that in the context of scarce noma data, case series and reports are an underutilised source of information that could potentially help address research priorities and bridge knowledge gaps. We conducted comprehensive literature searches from database inception to 7 December 2022 across 11 different global and regional databases. Searches were not restricted on language of publication. Both English and French search strings were developed. English search terms included: “noma OR cancrum oris OR necrotising ulcerative stomatitis OR necrotising stomatitis OR Gangrenous stomatitis”. Noma publications on patients of any age and all primary research designs were included. Animal studies and studies on people with noma-like illnesses were excluded.

*What this study adds:* This review includes a wider range of published material, encompassing case reports and case studies not included in previous reviews, therefore providing an exhaustive synthesis of all reported information. This work creates a comprehensive baseline knowledge of any reported risk factors, microbiology, and treatment modalities for noma from which to innovatively approach research and address gaps. From synthesis across a broad scope of study designs, we provide recommendations to guide methodology and reporting for future noma research.

*How this study might affect research, practice or policy:* The current research evidence base has a considerable gap concerning the multi-factorial etiopathogenesis of noma. The paucity and methodological heterogeneity of high-quality studies in this area means that caution is needed in interpreting this evidence when designing future evidence-guided strategies and interventions to tackle noma. By including a broad range of study designs, this work provides valuable insights and a rich and exhaustive reference point from which to approach the design of future strategies and new robust studies that are desperately needed to address the many outstanding knowledge gaps of noma.

## INTRODUCTION

Noma is a rapidly progressive and debilitating gangrenous disease that involves the mouth and orofacial structures. At present, substantial research gaps remain regarding the aetiology, pathogenesis, and treatment of noma. Often reported as a polymicrobial infection, noma is commonly observed with multiple concomitant risk factors, including malnutrition, immunosuppression, infectious comorbidities, poor oral hygiene and extreme poverty.^1-3^ The evidence, causality and impact of risk factors on the development and progression of noma remain unclear.

The “noma belt”— an area of Sub-Saharan Africa between Mauritania and Ethiopia—is often considered to be the main location of the disease. However, more recent reports also continue to identify noma cases in other parts of the world.^3-5^ High-quality epidemiological data on noma are scarce, with few and varying reports (often citing outdated estimates) on the incidence and prevalence of noma.^4^ Accordingly, the current regional and global burden of noma is unclear.^6^

Noma disproportionately affects the world’s poorest and most remote communities and is common in children younger than six years.^7^ Left untreated, the case-fatality rate is high, and survivors are often left with severe disfigurement and varying degrees of functional impairment for life, including difficulty in eating and speaking that can have devastating physical, social and psychological effects. However, early diagnosis and treatment with antibiotics, nutritional support, and surgical management (e.g., wound care, debridement of necrotic areas and sequestrated bone) in early and intermediary stages of noma can reduce case-fatality.^8^ Yet, detection is difficult because of the disease’s elusive pathogenesis, a diagnosis based solely on clinical characteristics, a lack of disease knowledge, social stigma, and, in some areas, inadequate knowledge in healthcare professionals about how to identify the condition.^9-11^

As of December 2023, noma was duly recognised and included in the official list of Neglected Tropical Diseases (NTDs) from the World Health Organization (WHO).^12^ Acknowledging noma as an NTD is a pivotal step in increasing the attention and resourcing required to eradicate this social and biomedical indicator of extreme poverty and malnutrition affecting the most vulnerable populations.

This study aimed to exhaustively review and synthesise the existing literature to generate an account of the present state of knowledge about various aspects of noma.

## METHODS

The protocol was prospectively designed and registered in accordance with PRISMA guidelines (PROSPERO 2019 CRD42019124839). The methodology is briefly described below: there were no protocol deviations from the detailed methodological approach published separately.^13^

Literature searches were initially conducted from databases inception to 4 March 2019, and updated on 7 December 2022, using the search strategy and information sources detailed below. The following databases were searched: MEDLINE, CAB Abstracts, Embase, Global Health (all via Ovid), Scopus, Web of Science (Core Collection), African Index Medicus; Pascal (French language search); ClinicalTrials.gov, and WHO ICTRP. African Journals Online: Health and OpenGrey.eu were searched in 2019. WHO-identified and grey literature was also searched. Supplementary File 1 (SF1) provides the comprehensive strategy, study selection criteria and search string by database. Noma publications on patients of any age and all primary research designs were included. Animal studies and studies on people with noma-like illnesses were excluded.

Supplementary File 2 (SF2) contains the pre-piloted data collection and variable dictionary with all data items extracted, definitions and examples. Patients were classed according to the 2016 WHO Noma Disease phases, where sufficient details were supplied.^14^ To aid evidence synthesis, categories for both treatment and risk factors were constructed for data extraction, due to the heterogeneity of reporting and variability in terminology and classifications (See SF2 for all definitions).

A narrative (descriptive) synthesis was performed owing to the heterogeneous data yielded. R (version 3.6.3, R Foundation, Vienna, Austria) was used for aggregate quantitative synthesis and data visualisation. The Oxford Centre for Evidence-Based Medicine Levels of Evidence was used to assess the level of evidence and quality of individual studies across this systematic review.^15^

This systematic review did not use any personal information and did not require ethics approval.

## RESULTS

### Study characteristics

From 11,843 records identified across all database and manual searches until December 2022, 4,808 unique publications were screened, of which 366 were eligible and included for analysis (PRISMA flow diagram, search results; see SF1&SF2). The 366 publications describe 15,082 noma patients from 69 countries published between 1839 and 2022 (Figure 1). More articles reporting noma cases were published since 2001 than in the 50 years from 1951 to 2000. The spatial distribution of studies for various time periods can be found in Supplementary File 3 (SF3_Section1a).

**Figure 1.**
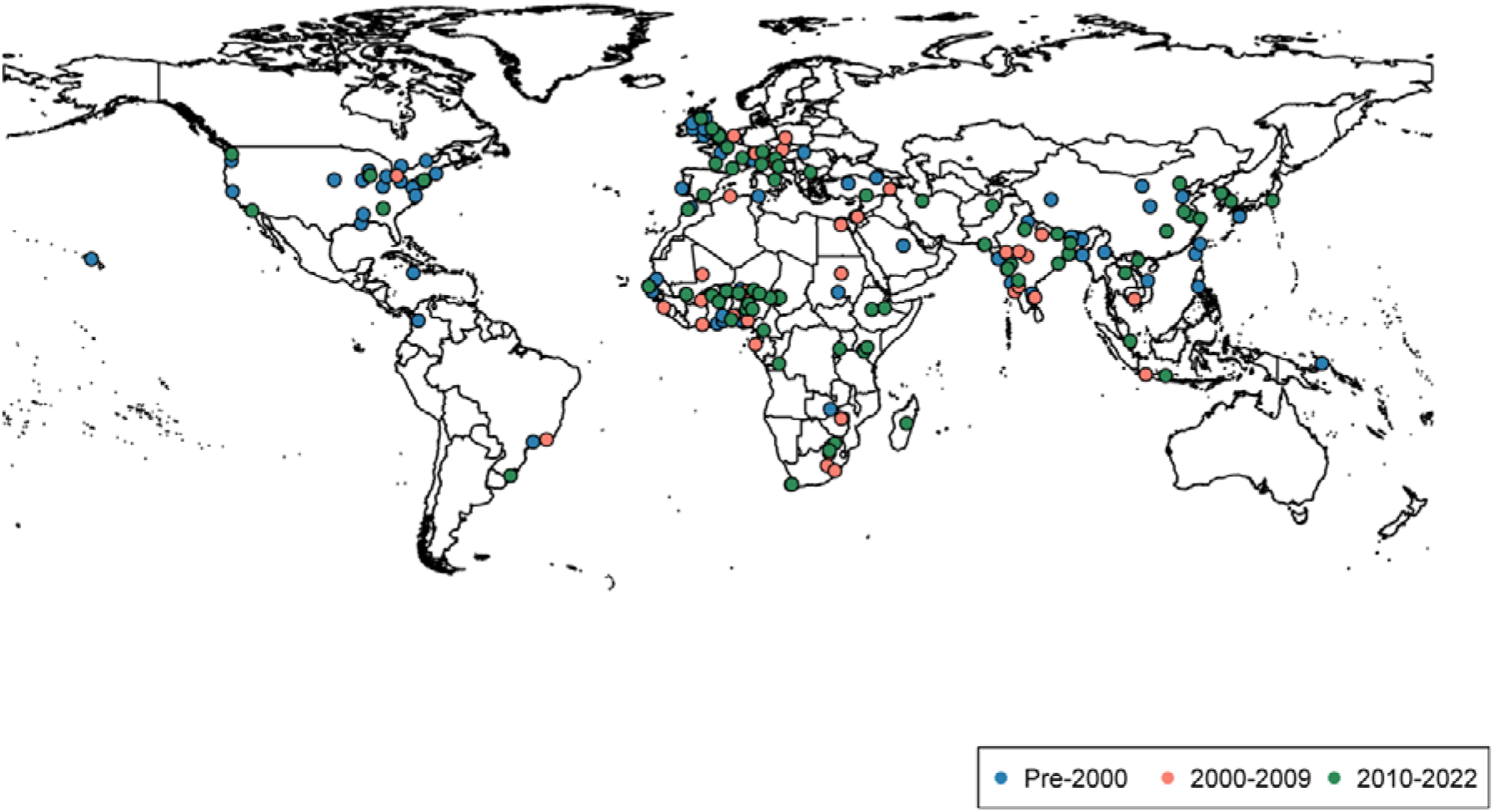
Spatial distribution of all included noma studies. Legend: The dots represent the study sites and are not weighted for the number of studies or the number of patients reported.

Of the 366 studies, 86.6% were single-centre studies (n=317), 9.6% multicentre studies (n=35), and unstated for 14 (3.8%). Study designs for the majority of publications were case series/reports (n=270). Only 6 interventional studies were identified: these included 101 patients, and the number of patients in individual studies ranged from 7 to 26 (Table 2). The median number of patients [range] included was 1 for case report/case series [1-69], 40 for cohort studies [2-212], 57 for cross-sectional studies [2-7195], 16 for interventional studies [7-26], and 65 for other study designs [8-296] (Figure 2 and SF3_Section1d).

**Figure 2.**
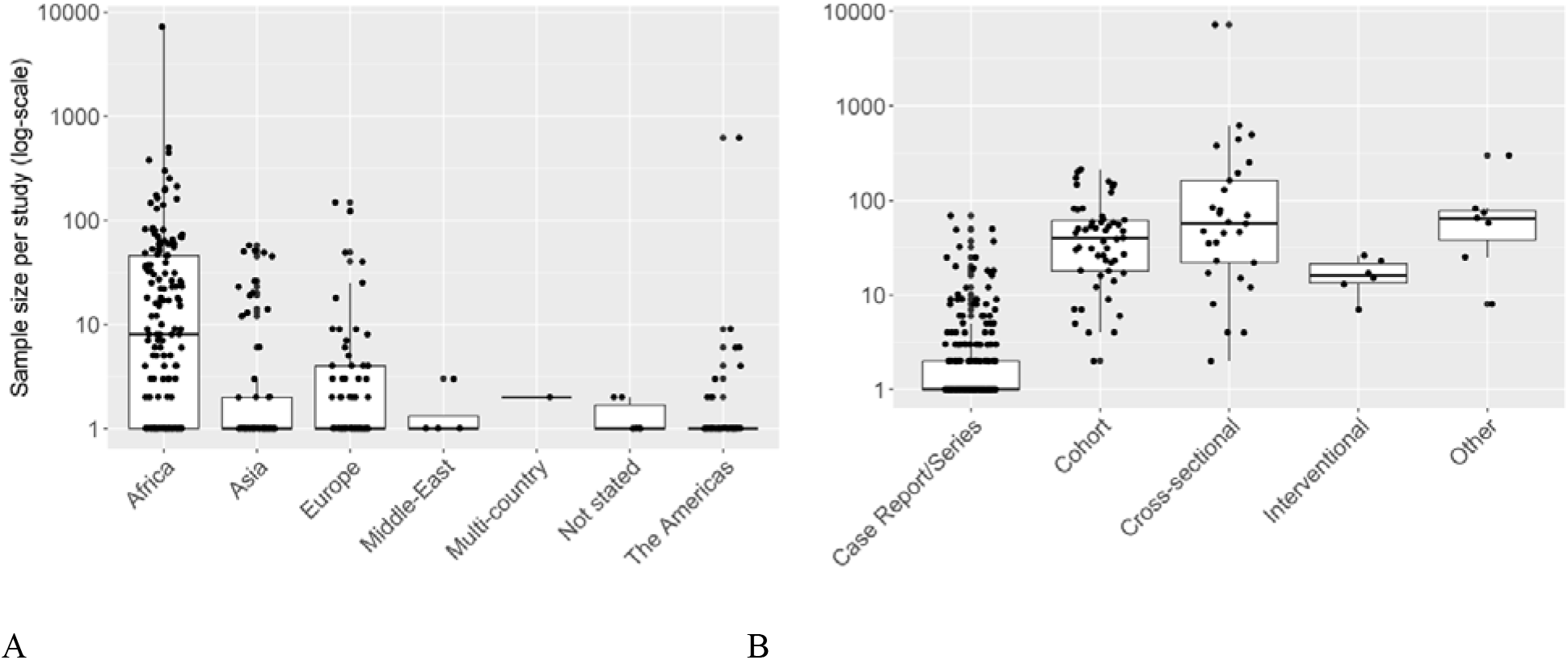
Number of included noma patients (logarithmic scale) per study by A) Region; B) Study design

Across all study designs, most publications included patients at different disease stages or reported on patients who progressed through more than one noma stage (Table 1, SF3_Table5). This category of multiple stages was assigned to 127 (35%) studies accounting for 10,981 patients. A single noma disease stage could only be assigned to 54% of studies, with the scarring stage underrepresented (0.8%).

**Table 1.**
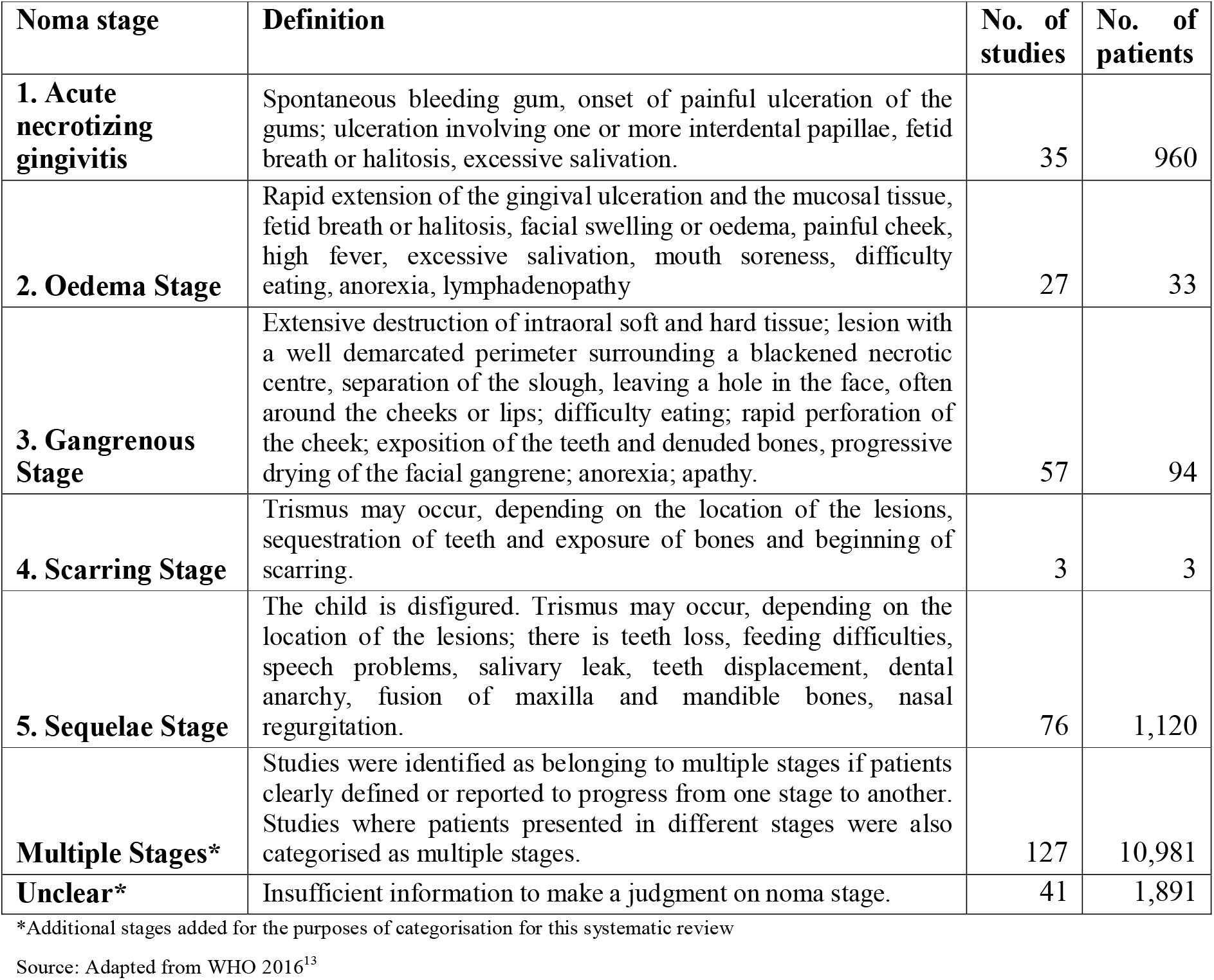
Noma studies by disease stage.

| Noma stage | Definition | No. of studies | No. of patients |
| --- | --- | --- | --- |
| **1. Acute necrotizing gingivitis** | Spontaneous bleeding gum, onset of painful ulceration of the gums; ulceration involving one or more interdental papillae, fetid breath or halitosis, excessive salivation. | 35 | 960 |
| **2. Oedema Stage** | Rapid extension of the gingival ulceration and the mucosal tissue, fetid breath or halitosis, facial swelling or oedema, painful cheek, high fever, excessive salivation, mouth soreness, difficulty eating, anorexia, lymphadenopathy | 27 | 33 |
| **3. Gangrenous Stage** | Extensive destruction of intraoral soft and hard tissue; lesion with a well demarcated perimeter surrounding a blackened necrotic centre, separation of the slough, leaving a hole in the face, often around the cheeks or lips; difficulty eating; rapid perforation of the cheek; exposition of the teeth and denuded bones, progressive drying of the facial gangrene; anorexia; apathy. | 57 | 94 |
| **4. Scarring Stage** | Trismus may occur, depending on the location of the lesions, sequestration of teeth and exposure of bones and beginning of scarring. | 3 | 3 |
| **5. Sequelae Stage** | The child is disfigured. Trismus may occur, depending on the location of the lesions; there is teeth loss, feeding difficulties, speech problems, salivary leak, teeth displacement, dental anarchy, fusion of maxilla and mandible bones, nasal regurgitation. | 76 | 1,120 |
| **Multiple Stages\*** | Studies were identified as belonging to multiple stages if patients clearly defined or reported to progress from one stage to another. Studies where patients presented in different stages were also categorised as multiple stages. | 127 | 10,981 |
| **Unclear\*** | Insufficient information to make a judgment on noma stage. | 41 | 1,891 |

\*Additional stages added for the purposes of categorisation for this systematic review
Source: Adapted from WHO 2016<sup>13</sup>

### Risk factors

Risk factors were reported in 269 (73.5%) publications (SF3_Table7). Malnutrition and concomitant co-infections appear to be the most frequently reported risk factors (SF3_Figure11), with no substantial variations in risk factors by geographical region (Figure 3). Specifically, 142 (38.8%) studies identified nutritional deficit as a risk factor, 36 (9.8%) did not, and 188 (51.4%) did not report nutritional status. Immunosuppression from non-infectious comorbidities was reported in 22.7% of risk factor studies (61/269), including cancer in 25, 12 being leukaemia. Infectious disease (ID) comorbidities were reported in 56.5% (152/269) of publications: measles constituted the main reported ID risk factor (35.5%, 54/152) across all study periods (18 before 1950, 18 between 1951-2000, and 17 from 2000 onwards). Other ID risk factors included malaria, HIV, visceral leishmaniasis, syphilis, dysentery, viral infections (CMV, EBV), enteric fevers, meningitis, plague, gonorrhoea, herpes zoster, chlamydia, tuberculosis, and poliomyelitis. Unsurprisingly, HIV as a risk factor was most reported after 2000 (32/154 publications during this period) (see SF3_Section2b). In 53 papers, unidentified pneumopathies were the most reported respiratory infections (30%). Poor oral hygiene was reported in 28% of publications (75/269). Only 39 publications detailed physical environment factors (e.g., close proximity to livestock, scant sanitation and unsafe drinking water) in 1,841 patients (1,797 from Africa). Lifestyle factors, including smoking, alcoholism, and intravenous drug use, were captured in 41 publications (737 patients), with 51% of these referring to adult patients (21/41). In publications post-2000, nutritional deficit, immunosuppression and infectious diseases remained the leading risk factor categories, accounting for 59.1% (75/127), 29.9% (38/127) and 48.8% (62/127), respectively.

**Figure 3.**
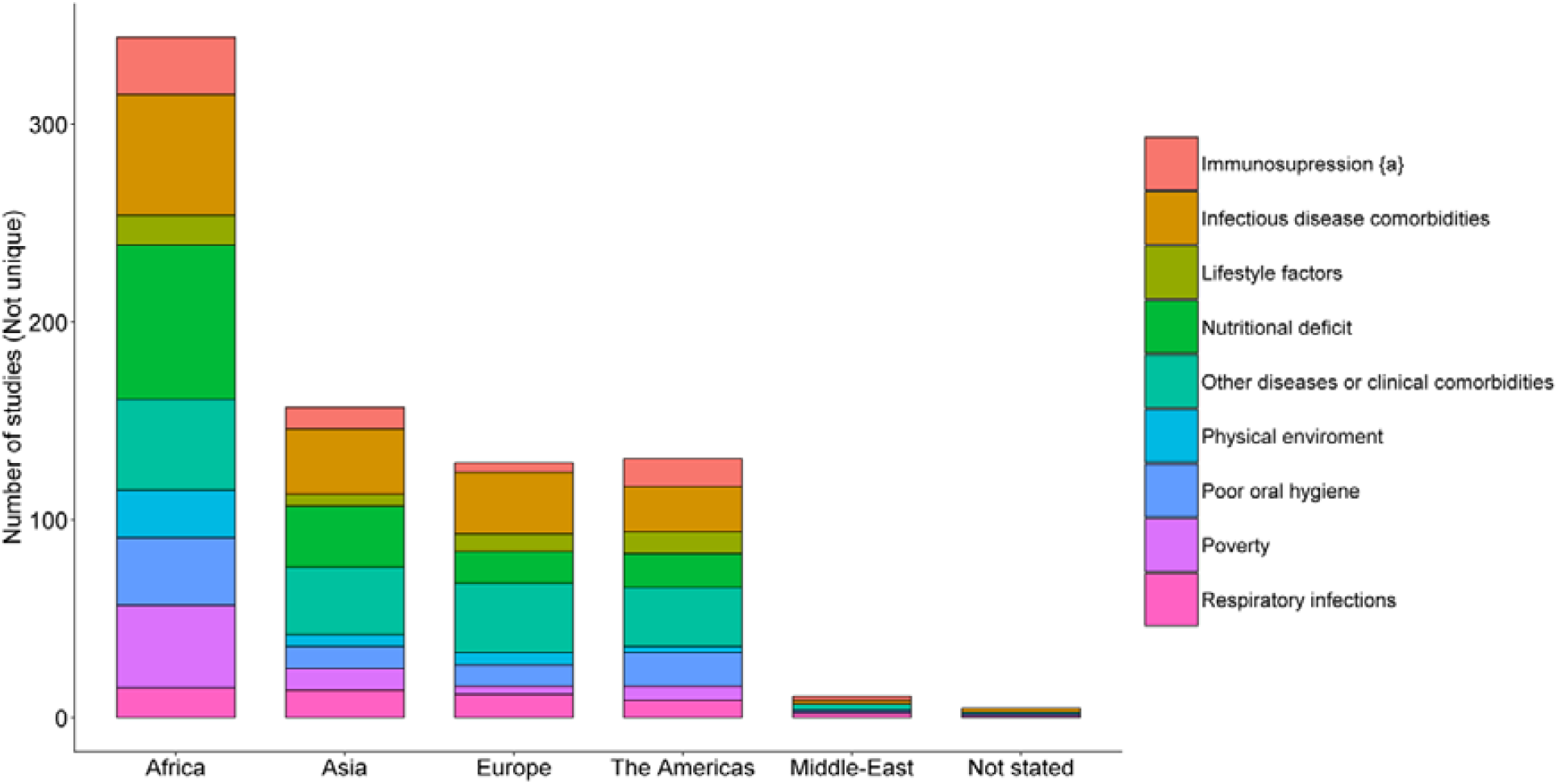
Number of publications reporting risk factor categories by geographical region. Same study might report multiple risk factors and hence the number of studies may sum to >100%. ^a^non-infectious comorbidities causing immunosuppression.

### Microbiology

There were 113 (31%) publications reporting microbiological agents or testing. Of these, 78 (69%) identified multiple microorganisms, 20 (18%) a single microorganism, and 15 (13%) did not report any specific organism. Information on culture techniques was seldomly reported. Substantial heterogeneity in the variety and level of classification of identified microorganisms (genus vs species) impacted the feasibility of evidence synthesis. A total of 117 different microorganisms (at the family/genus/or species level) were described across these 113 publications. The most frequently documented microorganisms, as per exact taxonomic level reported, included: *Staphylococcus aureus* (22/113, 19%), *Pseudomonas aeruginosa* (20/113, 18%), fusiform bacilli (18/113, 16%), spirochetes (14/113, 12%), *Fusobacterium necrophorum* (13/113, 11%), *Prevotella* genus (13/113, 11%), *Klebsiella pneumoniae* (11/113, 10%), *Peptostreptococcus* genus (8/113, 7%) and *Actinomyces* species (6/113, 5%). One publication could report multiple microbes, so percentages add up to more than 100. More recent case-studies reported cultures of resistant organisms.^16-18^ See SF3_Section3 for a comprehensive list of microorganisms as reported and differences observed by region.

### Treatment modalities

Most studies (96%, 352/366, n=4,207 patients) reported treatment details for noma cases or study population. 30% of studies described medical interventions (108/366, n=654), 26% surgical interventions (94/366, n=1,554), and 33% of publications reported both medical and surgical interventions (119/366, n=1,999). A further 8% reported neither medical nor surgical interventions (31/366, n=2,665). The intervention was unknown for a large number of patients (n=8,210), though from a limited number of studies in 4% (14/366).

Across 352 studies, 381 different therapeutic interventions were described (SF3_Table10). **Error! Reference source not found**. depicts the total publications and patients for each of the 5 sub-group intervention categories. While providing an overview of the reported literature, these results may not align with current intervention proportions. Notably, the majority of included patients received alternative therapies, a result influenced by a single retrospective cross-sectional study of over 7,000 patients.^19^ In this study, the most common intervention was home treatment with charcoal in 42% of patients (n=3,011), followed by 13% (n=951) opting for traditional treatment, including cultural rituals. Antibiotics were prescribed in 176 studies, with no evidence to support better results from any antibiotic family (SF3_Table15 and Figure13).

For each study design, the number of patients and studies by intervention category are presented in Table 2. Further discussion of noma treatment by study design, noma stage and year of publication, including numbers by additional sub-group intervention categories and detailed observations are provided in SF3_Section5. This review found little evidence on noma prevention (SF3_Section4).

**Table 2.**
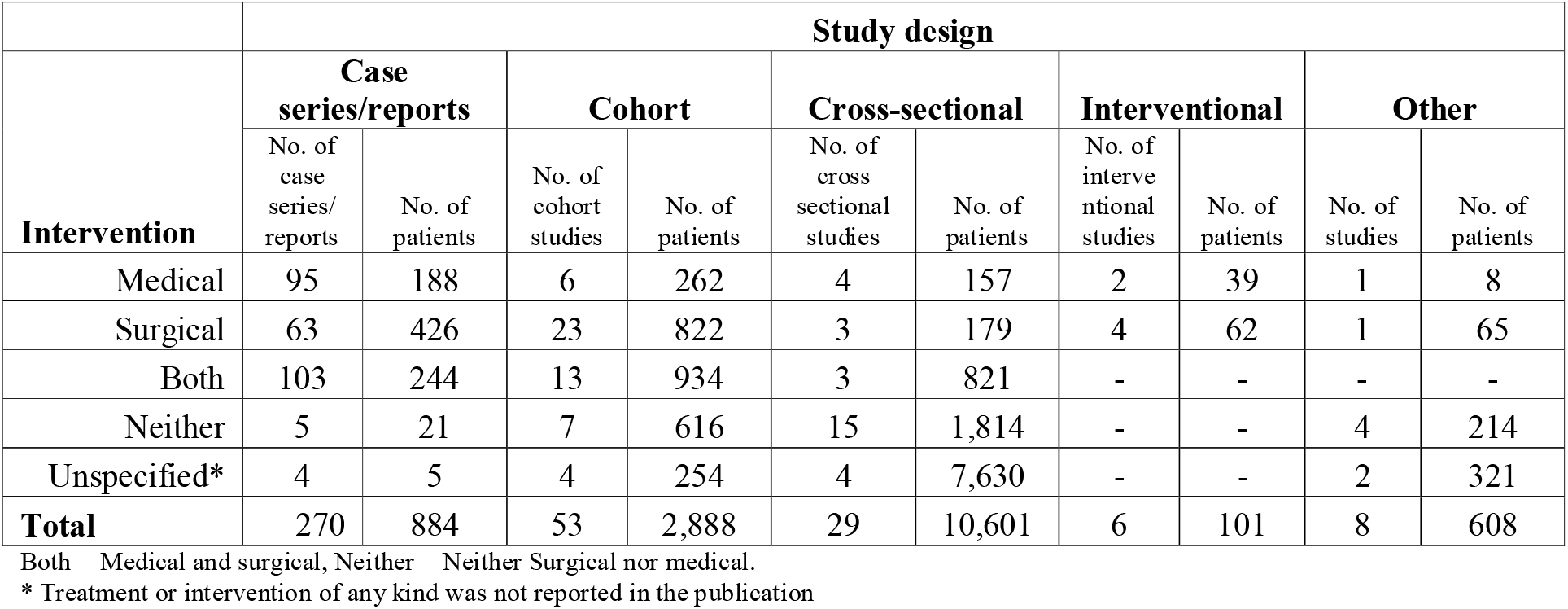
Number of studies and patients by intervention and study design.

### Mortality

Mortality data was reported in 77% of publications (283/366). This included 187 studies explicitly reporting no noma patient fatalities. 291 deaths were reported in 283 publications, with 4,181 noma participants. Out of those 291 patients, 88 died from noma’s acute effects shortly after admission to a health facility. The remaining 203 stated cause of death was not noma-related. SF3_Section6 provides the distribution of death by age, stage, and publication period. The median [IQR] case-fatality rate observed from the 37 cohort or interventional studies was 0% [IQR: 0-9.7%, range: 0-83.3%] (SF3_Figure21). Of the 12 studies that did not use any surgery (facial reconstruction or other forms), the median case-fatality rate was 4.6% [IQR: 0-28.4%; range: 0-83.3%]. Using a formal meta-analysis of these 12 studies, the estimate of case-fatality was 9.9% [95% CI: 7.9%-12.2%; *I*^*2*^=87.3%] using a common-effect meta-analysis, and 5.5% [95% CI:0.9%-26.7%] from random-effects meta-analysis. The estimated 95% prediction interval was wide (0-94.2%), indicating large heterogeneity of the estimated effect (SF3_Table23).

### Post-noma complications

Post-noma complications were reported in 205/366 (56.0%) articles, with 142/205 (62.3%) of these publications reporting complications related to the facial region, including trismus, sequestration of the jaws, bony ankylosis of temporomandibular joints, severe scarring, fistulisation of cheeks, and oro-nasal fistula (SF3_Section7).

### Burden of disease

Of 366 studies, 118 (32.2%) reported indicators related to the burden of disease: incidence, prevalence, quality-adjusted life years (QALYs), disability-adjusted life years (DALYs), financial costs, and morbidity or mortality rates. Of 118 publications, only 17 reported primary source estimates (i.e., rates or costs calculated directly from primary research reported in the publication). Secondary sources (i.e., rates reported as sourced from other literature and not calculated from the primary data) were cited to report the burden of disease in 91 studies. Ten publications cited both primary and secondary source estimates.

Since 2000, studies citing a secondary source of the burden of noma most commonly used WHO’s 1998 estimate of global incidence: this was reported in 11 publications, estimating an incidence of 140,000 new cases/year.^20^ The latest global estimate was published in 2003. Fieger *et al*. reported a lower global incidence of 30,000-40,000 cases/year. Of these, 25,600 cases were expected to originate from Sub-Saharan Africa.^21^ Note that this figure was calculated using observed noma patients in north-west Nigeria, then extrapolating to a national incidence before being extrapolated to global incidence. The global prevalence of noma survivors was estimated to be 210,000 worldwide, based on a 15% untreated survival rate and a life expectancy of 40 years following the disease.^9,22^ See SF3_Section8 for further results.

### Risk of Bias

Of the 366 included studies, 1 (0.3%) study was graded at Level 2 risk of bias, 24 (6.6%) studies were Level 3, and the remaining 341 (93.2%) studies were at Level 4 risk of bias. No studies were graded to be at Level 1 (lowest) or Level 5 (highest) studies risk of bias (SF3_Table33).

## DISCUSSION

Box 1 summarises the key findings and recommendations of this systematic review.

The 366 publications eligible for inclusion represented 15,082 noma cases from 69 countries, published between 1839 and 2022. Research publications on noma increased significantly in the last two decades, in line with the 1994 WHO initiative to launch an international programme to control noma, which increasingly raised the disease profile as a public health issue. However, nearly three-quarters of studies identified were case-reports or case-series, representing a very low level of evidence for policy guidance. Only 6 interventional studies enrolling 101 patients (range 7 to 26 patients) were identified, with only one study from the last decade.

Since 2000, the majority of noma cases have been reported in Africa, but not solely in the historical ‘noma belt’: cases are also reported in South and South-East Asia, Asia-Pacific and the Middle East. Recent cases reported in Europe and the USA have either been patients imported for surgical reconstruction or, mostly been linked to vulnerable minorities, lower levels of education and oral hygiene practices, cancer and HIV-coinfection.^23,24^ Importantly, these countries reflect the country in which the included studies were conducted.

### Risk Factors

The literature reported 212 different risk factors associated with noma, with infectious diseases detailed in half of them, and measles the most frequently named disease. This corroborates the hypothesis that measles may be an important contributory factor for noma.^25,26^ While many noma-affected children are reportedly HIV-seronegative,^27,28^ recent studies have also focused on factors associated with HIV infection which are involved in the development of noma precursors.^29-31^ Existing health infrastructure, disease prevention, and management programs for these associated infectious diseases would be paramount in future strategies for effective awareness, early case identification, management, and referral for noma.

Chronic malnutrition has commonly been reported in communities affected by noma, and may increase susceptibility to infection.^27,32^ In over a third of publications, nutritional deficit was described as a risk factor for noma, including wasting, stunting, kwashiorkor, marasmus, or a low-protein diet. Reduced immune fitness and disturbed metabolism resulting from present or past malnutrition could play a vital role in both the pathogenesis and progression of noma.^33^ The evidence synthesised supports common knowledge that noma manifests itself in settings of severe poverty.^3,34^ However, the causality and interdependency of many poverty-associated risk factors could not be accurately teased out, given the level of reporting.

### Microbiology

A conservative reading of the results of this review points towards the causative agents of noma being non-specific polymicrobial organisms. Recent publications support this assumption.^35-36^ While fusobacteria and spirochetes are historically reported to be the causative agent of noma, this review aligns with more recent findings that report no clear associations in microbiology analyses, nor noma replication in animal models inoculated with these species.^1,26,35-37^ From the 117 microorganisms reported, none appear to be more widespread or uniquely related to the development of noma. The diagnosis of noma, therefore, still remains a clinical one. Without further research, current evidence does not support the development of a definitive technique to diagnose acute noma. The scarcity of information on the aetiopathogenesis of noma, therefore, demands urgent basic science research on the microbiology, pathophysiology, pathobiology, and histopathologic features of noma.^1^

### Treatment modalities and outcomes

With over 381 different descriptions of noma management reported across 352 publications, the level of evidence for noma treatment was restricted to narrative synthesis. Interventions observed in included studies differed considerably across time periods, study design and noma stage.

Unsurprisingly, antibiotics (most commonly penicillin, metronidazole, clindamycin, streptomycin and ceftriaxone) were the predominant treatment for noma after 1950 once they were widely available.^38-40^ Although 176 publications (across all time-periods) reported the use of antibiotics (accounting for 2,254 patients), the heterogeneity of specific antibiotic(s) used, dosing regimens, treatment assessment and quality of study design prevented further comparisons to support one antibiotic or regimen over another. The current levels of evidence to guide one specific antibiotic treatment regimen over another for noma are, therefore, very weak.

However, the evidence in this review does support expert observation that antibiotics in the acute stages of noma reduce case fatality.^11,29^ Due to the polymicrobial nature of noma concomitant infection, broad-spectrum antibiotics should be considered. Currently, the WHO proposes a combination of antibiotics for 14 days, although this is a short antibiotic regimen if the infection is already affecting the bones, where antibiotics should be given for 4 to 6 weeks.^14,41^

Surgical reconstruction was assessed in 4 of the 6 interventional studies, with comparable results demonstrating favourable patient and clinical outcomes, with no serious complications and no deaths.^42,45^ Similarly, the most common interventions observed in cohort studies identified in this review was surgical reconstruction of the face in noma survivors, mainly through the use of flaps.^23,46^

### Mortality and post-noma complications

While the mortality rate for untreated patients is reported to be as high as 70-90%,^11,20^ with appropriate treatment, the mortality rate of noma reported in the literature is usually less than 10%.^2,48^ Considering the totality of published evidence reviewed, our results support this estimate. Mortality rates are likely to be underestimated, but these rates still make noma one of the deadliest diseases in young populations.

However, serious caution is required in interpreting these results. Most patients across the included studies attended a care facility or hospital and are therefore likely to be unrepresentative of noma cases in the community. Furthermore, many included publications described survivors of noma, with a likelihood of selection bias for surgical rehabilitation studies by expert teams.^2,49^

Although the reported mortality associated with noma has decreased substantially over the last 100 years, the morbidity associated with post-noma complications among survivors has increased the global burden of this disease. Mental health and psychosocial management of patients post-noma is required due to complications, changes in physical appearance, functional debilitation and social stigma, yet such management is severely underreported in the literature included in this review.^50^ Facial reconstruction can reduce this burden, but often requires multiple surgeries, which are out of reach for the majority of noma patients because of their limited availability and high costs. Given the disease disproportionately affects vulnerable populations and predominately children, stigmatisation and isolation due to post-noma deformity is often in itself a death sentence or for life-long suffering, even when patients survive.

### Burden of disease

There are large variations in the quality of evidence provided by primary source estimates of incidence and prevalence reported. Much of the burden of disease data, with similar estimates in two recent reviews,^4,6^ is obtained from retrospective hospital records, precluding the robust estimation of incidence. Other studies report noma prevalence from hospital based or local cross-sectional surveys, which vary greatly within and across countries, or as a complication in cohorts of patients suffering from other infectious diseases. Evidence suggests noma occurs in clusters within countries, and there is significant heterogeneity in the estimated incidence, observed number of reported cases, and risk factors that often precede the disease. ^4,6,51^

### Limitations

The results provide an overview of all treatments described across the literature, and cannot be used to estimate the proportion of interventions as used today. Techniques for the culture of microorganisms were not reported consistently or clearly. Where reported, the timing of when samples were collected varied from days to months after the first symptoms, further making comparisons challenging. Therefore, the evidence to support a clear association between noma and any one microorganism is extremely weak. The vast number of proposed risk factors and the heterogeneity of terminologies and categorisation substantially impacted evidence synthesis. Although risk factors captured here have been observed in at least one noma patient, more often, the factors reported related to the collective study population - the prevalence or combination of factors at the individual patient level were not evidenced. Largely due to when they were published, few studies explicitly reported noma stage using the 2016 WHO definitions. In publications where WHO staging was not used, extrapolation of noma phase was attempted, and judgements made through careful consideration of reported symptoms and diagnosis. Unless clearly reported or evidenced between conference abstracts and subsequent publications, it was not possible to accurately determine if patients are represented and, therefore, double-counted across publications. The literature is hardly representative of all noma cases, mostly representing those patients who attend healthcare facilities and are subsequently reported – this cohort is conservatively estimated as less than 30% of noma cases.^20^ By necessity, there are severe limitations in interpreting and generalising reported deaths and rates of post-noma complications as described above. All results should be interpreted with caution, with these limitations taken into consideration.

## CONCLUSION

This systematic review highlights a dearth of high-quality, comprehensive research studies which extend our understanding of noma aetiology, pathogenesis, prevention and treatment. Limited evidence, therefore, exists to inform future public health programs and assist the development of effective noma control strategies and interventions. Greater surveillance and well-designed epidemiological studies are required globally. The recent inclusion of noma in the WHO NTD list may help prioritise urgently needed research to address the many knowledge gaps about this disease, which disproportionately afflicts the poorest in society. In comparison to other NTDs, noma is among the neglected of the neglected.

## Supporting information

SF1.Supplementary File 1_LitSearchStrategies

SF2. Supplementary File 2_Data_LitScreening

SF3. Supplementary File 3_Extended Results

## Data Availability

The project contains the following underlying and extended data provided in supplementary files to this publication:
Supplementary File 1 (SF1): including literature search strategies, search terms by database, search results, PRISMA diagram, PRISMA Checklist
Supplementary File 2 (SF2): including list of included studies, data and variable dictionary, results data associated with this article, studies excluded at full-text screening stage with reasons.
Supplementary File 3 (SF3): extended results
Copies of all supplementary files, custom scripts and data associated with the protocol publication and this article will also be made accessible online following publication at Open Science Framework: Extended data for Protocol for a systematic review of the evidence-based knowledge on the distribution, associated risk factors, the prevention and treatment modalities for noma, https://doi.org/10.17605/OSF.IO/3DN6G 12

https://doi.org/10.17605/OSF.IO/3DN6G

## CONTRIBUTIONS

**Conceptualisation:** Brittany J. Maguire, Philippe J. Guérin, Benoit Varenne.

**Funding acquisition:** Benoit Varenne, Yuka Makino, Philippe J. Guérin.

**Methodology:** Brittany J. Maguire, Poojan Shrestha, Elinor Harriss.

**Project administration:** Brittany J. Maguire, Yuka Makino.

**Supervision:** Brittany J. Maguire, Philippe J. Guérin.

**Data curation:** Rujan Shrestha, Roland Ngu, Lionel Nizigama, Sumayyah Rashan, Brittany J. Maguire.

**Investigation:** Brittany J. Maguire, Rujan Shrestha, Roland Ngu, Lionel Nizigama, Sumayyah Rashan.

**Formal analysis:** Prabin Dahal, Brittany J. Maguire.

**Validation:** Prabin Dahal, Brittany J. Maguire.

**Visualisation:** Prabin Dahal.

**Writing – original draft:** Brittany J. Maguire, Prabin Dahal, Philippe J. Guérin.

**Writing – review & editing:** Brittany J. Maguire, Sumayyah Rashan, Poojan Shrestha, Rujan Shrestha, Lionel Nizigama, Roland Ngu, Prabin Dahal, Elinor Harriss, Paul N. Newton, Yuka Makino, Benoit Varenne, Philippe J. Guérin.

## DATA SHARING

The project contains the following underlying and extended data provided in supplementary files to this publication:

**Supplementary File 1 (SF1)**: including literature search strategies, search terms by database, search results, PRISMA diagram, PRISMA Checklist

**Supplementary File 2 (SF2)**: including list of included studies, data and variable dictionary, results data associated with this article, studies excluded at full-text screening stage with reasons.

**Supplementary File 3 (SF3):** extended results

Copies of all supplementary files, custom scripts and data associated with the protocol publication and this article will also be made accessible online following publication at Open Science Framework: Extended data for ‘Protocol for a systematic review of the evidence-based knowledge on the distribution, associated risk factors, the prevention and treatment modalities for noma’, https://doi.org/10.17605/OSF.IO/3DN6G 12

Data are available under the terms of the Creative Commons Zero “No rights reserved” data waiver (CC0 1.0 Public domain dedication).

## ACKNOWLEDGEMENTS

Special thanks to Charvy Narain for editorial support.

## BOX

**Box 1 Key findings and recommendations**

| Key Findings | Recommendations |
| --- | --- |
| The number of noma cases reported since 2000 demonstrates the need to reconsider the existing understanding that noma is most prevalent in the “noma belt” region, with a geographic distribution observed in African countries outside this area and other regions including Asia and the Americas. | Greater surveillance and well-designed epidemiological studies should be conducted globally. Education and awareness efforts of noma, particularly the training of primary care workers and community actors should be expanded globally beyond high-risk countries. |
| There are large variations across the few reported burden of disease indicators with most estimates extrapolated from retrospective studies, studies with small sample size, and local surveys. | To improve the understanding of the global distribution and burden of noma more robust, population-based epidemiological studies need to extend beyond the “noma belt” region. Consider promoting systematic reporting at a national level to integrate noma into active surveillance in addition to passive surveillance. |
| The definition and stages of noma progression are poorly standardised and reported across the literature. | Greater awareness of the WHO’s five key stages of noma and use of these criteria in both the case definition and reporting is highly recommended. |
| There is substantial heterogeneity in the risk factors reported to precede noma. The current level of evidence for these risk factors and often the lack of reporting at the individual patient level limits assessment of their causality and interdependency with the development and progression of noma. | • Risk factors and patient comorbidities should be collected and reported at the individual patient level. New studies should be designed to robustly examine the association between such hypothesised risk factors and noma onset. Appropriate statistical methods should be adopted to adjust for confounding when investigating the relationship between hypothesised risk factor and disease acquisition.<br>• Discussion within the noma research community on reporting guidance is advised to encourage standardisation and inclusion of details (such as risk factors and patient’s country of origin), which are often omitted in prospective publications. More robust reporting alongside the clinical categorization of patients may assist in providing additional context and data needed to address several noma research questions. |
| Infectious diseases as a possible risk factor for noma were detailed in 52% of included publications. | Existing health infrastructure, disease prevention and control, vaccination and management programmes for these associated infectious diseases should be considered in future initiatives for effective awareness, case identification, treatment and referral for noma. |
| Malnutrition was the most frequently reported word across the included publications when examining risk factors. | Programmes should incorporate or align with existing infrastructure which targets nutritional status, growth monitoring and intervention. |
| Paucity of information and limited reporting on the microbiology of noma. The association and role of different microbiology in the development and progression of noma is unclear with 117 different microorganisms | • Urgent demand for basic science research on the microbiology, pathophysiology, pathobiology, and histopathologic features of noma.<br>• More detailed reporting of the culture techniques and |
| observed across reported noma cases. | methodologies for identifying microorganisms is needed.<br>Investigation of the spectrum of microorganisms at different phases of noma if/when patient progresses through each stage. |
| Case reports showed early intervention during the acute necrotizing gingivitis stage with proper oral hygiene, debridement, antibiotics therapy, and chlorhexidine mouthwash leads to proper wound healing and no long-term adverse effects. However, there are no robust clinical studies assessing the efficacy of preventative interventions. | Programs and strategies targeted towards increasing disease knowledge and early diagnosis are key to preventing more severe stage progression. Includes encouraging early reporting and rapid referral.<br>Future research including well-designed cohort studies are advised to gain greater evidence in support of recommended preventative interventions.<br>Utilising existing health infrastructure, cross-cutting initiatives or routine assessments for the collection of noma epidemiological data, awareness, and education. |
| There are very few high-quality studies which assess treatment modalities for noma. There is wide heterogeneity in modalities reported and insufficient reporting of treatment by disease stage.<br>Even if individual patient data were collected across the included studies describing antibiotic therapy, there is too much heterogeneity in the treatments, dosing regimens and outcome measures to assess the most effective antibiotic treatment protocols. | Future research should look to more robust study designs, enrolling larger numbers of patients, clearly report treatment outcomes by stage of noma.<br>Discussion within the noma research community on core outcome measures, treatment protocols by disease stage, minimum and consistent follow-up timepoints and further development of standard case report forms may support prospective data collection conducive to meta-analysis across different research studies.<br>The WHO advice for the early detection and management of noma are recommended until further studies indicate a better approach.<sup>13</sup> |
| Reported noma case fatality rate has decreased substantially over the last 100 years. However, there is substantial population bias and study design bias (mostly case reports) with most experts conservatively assuming less than 30% of patients affected by noma would reach a care centre. | Community based surveillance to describe the mortality rate of noma is needed. |

