## Supplementary material for "A systematic review of the noma evidence landscape: current knowledge and gaps": SF1.Supplementary File 1_LitSearchStrategies

**SUPPLEMENTARY FILE 1: LITERATURE SEARCH STRATEGIES AND RESULTS**

A systematic review of the associated risk factors and the treatment modalities for noma

### 1. SEARCH DATABASES

Table 1	Databases searched in 2019 and protocol adaptations necessary in 2022

|  | 04/03/19  Search | 07/12/2022  Search |
| --- | --- | --- |
| Database Searches | | |
| Ovid Medline | Yes | Identical strategy |
| Ovid CAB Abstracts | Yes | Identical strategy |
| Ovid Embase | Yes | Identical strategy |
| Ovid Global Health | Yes | Identical strategy |
| Scopus | Yes | Adapted strategy* |
| Web of Science Core Collection | Yes | Adapted strategy* |
| African Index Medicus | Yes | Identical strategy |
| Pascal | Yes | Adapted strategy* |
| Clinicaltrials.gov | Yes | Identical strategy |
| WHO ICTRP | Yes | Identical strategy |
| Website Searches | | |
| African Journals Online | Yes | Not included^+^ |
| Open Grey | Yes | Not Included^+^ |

*The search strategies for these databases have been adapted due to changes in the databases since 2019: Scopus; Web of Science Core Collection; Pascal. The Scopus search strategy has been adapted due to changes in the database.

^+^A protocol deviation for the 2022 searches was necessary and formal searches of these websites were excluded due to substantial changes to these resources and their respective search functions since 2019.

### 2. SEARCH STRATEGIES

The search strategies by database for both the 2019 and 2022 searches are detailed below.

#### 2022 Search Strategies

Search strategies run on 04/03/19 by Eli Harriss, a librarian at the Bodleian Health Care Libraries, University of Oxford.

**Database: Medline (Ovid MEDLINE® Epub Ahead of Print, In-Process & Other Non-Indexed Citations, Ovid MEDLINE® Daily and Ovid MEDLINE®) 1946 to present**

Search Strategy:

--------------------------------------------------------------------------------

1	noma/ (537)

2	noma.ti,ab. (664)

3	"cancrum oris".ti,ab. (209)

4	"necrotising ulcerative stomatitis".ti,ab. (1)

5	"necrotizing ulcerative stomatitis".ti,ab. (10)

6	"necrotising stomatitis".ti,ab. (9)

7	"necrotizing stomatitis".ti,ab. (45)

8	"gangrenous stomatitis".ti,ab. (36)

9	"cancrus oris".ti,ab. (0)

10	"stomatite ulcéro-necrotique".ti,ab. (0)

11	"stomatite nécrosante".ti,ab. (0)

12	"stomatite gangreneuse".ti,ab. (0)

13	"gangrène à fusospirochètes".ti,ab. (0)

14	1 or 2 or 3 or 4 or 5 or 6 or 7 or 8 or 9 or 10 or 11 or 12 or 13 (936)

15	exp animals/ not humans.sh. (5072762)

16	14 not 15 (889)

**Database: CAB Abstracts <1910 to 2022 Week 48>**

Search Strategy:

--------------------------------------------------------------------------------

1	noma/ (19)

2	noma.ti,ab. (91)

3	"cancrum oris".ti,ab. (18)

4	"necrotising ulcerative stomatitis".ti,ab. (0)

5	"necrotizing ulcerative stomatitis".ti,ab. (2)

6	"necrotising stomatitis".ti,ab. (5)

7	"necrotizing stomatitis".ti,ab. (15)

8	"gangrenous stomatitis".ti,ab. (4)

9	"cancrus oris".ti,ab. (0)

10	"stomatite ulcéro-necrotique".ti,ab. (0)

11	"stomatite nécrosante".ti,ab. (0)

12	"stomatite gangreneuse".ti,ab. (0)

13	"gangrène à fusospirochètes".ti,ab. (0)

14	1 or 2 or 3 or 4 or 5 or 6 or 7 or 8 or 9 or 10 or 11 or 12 or 13 (119)

**Database: Embase 1974 to present**

Search Strategy:

--------------------------------------------------------------------------------

1	"cancrum oris".ti,ab. (178)

2	"necrotising ulcerative stomatitis".ti,ab. (3)

3	"necrotizing ulcerative stomatitis".ti,ab. (11)

4	"necrotising stomatitis".ti,ab. (10)

5	"necrotizing stomatitis".ti,ab. (43)

6	"gangrenous stomatitis".ti,ab. (24)

7	"cancrus oris".ti,ab. (0)

8	"stomatite ulcéro-necrotique".ti,ab. (0)

9	"stomatite nécrosante".ti,ab. (0)

10	"stomatite gangreneuse".ti,ab. (0)

11	"gangrène à fusospirochètes".ti,ab. (0)

12	noma.mp. (1225)

13	1 or 2 or 3 or 4 or 5 or 6 or 7 or 8 or 9 or 10 or 11 or 12 (1336)

14	(exp animal/ or animal.hw. or nonhuman/) not (exp human/ or human cell/ or (human or humans).ti.) (6970051)

15	13 not 14 (1256)

**Database: Global Health <1973 to 2022 Week 48>**

Search Strategy:

--------------------------------------------------------------------------------

1	noma/ (55)

2	noma.ti,ab. (108)

3	"cancrum oris".ti,ab. (45)

4	"necrotising ulcerative stomatitis".ti,ab. (1)

5	"necrotizing ulcerative stomatitis".ti,ab. (7)

6	"necrotising stomatitis".ti,ab. (3)

7	"necrotizing stomatitis".ti,ab. (6)

8	"gangrenous stomatitis".ti,ab. (7)

9	"cancrus oris".ti,ab. (0)

10	"stomatite ulcéro-necrotique".ti,ab. (0)

11	"stomatite nécrosante".ti,ab. (0)

12	"stomatite gangreneuse".ti,ab. (0)

13	"gangrène à fusospirochètes".ti,ab. (0)

14	1 or 2 or 3 or 4 or 5 or 6 or 7 or 8 or 9 or 10 or 11 or 12 or 13 (132)

**Scopus 07/12/2022**

TITLE-ABS-KEY ( noma OR "cancrum oris" OR "necrotising ulcerative stomatitis" OR "necrotizing ulcerative stomatitis" OR "necrotising stomatitis" OR "necrotizing stomatitis" OR "gangrenous stomatitis" OR "cancrus oris" OR "stomatite ulcéro-necrotique" OR "stomatite nécrosante" OR "stomatite gangreneuse" OR "gangrène à fusospirochètes" ) AND ( LIMIT-TO ( EXACTKEYWORD , "Human" ) OR LIMIT-TO ( EXACTKEYWORD , "Humans" ) )

**This is the previous version of the search strategy for Scopus, as run on 04/03/2019. The Nonhuman keyword option was not available on 07/12/2022. Human or Humans were selected as keywords on 07/12/2022 to limit to instead to reduce the amount of noise about an acronym, NOMA.**

( TITLE-ABS-KEY ( ( noma OR "cancrum oris" OR "necrotising ulcerative stomatitis" OR "necrotizing ulcerative stomatitis" OR "necrotising stomatitis" OR "necrotizing stomatitis" OR "gangrenous stomatitis" ) ) ) OR ( TITLE-ABS-KEY ( "cancrus oris" OR "stomatite ulcéro-necrotique" OR "stomatite nécrosante" OR "stomatite gangreneuse" OR "gangrène à fusospirochètes" ) ) AND ( EXCLUDE ( EXACTKEYWORD , "Nonhuman" ) )

**Web of Science Core Collection 07/12/2022** [**https://www.webofscience.com/wos/woscc/summary/5e29a2e8-0e77-4341-a42d-217e8d390c13-642975e4/relevance/1**](https://www.webofscience.com/wos/woscc/summary/5e29a2e8-0e77-4341-a42d-217e8d390c13-642975e4/relevance/1)

**noma OR "cancrum oris" OR "necrotising ulcerative stomatitis" OR "necrotizing ulcerative stomatitis" OR "necrotising stomatitis" OR "necrotizing stomatitis" OR "gangrenous stomatitis"** (Topic) and **Surgery** or **Chemistry** or **Dentistry Oral Surgery Medicine** or **Neurosciences Neurology** or **Cardiovascular System Cardiology** or **Oncology** or **Pediatrics** or **Public Environmental Occupational Health** or **Pathology** or **Tropical Medicine** or **Dermatology** or **Infectious Diseases** or **Biochemistry Molecular Biology** or **Research Experimental Medicine** or **Endocrinology Metabolism** or **Cell Biology** or **Microbiology** or **Immunology** or **Pharmacology Pharmacy** or **Obstetrics Gynecology** or **Parasitology** or **Urology Nephrology** or **Health Care Sciences Services** or **Psychology** or **Physiology** or **Life Sciences Biomedicine Other Topics** or **Social Issues** or **Geriatrics Gerontology** or **Respiratory System** or **Psychiatry** or **Women S Studies** or **Nursing** or **Sociology** or **Hematology** or **Reproductive Biology** or **Transplantation** or **Toxicology** or **Virology** or **Theater** or **Anatomy Morphology** or **Anthropology** or **Archaeology** or **Classics** or **Orthopedics** or **Spectroscopy** or **Public Administration** (Research Areas)

**This is the previous version of the search strategy for the Web of Science Core Collection as run on 04/03/2019. Medical and human headings were selected as keywords on 07/12/2022 to limit to instead to reduce the amount of noise about an acronym, NOMA.**

TOPIC: (noma OR "cancrum oris" OR "necrotising ulcerative stomatitis" OR "necrotizing ulcerative stomatitis" OR "necrotising stomatitis" OR "necrotizing stomatitis" OR "gangrenous stomatitis")

Timespan: All years. Indexes: SCI-EXPANDED, SSCI, A&HCI, CPCI-S, CPCI-SSH, BKCI-S, BKCI-SSH, ESCI, CCR-EXPANDED, IC.

**African Index Medicus -** <http://search.bvsalud.org/ghl/?lang=en> - limited to one selected WHO regional database – AIM (AFRO)

(tw:(noma OR "cancrum oris" OR "necrotising ulcerative stomatitis" OR "necrotizing ulcerative stomatitis" OR "necrotising stomatitis" OR "necrotizing stomatitis" OR "gangrenous stomatitis"))

OR

(tw: "cancrus oris" OR "stomatite ulcéro-necrotique" OR "stomatite nécrosante" OR "stomatite gangreneuse" OR "gangrène à fusospirochètes")

**Pascal -** [**http://pascal-francis.inist.fr/vibad/index.php?action=advancedSearch**](http://pascal-francis.inist.fr/vibad/index.php?action=advancedSearch) **07/12/2022**

ti.\*:(noma OR "cancrum oris" OR "necrotising ulcerative stomatitis" OR "necrotizing ulcerative stomatitis" OR "necrotising stomatitis" OR "necrotizing stomatitis" OR "gangrenous stomatitis") OR ti.\*:("cancrus oris" OR "stomatite ulcéro-necrotique" OR "stomatite nécrosante" OR "stomatite gangreneuse" OR "gangrène à fusospirochètes") OR kw.\*:("NOMA")

**Pascal -** [**http://pascal-francis.inist.fr/vibad/index.php?action=advancedSearch**](http://pascal-francis.inist.fr/vibad/index.php?action=advancedSearch)

**This is the previous version of the search strategy for Pascal as run on 04/03/2019.**

1. title.\*:(noma OR "cancrum oris" OR "necrotising ulcerative stomatitis" OR "necrotizing ulcerative stomatitis" OR "necrotising stomatitis" OR "necrotizing stomatitis" OR "gangrenous stomatitis") OR abstract.\*:(noma OR "cancrum oris" OR "necrotising ulcerative stomatitis" OR "necrotizing ulcerative stomatitis" OR "necrotising stomatitis" OR "necrotizing stomatitis" OR "gangrenous stomatitis")
2. title.\*:("cancrus oris" OR "stomatite ulcéro-necrotique" OR "stomatite nécrosante" OR "stomatite gangreneuse" OR "gangrène à fusospirochètes") OR abstract.\*:("cancrus oris" OR "stomatite ulcéro-necrotique" OR "stomatite nécrosante" OR "stomatite gangreneuse" OR "gangrène à fusospirochètes")
3. keyword.\*:(noma)
4. 1 OR 2 OR 3

**Clinicaltrials.gov** [**https://clinicaltrials.gov/ct2/search/advanced?cond=&term=&cntry=&state=&city=&dist**](https://clinicaltrials.gov/ct2/search/advanced?cond=&term=&cntry=&state=&city=&dist)**=**

Condition or disease: noma OR cancrum oris OR necrotising ulcerative stomatitis OR necrotizing ulcerative stomatitis OR necrotising stomatitis OR necrotizing stomatitis OR gangrenous stomatitis

**WHO ICTRP** <http://apps.who.int/trialsearch/Default.aspx> (Phases are ALL)

noma OR cancrum oris OR necrotising ulcerative stomatitis OR necrotizing ulcerative stomatitis OR necrotising stomatitis OR necrotizing stomatitis OR gangrenous stomatitis

#### 2019 Search Strategies

Search strategies run on 04/03/19 by Eli Harriss, a librarian at the Bodleian Health Care Libraries, University of Oxford.

**Database: Medline (Ovid MEDLINE® Epub Ahead of Print, In-Process & Other Non-Indexed Citations, Ovid MEDLINE® Daily and Ovid MEDLINE®) 1946 to present**

Search Strategy:

--------------------------------------------------------------------------------

1	noma/ (478)

2	noma.ti,ab. (460)

3	"cancrum oris".ti,ab. (187)

4	"necrotising ulcerative stomatitis".ti,ab. (1)

5	"necrotizing ulcerative stomatitis".ti,ab. (9)

6	"necrotising stomatitis".ti,ab. (7)

7	"necrotizing stomatitis".ti,ab. (40)

8	"gangrenous stomatitis".ti,ab. (35)

9	"cancrus oris".ti,ab. (0)

10	"stomatite ulcéro-necrotique".ti,ab. (0)

11	"stomatite nécrosante".ti,ab. (0)

12	"stomatite gangreneuse".ti,ab. (0)

13	"gangrène à fusospirochètes".ti,ab. (0)

14	1 or 2 or 3 or 4 or 5 or 6 or 7 or 8 or 9 or 10 or 11 or 12 or 13 (728)

15	exp animals/ not humans.sh. (4552221)

16	14 not 15 (687)

**Database: CAB Abstracts <1910 to 2019 Week 08>**

Search Strategy:

--------------------------------------------------------------------------------

1	noma/ (13)

2	noma.ti,ab. (138)

3	"cancrum oris".ti,ab. (89)

4	"necrotising ulcerative stomatitis".ti,ab. (0)

5	"necrotizing ulcerative stomatitis".ti,ab. (2)

6	"necrotising stomatitis".ti,ab. (4)

7	"necrotizing stomatitis".ti,ab. (14)

8	"gangrenous stomatitis".ti,ab. (15)

9	"cancrus oris".ti,ab. (0)

10	"stomatite ulcéro-necrotique".ti,ab. (0)

11	"stomatite nécrosante".ti,ab. (0)

12	"stomatite gangreneuse".ti,ab. (0)

13	"gangrène à fusospirochètes".ti,ab. (0)

14	1 or 2 or 3 or 4 or 5 or 6 or 7 or 8 or 9 or 10 or 11 or 12 or 13 (232)

**Database: Embase 1974 to present**

Search Strategy:

--------------------------------------------------------------------------------

1	"cancrum oris".ti,ab. (149)

2	"necrotising ulcerative stomatitis".ti,ab. (2)

3	"necrotizing ulcerative stomatitis".ti,ab. (11)

4	"necrotising stomatitis".ti,ab. (7)

5	"necrotizing stomatitis".ti,ab. (36)

6	"gangrenous stomatitis".ti,ab. (24)

7	"cancrus oris".ti,ab. (0)

8	"stomatite ulcéro-necrotique".ti,ab. (0)

9	"stomatite nécrosante".ti,ab. (0)

10	"stomatite gangreneuse".ti,ab. (0)

11	"gangrène à fusospirochètes".ti,ab. (0)

12	noma.mp. (750)

13	1 or 2 or 3 or 4 or 5 or 6 or 7 or 8 or 9 or 10 or 11 or 12 (851)

14	(exp animal/ or animal.hw. or nonhuman/) not (exp human/ or human cell/ or (human or humans).ti.) (6053957)

15	13 not 14 (788)

**Database: Global Health <1973 to 2019 Week 08>**

Search Strategy:

--------------------------------------------------------------------------------

1	noma/ (44)

2	noma.ti,ab. (77)

3	"cancrum oris".ti,ab. (35)

4	"necrotising ulcerative stomatitis".ti,ab. (1)

5	"necrotizing ulcerative stomatitis".ti,ab. (7)

6	"necrotising stomatitis".ti,ab. (2)

7	"necrotizing stomatitis".ti,ab. (6)

8	"gangrenous stomatitis".ti,ab. (6)

9	"cancrus oris".ti,ab. (0)

10	"stomatite ulcéro-necrotique".ti,ab. (0)

11	"stomatite nécrosante".ti,ab. (0)

12	"stomatite gangreneuse".ti,ab. (0)

13	"gangrène à fusospirochètes".ti,ab. (0)

14	1 or 2 or 3 or 4 or 5 or 6 or 7 or 8 or 9 or 10 or 11 or 12 or 13 (100)

**Scopus**

( TITLE-ABS-KEY ( ( noma OR "cancrum oris" OR "necrotising ulcerative stomatitis" OR "necrotizing ulcerative stomatitis" OR "necrotising stomatitis" OR "necrotizing stomatitis" OR "gangrenous stomatitis" ) ) ) OR ( TITLE-ABS-KEY ( "cancrus oris" OR "stomatite ulcéro-necrotique" OR "stomatite nécrosante" OR "stomatite gangreneuse" OR "gangrène à fusospirochètes" ) ) AND ( EXCLUDE ( EXACTKEYWORD , "Nonhuman" ) )

**Web of Science Core Collection**

TOPIC: (noma OR "cancrum oris" OR "necrotising ulcerative stomatitis" OR "necrotizing ulcerative stomatitis" OR "necrotising stomatitis" OR "necrotizing stomatitis" OR "gangrenous stomatitis")

Timespan: All years. Indexes: SCI-EXPANDED, SSCI, A&HCI, CPCI-S, CPCI-SSH, BKCI-S, BKCI-SSH, ESCI, CCR-EXPANDED, IC.

**African Index Medicus -** <http://search.bvsalud.org/ghl/?lang=en> - limited to one selected WHO regional database – AIM (AFRO)

(tw:(noma OR "cancrum oris" OR "necrotising ulcerative stomatitis" OR "necrotizing ulcerative stomatitis" OR "necrotising stomatitis" OR "necrotizing stomatitis" OR "gangrenous stomatitis"))

OR

(tw: "cancrus oris" OR "stomatite ulcéro-necrotique" OR "stomatite nécrosante" OR "stomatite gangreneuse" OR "gangrène à fusospirochètes")

**Pascal -** [**http://pascal-francis.inist.fr/vibad/index.php?action=advancedSearch**](http://pascal-francis.inist.fr/vibad/index.php?action=advancedSearch)

1. title.\*:(noma OR "cancrum oris" OR "necrotising ulcerative stomatitis" OR "necrotizing ulcerative stomatitis" OR "necrotising stomatitis" OR "necrotizing stomatitis" OR "gangrenous stomatitis") OR abstract.\*:(noma OR "cancrum oris" OR "necrotising ulcerative stomatitis" OR "necrotizing ulcerative stomatitis" OR "necrotising stomatitis" OR "necrotizing stomatitis" OR "gangrenous stomatitis")
2. title.\*:("cancrus oris" OR "stomatite ulcéro-necrotique" OR "stomatite nécrosante" OR "stomatite gangreneuse" OR "gangrène à fusospirochètes") OR abstract.\*:("cancrus oris" OR "stomatite ulcéro-necrotique" OR "stomatite nécrosante" OR "stomatite gangreneuse" OR "gangrène à fusospirochètes")
3. keyword.\*:(noma)
4. 1 OR 2 OR 3

**Clinicaltrials.gov** [**https://clinicaltrials.gov/ct2/search/advanced?cond=&term=&cntry=&state=&city=&dist**](https://clinicaltrials.gov/ct2/search/advanced?cond=&term=&cntry=&state=&city=&dist)**=**

Condition or disease: noma OR cancrum oris OR necrotising ulcerative stomatitis OR necrotizing ulcerative stomatitis OR necrotising stomatitis OR necrotizing stomatitis OR gangrenous stomatitis

**WHO ICTRP** <http://apps.who.int/trialsearch/Default.aspx> (Phases are ALL)

noma OR cancrum oris OR necrotising ulcerative stomatitis OR necrotizing ulcerative stomatitis OR necrotising stomatitis OR necrotizing stomatitis OR gangrenous stomatitis

WEBSITE SEARCHES

**African journals online** <https://www.ajol.info/> was manually searched by Brittany Maguire on 4^th^ March 2019 using both English and French search terms. A total of 48 records were identified. After de-duplication a total of 42 unique citations were included for abstract screening.

ENGLISH TERMS

noma OR "cancrum oris" OR "necrotising ulcerative stomatitis" OR "necrotizing ulcerative stomatitis" OR "necrotising stomatitis" OR "necrotizing stomatitis" OR "gangrenous stomatitis"

FRENCH TERMS

noma OR cancrus oris OR stomatite ulcéro-necrotique OR stomatite nécrosante OR stomatite gangreneuse OR gangrène à fusospirochètes

GREY LITERATURE SEARCH

**Open Grey** <http://www.opengrey.eu/> was manually searched by BJM on 4^th^ March 2019 using both English and French search terms. A total of 57 records were identified (n=28 English search terms; n=29 French search terms). After de-duplication a total of 29 unique citations were included for abstract screening.

ENGLISH TERMS

noma OR "cancrum oris" OR "necrotising ulcerative stomatitis" OR "necrotizing ulcerative stomatitis" OR "necrotising stomatitis" OR "necrotizing stomatitis" OR "gangrenous stomatitis"

FRENCH TERMS

noma OR cancrus oris OR stomatite ulcéro-necrotique OR stomatite nécrosante OR stomatite gangreneuse OR gangrène à fusospirochètes

**WHO Grey literature**

Grey literature and WHO internal materials as well as papers published on noma was sent to IDDO on 17^th^ April to supplement the Systematic Review. A total of 94 files were received.

### 4. ELIGIBILITY CRITERIA FOR SCREENING

Table 2	Study inclusion and exclusion criteria

| **Selection Criterion** | **Inclusion criteria** | **Exclusion criteria** |
| --- | --- | --- |
| Population | Patients of all ages diagnosed with noma* | - Studies dealing with non-noma illnesses or conditions - Animal studies - Studies dealing with noma-like illness and noma neonatorum - All publications that do not include noma patients |
| Study Design | All primary research studies   - clinical trials - cohort studies - case-control - cross-sectional - other observational studies - case reports and case series | - Non-primary research studies   - Opinion pieces   - Clinical guidelines   - Text-book chapters   - Letters etc. - Qualitative surveys - Conference proceedings   - conference abstract published prior to 2017 (≤2016)   - conference abstracts 2017-2022 that have been subsequently published |
| Intervention | *The search strategy was not restricted by intervention, comparator or outcomes given the desire to review studies evaluating all noma treatment modalities, as well as capturing non-intervention studies* | |
| Comparators |  |  |
| Outcome |  |  |
| Language and date | - Publications in English/French/Chinese were included for comprehensive data extraction and subsequent synthesis | - Publications not in English/French/Chinese have been identified but excluded from data extraction and further analysis |

*During literature screening, publications were included if they used any term synonymous for noma, e.g.: "cancrum oris" OR "necrotising ulcerative stomatitis" OR "necrotizing ulcerative stomatitis" OR "necrotising stomatitis" OR "necrotizing stomatitis" OR "gangrenous stomatitis" OR "oro-facial gangrene" OR “trench mouth”

### 5. LITERATURE SEARCH RESULTS

#### Results of literature searches

**2019 Search**

The original search strategies were run on and inclusive of 4 March 2019. The African Journals Online (AJOL) database and OpenGrey.eu were manually searched by a member of the review team also on 4 March 2019. The number of records identified from these searches by data source is summarised in Table 3.

The database searches identified 6,931 records of which 3,934 unique citations were included for abstract screening after deduplication. The number of unique citations identified from the manual AJOL and OpenGrey searches were 42 and 29, respectively. Additionally, grey literature comprising 94 papers were provided to the review team on 17 April 2019 by the WHO Global Oral Health Programme.

A total of 3,934 records were screened by two independent reviewers against the pre-specified study eligibility criteria (Table 2).

**2022 Update Search**

The update literature searches were conducted on 7th December 2022. Due to changes with some search databases between 2019 and 2022, minor adaptations were necessary for some database search strings. These are outlined above. The database searches identified 4,963 records of which there were 856 unique citations since 4 March 2019 which were included for abstract screening after deduplication.

#### Results of the literature screening

The PRISMA flow diagram and summary of all literature screening is provided in Figure 1.

**2019 Search**

Of the 3,934 records screened and identified in 2019, a total of 3,004 records could be excluded from the abstract and title alone. The full-texts of 930 records were required for review to assess eligibility for inclusion. A total of 794 full-text articles have been reviewed by two independent researchers of which 293 have been included in the analysis. As detailed further below, there are 136 missing papers from the 2019 searches which could not be retrieved.

**2022 Update Search**

After deduplication, a total of 874 records were screened from the 2022 update searches. 683 records could be excluded on the abstract and title alone. The full-texts of 191 records were required for review to assess eligibility for inclusion. A total of 163 full-text articles have been reviewed by two independent researchers of which 78 have been included in the analysis presented. As detailed further below, there are 28 missing papers from the 2022 searches which could not be retrieved.

**Languages**

Both English and French full-text publications were included in this analysis. Although Chinese publications were eligible for inclusion none were identified that met the eligibility criteria. A total of 63 publications that appeared to meet the eligibility criteria were excluded from further analysis at the full-text review stage due to no available resourcing for translation. The languages of these 63 publications included German, Dutch, Spanish, Italian, Greek, Japanese and Russian.

**Systematic Review Search**

We identified 6 systematic reviews of high quality and relevance during the literature searches. We reviewed the bibliography of these 6 systematic reviews and cross-checked for any references that met the eligibility criteria of our review that had not already been. An additional 20 studies that met our eligibility were identified and have been included for data extraction and analysis.

Table 3	Summary of the literature search sources and number of records identified from the searches

| **Database or Source** | **Number of records identified** | |
| --- | --- | --- |
|  | **Original Search Results^a^** | **Update**  **Search Results^b^** |
| FORMAL DATABASE SEARCHES | |  |
| Ovid Medline | 687 | 889 |
| Ovid CAB Abstracts | 232 | 119 |
| Ovid Embase | 788 | 1,256 |
| Ovid Global Health | 100 | 132 |
| Scopus | 2,824 | 1,135^e^ |
| Web of Science Core Collection | 2,019 | 940^e^ |
| African Index Medicus | 14 | 14 |
| Pascal | 256 | 207^e^ |
| Clinicaltrials.gov | 5 | 1 |
| WHO ICTRP | 6 | 0 |
| TOTAL | 6,931 | 4,693 |
| **Total after deduplication** | **3,769** |  |
| **Unique since 04/03/2019** |  | **854** |
| WEBSITE SEARCHES | |  |
| African Journals Online (AJOL)^c^ | 48 | N/A |
| **Total after deduplication** | **42** | **-** |
| MANUAL LITERATURE SEARCHES | |  |
| OpenGrey.eu^c^ | 57 | N/A |
| WHO Grey Literature Shared^d^ | 94 | N/A |
| Systematic Review Bibliography Searches | - | 20 |
| **Total after deduplication** | **123** | **-** |
| **Total screened in original search** | **3,934** |  |
| **Total screened in update search** |  | **874** |
| **TOTAL SCREENED** | **4,808** | |

a Searches conducted by librarian at Oxford Bodleian Health Care Libraries on 4 March 2019 unless otherwise specified

b Searches conducted by librarian at Oxford Bodleian Health Care Libraries on 7 December 2022

c Searches conducted manually by the research team on 4 March 2019

d Provided by WHO Global Oral Health Programme on 17 April 2019

e The search strategies for these databases have been adapted due to changes in the databases since 2019: Scopus; Web of Science Core Collection; Pascal. The Scopus search strategy has been adapted due to changes in the database.

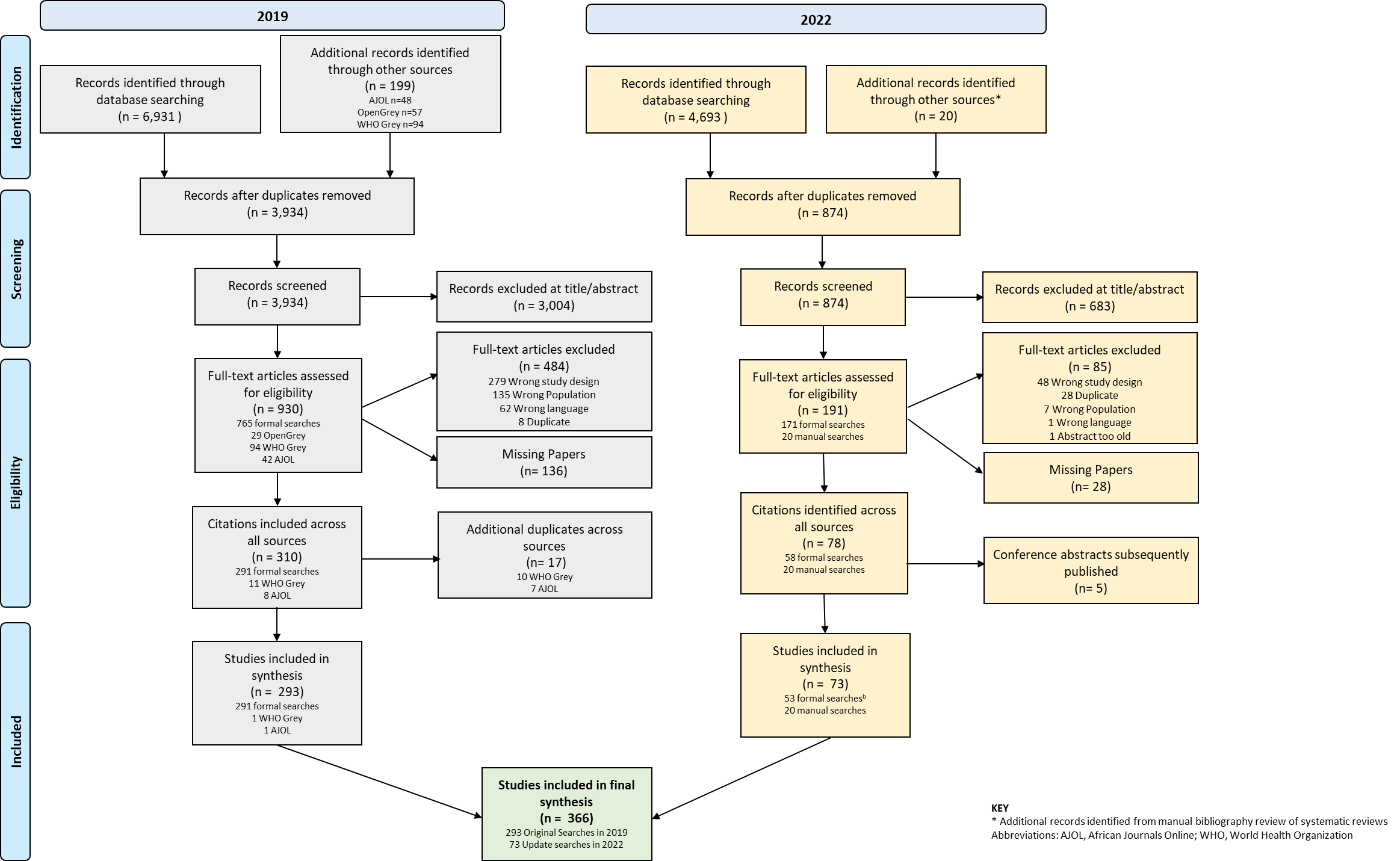

Figure 1	PRISMA flow diagram of the noma systematic review

#### Missing papers

Following the title and abstract screening, there were a total of 164 potentially relevant citations for which the full-text has not been located (2019 n=136; 2022 n=28). This was despite exhaustive investigation including additional support from the WHO library in October 2019 and again in February 2023. Therefore, a definitive inclusion or exclusion cannot be made. Conservatively and in the absence of full-text publications, these 164 citations were excluded from further analysis and were not included in the results. However, work has been conducted to categorise these citations (Table 4).

Requests to experts in the field and authors of missing papers were submitted. It is important to note that the main reason for not retrieving these papers is that they are published in national journals not available or accessible to the Oxford University library or partner libraries including the WHO library, despite very large journal subscriptions.

Table 4	Missing paper summary and likely decision on available information

| **Decision based on the available information** | **Number of publications (N=164)** |
| --- | --- |
| Abstract Available | 49 |
| Likely include on abstract* | 26 |
| Unclear from abstract | 11 |
| Likely exclude on abstract | 12 |
| Citation Only | 115 |
| Likely include on title alone* | 28 |
| Unclear | 87 |

* Denotes the abstracts/citations from which data extraction has been conducted with the available information

#### Included studies

A total of 366 publications definitively included with full-text available have been included in the synthesis of evidence for this publication. A complete list of the included studies can be found below.

### 6. LIST OF INCLUDED STUDIES

1. Adams-Ray, W. E., & James, J. H. (1992). Cancrum oris. Functional and Cosmetic Reconstruction in Patients with Ankylosis of the Jaws. British Journal of Plastic Surgery, 45(3), 193-198.
2. Adedoja, D., Kabue, M. M., & Sahila, P. (2002). Cancrum oris in HIV infected children in Lesotho: report of two cases. East African Medical Journal, 79(9), 499-501.
3. Adekeye, E. O., & Ord, R. A. (1983). Cancrum oris: principles of management and reconstructive surgery. Journal of Maxillofacial Surgery, 11(4), 160-170.
4. Adekeye, E. O., Lavery, K. M., & Nasser, N. A. (1986). The versatility of pectoralis major and latissimus dorsi myocutaneous flaps in the reconstruction of cancrum oris defects of children and adolescents. Journal of Maxillofacial Surgery, 14(2), 99-102.
5. Adeniyi, S. A., & Awosan, K. J. (2019). Pattern of noma (cancrum oris) and its risk factors in Northwestern Nigeria: A hospital-based retrospective study. Annals of African Medicine, 18(1), 17-22.
6. Adeola, D. S., & Obiadazie, A. C. (2009). Protocol for managing acute cancrum oris in children: an experience in five cases. African Journal of Paediatric Surgery: AJPS, 6(2), 77-81.
7. Adeola, D. S., Eguma, S. A., & Ononiwu, C. N. (2004). Cancrum oris among Nigerian children. Nigerian Journal of Surgical Research, 6(1-2).
8. Adolph, H. P., Yugueros, P., & Woods, J. E. (1996). Noma: a review. Annals of Plastic Surgery, 37(6), 657-668.
9. Agrawal, K., & Panda, K. N. (2001). Moustache reconstruction using an extended midline forehead flap. British Journal of Plastic Surgery, 54(2), 159-161.
10. Aguiar, A. M., Enwonwu, C. O., & Pires, F. R. (2003). Noma (cancrum oris) associated with oral myiasis in an adult. Oral Diseases, 9(3), 158-159.
11. Akula, S. K., Creticos, C. M., & Weldon-Linne, C. M. (1989). Gangrenous stomatitis in AIDS. Lancet, 1(8644), 955.
12. Ampofo, O., & Findlay, G. M. (1950). Oral aureomycin in the treatment of tropical ulcers and cancrum oris. Transactions of the Royal Society of Tropical Medicine & Hygiene, 44(3), 307-310.
13. Anonymous. (1848). Cancrum Oris. The American Journal of Dental Science, 9(1), 135.
14. Anonymous. (1852). Cancrum Oris. The American Journal of Dental Science, 3(1), 112-116.
15. Anonymous. (1870). Gangrenous Stomatitis: History of Two Cases, with Remarks. The American Journal of Dental Science, 3(9), 425-430.
16. Anonymous. (1887). Cancrum Oris. The American Journal of Dental Science, 20(9), 414-415.
17. Anonymous. (1893). A Fatal Case of Noma in an Adult. The American Journal of Dental Science, 26(9), 427.
18. Artis, J. P., Cassang, S., Artis, M., & Bennani, I. (1990). Noma. Apropos of a case in a 10-year old child. Journal de parodontologie, 9(4), 363-367.
19. Arvind, K., Anita, V., Satish, S., Shirley, S., & Krishnarathinam. (2010). Cancrum oris-associated with acute myeloid leukemia: a forgotten disease. International Journal of Head and Neck Surgery, 1(3), 167-169.
20. Asbell, M. B. (1939). Case report no. 26 Gangrenous stomatitis. American Journal of Orthodontics and Oral Surgery, 25(6), 592-596.
21. Auluck, A., & Pai, K. M. (2005). Noma: Life Cycle of a Devastating Sore--Case Report and Literature Review. Journal of the Canadian Dental Association, 71(10).
22. Bagewadi, S. B., Awasthi, U. R., Mody, B. M., Suma, G. N., & Garg, S. (2015). Bony fusion of the maxilla and mandible as a sequelae of noma: A rare case report. Imaging Science in Dentistry, 45(3), 193-198.
23. Bah, A. T., Barry, M., Camara, S. A. T., Cisse, A., & Diallo, O. R. (2009). Le noma chez les enfants infectes par le VIH au CHU de Conakry: 5 cas. Mali Medical, 24(3), 71-74.
24. Bala, M, et al., (2022). Is Blood Transfusion Justified During Soft Tissue Surgery in Noma Patients? A One-Year Appraisal at Noma Children Hospital, Sokoto, Nigeria. Journal of the West African College of Surgeons, 12(2), 1–6.
25. Balaka, B., Bonkoungou, P., Ouedraogo, D., Sawadogo, A. A., & Zoubga, Z. A. (2006). Le noma de l’enfant en milieu hospitalier de Bobo-Dioulasso (Burkina-Faso): aspects epidemiologiques et diagnostiques.
26. Balaka, B., Bonkoungou, P., Sawadogo, A., & Tall, F. R. (2005). Le noma de l’enfant au Burkina-Faso : aspects therapeutiques et evolutifs.
27. Baniulyte, G., et al., (2019). NOMA - a possible presentation in Glasgow. British Journal of Oral & Maxillofacial Surgery, 57(10), e56–e57.
28. Baratti-Mayer, D., Gayet-Ageron, A., & Pittet, D. (2015). Life conditions and dental behavior as risk factors for noma among children in Niger; preliminary results. Antimicrobial Resistance and Infection Control. Conference: 3rd International Conference on Prevention and Infection Control, ICPIC, 4(SUPPL. 1).
29. Baratti-Mayer, D., Gayet-Ageron, A., Cionca, N., Mossi, M. A., Pittet, D., & Mombelli, A. (2017). Acute necrotising gingivitis in young children from villages with and without noma in Niger and its association with sociodemographic factors, nutritional status and oral hygiene practices: results of a population-based survey. BMJ Global Health, 2(3), e000253.
30. Baratti-Mayer, D., Gayet-Ageron, A., Hugonnet, S., Francois, P., Pittet-Cuenod, B., Huyghe, A., … Geneva Study Group on Noma. (2013). Risk factors for noma disease: a 6-year, prospective, matched case-control study in Niger. The Lancet Global Health, 1(2), e87-e96.
31. Baratti-Mayer, D., Pittet, B., & Montandon, D. (2004). GESNOMA (Geneva Study group on Noma): State-of-the-art medical research for humanitarian purposes. [French]. [GESNOMA (Geneva Study group on Noma): Une recherche medicale de pointe a but humanitaire.]. Annales de Chirurgie Plastique et Esthetique, 49(3), 302-305.
32. Barker, T. H. (1852). Case of Cancrum Oris, With Necrosis of A Large Portion of The Inferior Maxillary Bone, Followed by Recovery. Provincial Medical and Surgical Journal, s1-16(24), 611.
33. Barlow, W. F. (1843). CANCRUM ORIS, NOT A RESULT OF MERCURY. ANALOGOUS AFFECTION. The Lancet, 40(1025), 120-121.
34. Barrera, J., & Connor, M. P. (2012). Noma in an Afghani child: A case report. International Journal of Pediatric Otorhinolaryngology, 76(5), 742-744.
35. Barrios, T. J., Aria, A. A., & Brahney, C. (1995). Cancrum oris in an HIV-positive patient. Journal of Oral & Maxillofacial Surgery, 53(7), 851-855.
36. Barthelemy, I., Martin, D., Sannajust, J. P., Marck, K., Pistre, V., & Mondie, J. M. (2002). Prefabricated superficial temporal fascia flap combined with a submental flap in noma surgery. Plastic & Reconstructive Surgery, 109(3), 936-940; discussion 941-932.
37. Baruchin, A. M., Scharf, S., & Nahlieli, O. (1993). Plastic surgery findings in Ethiopian immigrants. Israel Journal of Medical Sciences, 29(6-7), 398-402.
38. Behanan, A. G., Auluck, A., & Pai, K. L. (2004). Cancrum oris. British Journal of Oral & Maxillofacial Surgery, 42(3), 267-269.
39. Bello, S. A. (2012). Gillies fan flap for the reconstruction of an upper lip defect caused by noma: case presentation. Clinical Cosmetic & Investigational Dentistry, 4, 17-20.
40. Bello, S. A., Aluko Olokun, B., Olaitan, A. A., & Ajike, S. O. (2012). Aetiology and presentation of ankylosis of the temporomandibular joint: Report of 23 cases from Abuja, Nigeria. British Journal of Oral and Maxillofacial Surgery, 50(1), 80-84.
41. Bello, S. A., et al., (2019). Estimated incidence and Prevalence of noma in north central Nigeria, 2010-2018: A retrospective study. PLoS neglected tropical diseases, 13(7), e0007574.
42. Bendl, B. J., Padmos, A., Harder, E. J., & McArthur, P. D. (1983). Noma: report of three adult cases. Australasian Journal of Dermatology, 24(3), 115-121.
43. Bertrand, B., et al., (2019). Twenty-Five Years of Experience with the Submental Flap in Facial Reconstruction: Evolution and Technical Refinements following 311 Cases in Europe and Africa. Plastic and reconstructive surgery, 143(6), 1747–1758.
44. Bewley, H. T. (1894). A case of cancrum oris in typhoid fever. Transactions of the Royal Academy of Medicine in Ireland, 12(1), 84-92.
45. Bisseling, P., Bruhn, J., Erdsach, T., Ettema, A. M., Sautter, R., & Berge, S. J. (2010). Long-term results of trismus release in noma patients. International Journal of Oral and Maxillofacial Surgery, 39(9), 873-877.
46. Biswal, N., Mahadevan, S., & Srinivasan, S. (1992). Gangrenous stomatitis following measles. Indian Pediatrics, 29(4), 509-511.
47. Black, F. O. (1973). Experiences of an otolaryngologist in Vietnam. Southern Medical Journal, 66(3), 308-312.
48. Bolivar, I., Whiteson, K., Stadelmann, B., Baratti-Mayer, D., Gizard, Y., Mombelli, A., . . . Geneva Study Group on Noma. (2012). Bacterial diversity in oral samples of children in Niger with acute noma, acute necrotizing gingivitis, and healthy controls. PLoS Neglected Tropical Diseases [electronic resource], 6(3), e1556.
49. Bonkoungou P, et al.,(2006). Le Noma de l’enfant en milieu hospitalier de Bobo-Dioulasso (Burkina Faso): Aspects epidemiolohiques et diagnostiques. Revue internationale du Collège d'Odonto-Stomatologie et de Chirurgie-Maxillo-Faciale, 13(3), 24-27.
50. Borcbakan, C. (1952). Two cases of noma sequelae treated with scalp flaps. Plastic & Reconstructive Surgery, 10(4), 283-287.
51. Bouaoud, J., et al., (2021). Humanitarian Maxillofacial Mission's Success Requires Experienced Surgeons, Careful Planning, and Meeting With the Local's Care Needs. Journal of oral and maxillofacial surgery : official journal of the American Association of Oral and Maxillofacial Surgeons, 79(10), 1999.e1–1999.e9.
52. Bouassalo, K. M., et al., (2019). Chronic lymphocytic leukemia revealed by a rare complication: Noma. First description from Togo. Journal of Oral Medicine and Oral Surgery, 25(3), 31.
53. Bouillat, M., & Ramiandrasoa, A. (1940). Eighteen cases of noma with thirteen cures after treatment by ascorbic acid. [Dix huit gangrenes de la bouche, dont treize gueries, traitees par I'acide ascorbique.]. Bulletin de la Societe de Pathologie Exotique, 33, 208-209.
54. Bouman, M. A., Marck, K. W., Griep, J. E. M., Marck, R. E., Huijing, M. A., & Werker, P. M. N. (2010). Early outcome of noma surgery. Journal of Plastic, Reconstructive and Aesthetic Surgery, 63(12), 2052-2056.
55. Bourgeois, D. M., Diallo, B., Frieh, C., & Leclercq, M. H. (1999). Epidemiology of the incidence of oro-facial noma: a study of cases in Dakar, Senegal, 1981-1993. American Journal of Tropical Medicine & Hygiene, 61(6), 909-913.
56. Brady-West, D. C., Richards, L., Thame, J., Moosdeen, F., & Nicholson, A. (1998). Cancrum oris (noma) in a patient with acute lymphoblastic leukaemia - A complication of chemotherapy induced neutropenia. West Indian Medical Journal, 47(1), 33-34.
57. Braimah, R. O., Taiwo, A. O., Ibikunle, A. A., Gbotolorun, M. O., Adeyemi, M., & Mujtaba, B. (2018). Regional Flaps in Maxillofacial and Oral Soft-Tissue Reconstruction: Experiences and Challenges in a Developing Country. Turkish Journal of Plastic Surgery, 26(4), 169-173.
58. Braimah, R. O., Taiwo, A. O., Ibikunle, A. A., Oladejo, T., Adeyemi, M., Adejobi, A. F., & Abubakar, S. (2018). Temporomandibular Joint Ankylosis with Maxillary Extension: Proposal for Modification of Sawhney's Classification. Craniomaxillofacial Trauma and Reconstruction Open, 2(1), e15-e21.
59. Braun, U., et al., (2020). Anaesthetic care for noma (cancrum oris) - the disease, the airway and how to provide anaesthetic care without a clinical safety infrastructure. Trends in Anaesthesia and Critical Care, 31, 16–20.
60. Brotherton B. J. (2019). Visual Diagnosis: Rapidly Progressing Oral Ulcer in a 13-year-old Girl. Pediatrics in review, 40(9), e32–e34.
61. Brown, J. B. (1933). Types of chronic infection about the mouth Report of cases of noma and of acrodynia. International Journal of Orthodontia and Dentistry for Children, 19(1), 59-62.
62. Buchanan, J. A., Cedro, M., Mirdin, A., Joseph, T., Porter, S. R., & Hodgson, T. A. (2006). Necrotizing stomatitis in the developed world. Clinical & Experimental Dermatology, 31(3), 372-374.
63. Burkwall, H. F. (1942). Noma. American Journal of Orthodontics and Oral Surgery, 28(7), B394-B398.
64. Cantaloube, D. and P. Combemale (1990). What is your diagnosis? Noma or cancrum oris. Annales De Dermatologie Et De Venereologie 117(10): 735-737.
65. Carayon, A., et al.,(1965). "[Plastic Surgery of Noma by Cutaneous Migratory Tubes]." Bulletin de la Societe Medicale d Afrique Noire de Langue Francaise 10: 129-138.
66. Casanas, B., Pothiawala, S., & Sinnott, J. T. (2009). An elderly man with noma orofacial gangrene. Infectious Diseases in Clinical Practice, 17(3), 208-209.
67. CASE OF CANCRUM ORIS. (1841). The Lancet, 36(940), 831.
68. Catros, S., Prudence, M., Lérici, S., & Fricain, J. C. (2014). A case of necrotizing stomatitis: Noma of the rich countries? Medecine Buccale Chirurgie Buccale, 20(1), 27-34.
69. Chaubal, T., & Bapat, R. (2017). Trench mouth. The American Journal of Medicine, 130(11), e493-e494.
70. Chiandussi, S., Luzzati, R., Tirelli, G., Di Lenarda, R., & Biasotto, M. (2009). Cancrum oris in developed countries. Aging-Clinical & Experimental Research, 21(6), 475-477.
71. Chidzonga, M. M. (1996). Noma (cancrum oris) in human immunodeficiency virus/acquired immune deficiency syndrome patients: report of eight cases. Journal of Oral & Maxillofacial Surgery, 54(9), 1056-1060.
72. Chidzonga, M. M., & Mahomva, L. (2008). Noma (cancrum oris) in human immunodeficiency virus infection and acquired immunodeficiency syndrome (HIV and AIDS): clinical experience in Zimbabwe. Journal of Oral & Maxillofacial Surgery, 66(3), 475-485.
73. Chidzonga, M. M., & Mahomva, L. (2008). Recurrent noma (cancrum oris) in human immunodeficiency virus infection and acquired immunodeficiency syndrome (HIV and AIDS): report of a case. Journal of Oral & Maxillofacial Surgery, 66(8), 1726-1730.
74. Chlorate of Potash in Ulcerative Stomatitis and Cancrum Oris. (1853). The Buffalo medical journal and monthly review of medical and surgical science, 9(5), 303.
75. Chung, H.-l., Chang, A., Chang, N.-c. u., Feng, S.-l., Lu, J.-p., & Tsao, Y.-p. (1949). The Efficacy of Penicillin in the Treatment of Noma. occurring in Kala-Azar Patients. Chinese Medical Journal, 67(9), 469-473.
76. Clow, J. M. (1941). Shensi Province as an Endemic Focus of Kala-Azar. A Preliminary Report. Chinese Medical Journal, 59(2), 150-155.
77. Clow, J. M. (1943). Kala-Azar in Shensi Province. Chinese Medical Journal, 61(4), 281-290.
78. Cocquempot, K., Javaudin, O., Lerasle, P., & Aigle, L. (2014). Noma in a 4 year-old girl: a case report from Chad. [French]. [Noma chez une enfant : une observation au Tchad.]. Medecine et Sante Tropicales, 24(1), 99-104.
79. Coupe, M. H., Johnson, D., Seigne, P., & Hamlin, B. (2013). Special article: airway management in reconstructive surgery for noma (cancrum oris). Anesthesia & Analgesia, 117(1), 211-218.
80. Cousins, J. W. (1893). Cancrum Oris, Followed by Extensive Ulceration of the Cheek and Ankylosis of the Jaw: Recovery. British Medical Journal, 1(1684), 739-740.
81. Coward, T. (1839). CANCRUM ORIS NOT THE PRODUCT OF MERCURY. The Lancet, 33(849), 410.
82. Cressy, A. Z. (1901). "IX. Cancrum Oris Successfully treated by Excision of the Cautery." Annals of Surgery 34(2): 288.282-290.
83. Cribier, B. (2008). "Presentation to the French Dermatology Society in 1908: Two cases of noma in Algeria. [French]." Annales De Dermatologie Et De Venereologie 135(11): 802-803.
84. D'Agostino, F. J. (1951). A review of noma; report of a case treated with aureomycin. Oral Surgery, Oral Medicine, Oral Pathology, 4(8), 1000-1006.
85. Daya, M., & Nair, V. (2009). Free radial forearm flap lip reconstruction: a clinical series and case reports of technical refinements. Annals of Plastic Surgery, 62(4), 361-367.
86. Daya, M., & Pillay, T. (2019). Head and neck microsurgical reconstruction using the superficial temporal vein for antegrade and retrograde drainage: A clinical case series. European Journal of Plastic Surgery.
87. Dean, J. A., & Magee, W. (1997). One-stage reconstruction for defects caused by cancrum oris (noma). Annals of Plastic Surgery, 38(1), 29-35.
88. Deeb, G. R., Yih, W. Y., Merrill, R. G., & Lundeen, R. C. (1999). Noma: report of a case resulting in bony ankylosis of the maxilla and mandible. Dento-Maxillo-Facial Radiology, 28(6), 378-382.
89. DEFORMITY FROM CANCRUM ORIS. (1857). The Lancet, 70 (1778), 325.
90. Del Bene, M., Amadei, F., Petrolati, M., Rovati, L. V., Confalonieri, P. L., & Caronni, E. P. (1999). Radial forearm fasciocutaneous free flap as a solution in case of Noma. Microsurgery, 19(1), 3-6.
91. Denloye, O. O., Aderinokun, G. A., Lawoyin, J. O., & Bankole, O. O. (2003). Reviewing trends in the incidence of cancrum oris in Ibadan, Nigeria. West African Journal of Medicine, 22(1), 26-29.
92. Dhanrajani, A., & Khubchandani, R. P. (2014). Acute gangrenous gingivitis - Early stage noma. Indian Pediatrics, 51(4), 333.
93. Diamond, J. S. (1935). A case of noma (cancrum oris) complicating non-specific ulcerative colitis. The American Journal of Digestive Diseases, 2(11), 698-706.
94. Dijkstra, R., Abate-Green, C., & Yoo, M. C. (1986). Noma. European Journal of Plastic Surgery, 9(2), 46-51.
95. Ding, X., et al., (2020). Case Report: Malignant Transformation of Noma: Repair by Forearm Flap. The American journal of tropical medicine and hygiene, 103(4), 1697–1699.
96. Dinh Van Truong, D. D. S., & Tung, N. T. Rehabilitation of Severe Maxillary Noma Defect with Zygomatic Implants: A Case Report. ATTENTION PROSPECTIVE, 16.
97. Diombana, M. L., Kussner, H., Soumare, S., Doumbo, O., & Penneau, M. (1998). Nomas dans le service de stomatologie de l'hopital national de Kati (République du Mali) : 22 cas
98. Diop, E. M. and L. S. Diop (1980). "Another exceptional form of noma, in the velum. [French]." Dakar Medical 24(1): 85-89.
99. Diop, L. S., Medji, L. A., Diop, E. H. M., Tending, G., & Agbalika, F. (1976). A contribution to the study of the clinical and therapeutic aspects of developing noma. [Contribution a l'etude clinique et therapeutique du noma evolutif.]. Medecine d'Afrique Noire, 23(8 /9), 533-539.
100. Diop, L., et al.,(1971). ""[An exceptional form of noma]."" Bulletin de la Societe Medicale d’Afrique Noire de Langue Francaise 16(4): 599-600.
101. Diop, L., Medji, L. A., Astabie, F., & Aihonnou, H. (1972). [Bilateral, evolutive, nonmutilating noma (a case)]. Bulletin de la Societe Medicale d’Afrique Noire de Langue Francaise, 17(4), 511-514.
102. Donald, J. (1892). NOMA. AS A COMPLICATION OF ENTERIC FEVER. The Lancet, 139(3573), 417.
103. Duke B. F. (1881). Cancrum Oris. The Southern medical record, 11(12), 451.
104. Duncan, J. F. (1852). On the administration of mercury in cases of cancrum oris. The Dublin Quarterly Journal of Medical Science, 14(2), 265-273.
105. Dunn, R. (1843). FATAL CASE OF CANCRUM ORIS, IN WHICH AN INQUEST WAS HELD ON THE BODY. The Lancet, 41(1050), 60-62.
106. Durrani, K. M. (1973). Surgical repair of defects from noma (cancrum oris). Plastic & Reconstructive Surgery, 52(6), 629-634.
107. Dutasta, B. (1987). 8 cases of African noma. The therapeutic importance of nutrition. [French]. [Huit cas de noma africain. Importance therapeutique de l'alimentation.]. Revue de Stomatologie et de Chirurgie Maxillo-Faciale, 88(2), 139-142.
108. Eckstein, A. (1940). NOMA. Archives of Pediatrics & Adolescent Medicine, 59(2), 219–237.
109. Eipe, N., Neuhoefer, E. S., La Rosee, G., Choudhrie, R., Samman, N., & Kreusch, T. (2005). Submental intubation for cancrum oris: a case report. Paediatric Anaesthesia, 15(11), 1009-1012.
110. Elazary, A. S., Lador, N., Shauer, A., Meir, K., Pollak, A., Wolf, D. G., . . . Mevorach, D. (2004). Tongue necrosis and pericarditis. Lancet, 363(9413), 948.
111. Elder, G. (1893). Case of Cancrum Oris. Transactions of the Medicochirurgical Society of Edinburgh, 12, 259-262.
112. Ellouz, M., Adouani, A., & Seghir, M. (1989). Case report of an African disease: Cancrum oris. [French]. [A propos d'une maladie africaine: Le noma.]. Annales de Chirurgie Plastique et Esthetique, 34(4), 334-338.
113. Ellouz, M., et al., (1989). Case report of an african disease: cancrum oris. Annales de chirurgie plastique esthetique, 34(4), 334-338.
114. Emeršič, N., et al., (2021). Case Report: Necrotizing Stomatitis as a Manifestation of COVID-19-Associated Vasculopathy. Frontiers in pediatrics, 9, 800576.
115. Enwonwu, C. O. (1972). Epidemiological and biochemical studies of necrotizing ulcerative gingivitis and noma (cancrum oris) in Nigerian children. Archives of Oral Biology, 17(9), 1357-1371.
116. Enwonwu, C. O. (2006). Noma - The ulcer of extreme poverty. New England Journal of Medicine, 354(3), 221-224.
117. Enwonwu, C. O., Falkler Jr, W. A., Idigbe, E. O., Afolabi, B. M., Ibrahim, M., Onwujekwe, D., . . . Meeks, V. I. (1999). Pathogenesis of cancrum oris (noma): Confounding interactions of malnutrition with infection. American Journal of Tropical Medicine and Hygiene, 60(2), 223-232.
118. Enwonwu, C. O., Phillips, R. S., & Ferrell, C. D. (2005). Temporal relationship between the occurrence of fresh noma and the timing of linear growth retardation in Nigerian children. Tropical Medicine & International Health, 10(1), 65-73.
119. Evans, L. M., Lane, H., & Jones, M. K. (2001). Cancrum oris in a Caucasian male with Type 2 diabetes mellitus. Diabetic Medicine, 18(3), 246-248.
120. Fackler, M. L. (1971). Reconstruction surgery in Vietnam. U. S. Navy medicine, 58(4), 12-17.
121. Falkler Jr, W. A., Enwonwu, C. O., & Idigbe, E. O. (1999). Isolation of Fusobacterium necrophorum from cancrum oris (noma). American Journal of Tropical Medicine and Hygiene, 60(1), 150-156.
122. Falkler Jr, W. A., Enwonwu, C. O., & Idigbe, E. O. (1999). Microbiological understandings and mysteries of noma (cancrum oris). Oral Diseases, 5(2), 150-155.
123. Fan, P. L., & Scott, A. V. (1934). A Study of Noma complicating Kala Azar in Children. Chinese Medical Journal, 48(10), 1046-1057.
124. Farina, R., Neto, D. S., Santos, S. P., & Castro, O. (1955). Surgical treatment of sequelae of gangrenous stomatitis (noma). Plastic & Reconstructive Surgery, 16(1), 47-56.
125. Farley, E. S., et al., (2020). Outcomes at 18 mo of 37 noma (cancrum oris) cases surgically treated at the Noma Children's Hospital, Sokoto, Nigeria. Transactions of the Royal Society of Tropical Medicine and Hygiene, 114(11), 812–819.
126. Farley, E., et al., (2018). Risk factors for diagnosed noma in northwest Nigeria: A case-control study, 2017. PLoS neglected tropical diseases, 12(8), e0006631.
127. Farley, E., et al., (2020). The prevalence of noma in northwest Nigeria. BMJ global health, 5(4), e002141.
128. Farley, E., Lenglet, A., Ariti, C., Jiya, N. M., Adetunji, A. S., van der Kam, S., & Bil, K. (2018). Risk factors for diagnosed noma in northwest Nigeria: A case-control study, 2017. PLoS Neglected Tropical Diseases [electronic resource], 12(8), e0006631.
129. Fatahzadeh, M. (2018). The dentist's role in the prevention and management of necrotizing stomatitis in the immunosuppressed. Quintessence International, 49(5), 399-405.
130. Faye, O., Keita, M., N'Diaye H, T., Konare, H. D., Darie, H., Keita, S., & Mahe, A. (2003). [Noma in HIV-infected adults]. Annales De Dermatologie Et De Venereologie, 130(2 Pt 1), 199-201.
131. Faye, O., Keita, M., N'diaye, H. T., Konare, H. D., Darie, H., Keita, S., & Mahe, A. (2003). Noma chez des adultes infectés par le VIH [Noma in HIV-infected adults]. Annales de dermatologie et de venereologie, 130(2 Pt 1), 199–201.
132. Feller, L., et al., (2012). Oral ulcers and necrotizing gingivitis in relation to HIV-associated neutropenia: a review and an illustrative case. AIDS research and human retroviruses, 28(4), 346-351.
133. Fetterolf, B., Zabella, A., & Strote, J. (2015). Oral Ulcerations. Western Journal of Emergency Medicine: Integrating Emergency Care with Population Health, 16(7).
134. Fieger, A., Marck, K. W., Busch, R., & Schmidt, A. (2003). An estimation of the incidence of noma in north-west Nigeria. Tropical Medicine & International Health, 8(5), 402-407.
135. Fleming, R. A. (1893). A Case of Cancrum Oris, with a Short Critique on the Disease. Transactions of the Medicochirurgical Society of Edinburgh, 12, 251-259.
136. Foussadier, F., & Servant, J. M. (2004). Activity report about hospital Saint-Louis's team for taking in charge sequellae of Noma. [French]. [Bilan d'activite des equipes de l'hopital Saint-Louis a Niamey pour la prise en charge des sequelles de noma (medecins du Monde-Operation sourire).]. Annales de Chirurgie Plastique et Esthetique, 49(4), 345-354.
137. Francois, P., Whiteson, K., Baratti-Mayer, D., Schrenzel, J., & Pittet, D. (2015). Noma disease: 10 years of research in the quest of a microbial etiology. Antimicrobial Resistance and Infection Control. Conference: 3rd International Conference on Prevention and Infection Control, ICPIC, 4(SUPPL. 1).
138. Gayet-Ageron, A., Baratti-Mayer, D., Courvoisier, D., & Pittet, D. (2011). Is noma an infectious disease? Is it transmissible? BMC Proceedings. Conference: International Conference on Prevention and Infection Control, ICPIC, 5(SUPPL. 6).
139. Gebretsadik, H. G., & de Kiev, L. C. (2022). A retrospective clinical, multi-center cross-sectional study to assess the severity and sequela of Noma/Cancrum oris in Ethiopia. PLoS neglected tropical diseases, 16(9), e0010372.
140. Ghorui, T., & Ray, A. (2018). Release of Extra Articular Ankylosis of Jaws as a Sequelae of Cancrum Oris with Extensive Gingival Myasis in a Scoliosis Patient: A Rare Case Report. Indian Journal of Otolaryngology and Head and Neck Surgery.
141. Ghose, A. K. (1937). A Case of Cancrum Oris as a Complication of Bacillary Dysentery. Indian Medical Gazette, 72(7), 419-420.
142. Ghosh, K. P. (1947). Use of penicillin in cancrum oris. The Indian Journal of Pediatrics, 14(4), 171-172.
143. Giessler, G. A., Cornelius, C. P., Suominen, S., Borsche, A., Fieger, A. J., Schmidt, A. B., & Fischer, H. (2007). Primary and secondary procedures in functional and aesthetic reconstruction of noma-associated complex central facial defects. Plastic & Reconstructive Surgery, 120(1), 134-143.
144. Giessler, G. A., Fieger, A., Cornelius, C. P., & Schmidt, A. B. (2005). Microsurgical reconstruction of noma-related facial defects with folded free flaps - An overview of 31 cases. Annals of Plastic Surgery, 55(2), 132-138.
145. Giessler, G. A., Schmidt, A. B., & Marck, K. W. (2003). Noma: Experiences with a microvascular approach under West African conditions. Discussion. Plastic and reconstructive surgery (1963), 112(4), 947-956.
146. Giessler, G. A., Schmidt, A. B., Deubel, U., & Cornelius, C. P. (2004). Free flap transfer for closure and interposition-arthroplasty in noma defects of the lateral face associated with bony ankylosis. Journal of Craniofacial Surgery, 15(5), 766-772; discussion 773.
147. Giessler,G.A., Borsche, A., Lim, P.K., Schmidt, A.B., Cornelius C.P. First Experiences with Simultaneous Skeletal and Soft Tissue Reconstruction of Noma-Related Facial Defects. J Reconstr Microsurg. 2012 Feb;28(2):85-94.
148. Ginisty, D., Piral, T., Adamsbaum, C., Camara, A., & Rak-Merkin, H. (1996). Permanent constriction of the jaws in children: 3 cases with extra-articular etiology. [French]. [Les constrictions permanentes des machoires de l'enfant. Trois cas d'etiologie extra-articulaire.]. Revue de Stomatologie et de Chirurgie Maxillo-Faciale, 97(1), 47-52.
149. Gollogly, J. G., & Mussomeli, I. (2007). Noma in Cambodia: scars from the past. Asian Biomedicine, 1(4), 377-381.
150. Gould, F. (1872). A CASE OF BRONCHO-PNEUMONIA,. FOLLOWED BY CANCRUM ORIS AND SUBSEQUENT NECROSIS OF JAW ; RECOVERY. The Lancet, 100(2553), 150.
151. Gray, J. (1856). Cancrum Oris. Glasgow Medical Journal, 4(13), 16-21.
152. Griffin, J. M., Bach, D. E., Nespeca, J. A., & Marshall, K. J. (1983). Noma. Report of two cases. Oral Surgery Oral Medicine and Oral Pathology, 56(6), 605-607.
153. Grozdeva, R. S., et al., (2020). First Case of Cancrum Oris (Noma) in an HIV-Positive Patient in Bulgaria. Acta Medica Bulgarica, 47(3), 34–37.
154. Guillozet, N. (1981). Oral stenosis in measles. International Journal of Dermatology, 20(4), 262-263.
155. Gupta, A., Sardana, K., & Gautam, R. K. (2018). An unusual case of noma caused by Klebsiella pnuemoniae and its management. Tropical Doctor, 48(3), 230-232.
156. Hardman F. G. (1973). A case of cancrum oris associated with agranulocytosis. Transactions of the International Conference on Oral Surgery, 4, 349–350.
157. Hartman, E. H., et al., (2006). The use of the pedicled supraclavicular flap in noma reconstructive surgery. Journal of plastic, reconstructive & aesthetic surgery, 59(4), 337-342.
158. Hartman, E. H., Van Damme, P. A., Rayatt, S., & Kuokkanen, H. O. (2010). Return of the waltzing flap in noma reconstructive surgery: revisiting the past in difficult circumstances. Journal of Plastic, Reconstructive & Aesthetic Surgery: JPRAS, 63(1), e80-81.
159. Hatcher, J., & Williamson, L. (2017). Noma in a patient with HIV. Lancet Infectious Diseases, 17(6), 672-672.
160. Hebting, J. M. (2016). Decision physiotherapy workshop: Issouf C. After-effects of cancrum-oris (noma). [Decision kinesitherapique : Issouf C. Sequelles de noma.]. Kinesitherapie, 16(176-177), 79-84.
161. Hefferman, L. W. (1923). Cancrum Oris Treated by Excision and Subsequent Tube Grafting. Indian Medical Gazette, 58(9), 434-437.
162. Heitland, A. S., & Pallua, N. (2005). The single and double-folded supraclavicular island flap as a new therapy option in the treatment of large facial defects in noma patients. Plastic & Reconstructive Surgery, 115(6), 1591-1596.
163. Hicken, N. (1935). Infectious gangrene of the skin due to bacterial synergism. Archives of Surgery, 31(2), 253.
164. Hill, A. J. (1978). Release of mandibular ankylosis due to gross tissue loss in the cheek. International Journal of Oral Surgery, 7(4), 369-373.
165. Hitzelberger, A. C. (1940). Case report no. 36 Noma of the cheek. American Journal of Orthodontics and Oral Surgery, 26(3), 290-292.
166. Holle, J. (2009). Lockjaw treatment after noma in the third world. Journal of Craniofacial Surgery, 20(8 SUPPL. 2), 1910-1912.
167. Holle, J., et al., (2020). Distraction Therapy to Correct Trismus Following Noma. The Journal of craniofacial surgery, 31(2), 488–491.
168. Honeyman, C., et al., (2020). Long-term outcomes associated with short-term surgical missions treating complex head and neck disfigurement in Ethiopia: A retrospective cohort study. Journal of plastic, reconstructive & aesthetic surgery : JPRAS, 73(5), 951–958.
169. Horsley, J. S. (1922). Plastic operations for defects due to noma. Southern Medical Journal, 15(7), 557-561.
170. Hu, J., Kent, P., Lennon, J. M., & Logan, L. K. (2015). Acute necrotising ulcerative gingivitis in an immunocompromised young adult. BMJ case reports, bcr2015211092.
171. Huijing, M. A., Marck, K. W., Combes, J., Mizen, K. D., Fourie, L., Demisse, Y., . . . McGurk, M. (2011). Facial reconstruction in the developing world: A complicated matter. British Journal of Oral and Maxillofacial Surgery, 49(4), 292-296.
172. Hunt, H. (1843). Remarks on cancrum oris and the gangrenous erosion of the cheek of Mr. Dease and Dr. Underwood, and more particularly on the efficacy of the chlorate of potash in the treatment of those diseases. Medicochirurgical Transactions, 26, 142-153.
173. Huyghe, A., Francois, P., Mombelli, A., Tangomo, M., Girard, M., Baratti-Mayer, D., . . . Schrenzel, J. (2013). Microarray Analysis of Microbiota of Gingival Lesions in Noma Patients. PLoS Neglected Tropical Diseases, 7 (9) (no pagination)(e2453).
174. Ibeziako, S. N., Nwolisa, C. E., & Nwaiwu, O. (2003). Cancrum oris and acute necrotising gingivitis complicating HIV infection in children. Annals of Tropical Paediatrics, 23(3), 225-226.
175. Idigbe, E. O., Enwonwu, C. O., Falkler, W. A., Ibrahim, M. M., Onwujekwe, D., Afolabi, B. M., . . . Meeks, V. I. (1999). Living conditions of children at risk for noma: Nigerian experience. Oral Diseases, 5(2), 156-162.
176. Ingals, E. F. (1872). A Case of Cancrum Oris. The American Journal of Dental Science, 6(4), 182-183.
177. Isah, S., et al., (2021). Model of care, Noma Children's Hospital, northwest Nigeria. Tropical medicine & international health : TM & IH, 26(9), 1088–1097.
178. Jain, M., Sarkar, N., & Lamba, P. A. (1985). Noma--a case report. Indian Journal of Ophthalmology, 33(4), 249-250.
179. Jayasankar, P. P., Chavda, J., & Shah, R. M. (2004). Oro-nasal fistula of infective origin and alveolar necrosis in a 13 year old boy. Indian Journal of Dental Research, 15(3), 107-109.
180. Jelliffe, D. B. (1952). Infective gangrene of the mouth (cancrum oris). Pediatrics, 9(5:1), 544-550.
181. Jimenez, M., & Baer, P. N. (1975). Necrotizing ulcerative gingivitis in children: a 9 year clinical study. Journal of Periodontology, 46(12), 715-720.
182. Jin, L. Y., Ou, X. R., He, Z. J., & Xie, X. L. (2010). Maxillofacial deformity caused by cancrum oris: a case report. [Chinese]. Hua xi kou qiang yi xue za zhi = Huaxi kouqiang yixue zazhi = West China journal of stomatology, 28(3), 342-344.
183. Kaba, M., & Traore, M. (2002). Noma: apropos of a case in a 9-year-old child at the Centre Hospitalier de Libreville (Gabon). [French]. [Le noma: a propos d'un cas chez un enfant de 9 ans au Centre Hospitalier de Libreville (Gabon).]. Odonto-stomatologie tropicale = Tropical dental journal, 25(99), 26-28..
184. Kaimenyi, J. T., & Guthua, S. W. (1994). Residual facial deformity resulting from cancrum oris: a case report. East African Medical Journal, 71(7), 476-478.
185. Kato, H., & Imamura, A. (2017). Unexpected acute necrotizing ulcerative gingivitis in a well-controlled hiv-infected case. Internal Medicine, 56(16), 2223-2227.
186. Katz, E., Engelhard, D., Kerem, E., Eid, A., & Berlatzki, Y. (1987). Noma in an unusual case of perforated appendicitis with pneumoperitoneum. Journal of Pediatric Surgery, 22(11), 1017-1018.
187. Keiller, A. (1862). On Cancrum Oris. Edinburgh Medical Journal, 7(10), 919-934.
188. Kikuchi, I., & Sakaguchi, E. (1989). Bacterial gangrene on the cheek of a comatose patient. Necrotizing fasciitis or noma. Journal of Dermatology, 16(3), 251-252.
189. King, H. D. (1911). Noma (cancrum oris) in an adult, with report of a case. Journal of the American Medical Association, LVI(20), 1449-1451.
190. Kingsford, E. C. (1891). CANCRUM ORIS. The Lancet, 138(3550), 607-608.
191. Kittredge, C. S. (1872). Cancrum Oris. The American Journal of Dental Science, 6(1), 22-27.
192. Kobus, K. (2003). Total lower lip reconstruction. European Journal of Plastic Surgery, 25(7-8), 357-361.
193. Koech, K. J. (2010). Cancrum oris in an adult with human immunodeficiency virus infection: case report. East African Medical Journal, 87(1), 38-40.
194. Konan K.E., et al., (2008). L’antisepsie dans la prise en charge du Noma: Role Du Dakin Cooper Stabalise. Medecine d’Afrique Noire, 55(5), 253-257.
195. Konan, K. E., Anzouan, K. E., N'Guessan, N. D., Assouan, C., & Assa, A. (2008). Antiseptics for control of noma: the role of Dakin Cooper StabiliseReg.. [French]. [L'antisepsie dans la prise en charge du noma: role du dakin cooper stabiliseReg..]. Medecine d'Afrique Noire, 55(5), 253-257.
196. Konsem, T., Millogo, M., Assouan, C., & Ouedraogo, D. (2014). [Evoluting form of cancrum oris, about 55 cases collected at the Academic Hospital Yalgado Ouedraogo of Ouagadougou]. Bulletin de la Societe de Pathologie Exotique, 107(2), 74-78.
197. Konsem, T., Millogo, M., Gare, J., Ouedraogo, D., & Ouoba, K. (2014). Noma and Burkitt disease; A particular association about three observations seen in the Teaching Hospital Center Yalgado Ouedraogo (Burkina Faso). Bulletin de la Societe de Pathologie Exotique, 107(3), 146-150.
198. Kovačić, M., Kovačić, I., & Veršić, M. (2021). Necrotic ulcerative stomatitis in a patient with long-standing celiac disease: a case report. Croatian medical journal, 62(5), 518–522.
199. Kuhnel, T. S., Dammer, R., Dunzl, B., Beule, A. G., & Strutz, J. (2003). New split scar cheek flap in reconstruction of noma sequelae. British Journal of Plastic Surgery, 56(6), 528-533.
200. Kumar, M., Sen, S., Zachariah, N., Thomas, R. J., & Mammen, K. E. (1998). Recurrent parotid abscess formation 8 years after an episode of cancrum oris. Pediatric Surgery International, 13(2-3), 162.
201. Lacour, M., & Castets, M. (1968). A case of noma treated with metronidazole. [Sur un cas de noma traite par le metronidazole.]. Bulletin de la Societe Medicale d'Afrique Noire de Langue Francaise, 13(3), 701-703.
202. Lagrot, F., Py, N., Greco, J., & Lavergne, E. (1961). [Noma and constriction of the jaws]. Annales de Chirurgie Plastique, 6, 243-246.
203. Lagundoye, S. B., & Oluwasanmi, J. O. (1975). Radiologic examination of trismus as a complication of cancrum oris. Oral Surgery, Oral Medicine, Oral Pathology, 39(5), 812-820.
204. Lamichhane, S., et al., (2020). Necrotizing Stomatitis in Varicella Zoster Infection. Kathmandu University medical journal (KUMJ), 18(70), 210–213.
205. Laslett, E. E. (1902). A CASE OF CANCRUM ORIS AFFECTING BOTH SIDES. The Lancet, 159(4095), 513-514.
206. Lathigara, M. D. (1935). An Unusual Case of Cancrum Oris. Indian Medical Gazette, 70(11), 626-627.
207. Layman, P. R. (1983). Bypassing a problem airway. Anaesthesia, 38(5), 478-480.
208. Lazarus, D., & Hudson, D. A. (1997). Cancrum oris - A 35-year retrospective study. South African Medical Journal, 87(10), 1379-1382.
209. LE NOMA DE L’ENFANT EN MILIEU HOSPITALIER DE BOBO-DIOULASSO : ASPECTS EPIDEMIOLOGIQUES, CLINIQUES ET PRISE EN CHARGE
210. Le, B. (1927). Noma Consecutive on Framboesial Lesions of the Lips. [Une observation de Noma consecutif a des lesions pianiques des le vres.]. Annales de Medecine et de Pharmacie Coloniales, 25(1), 127-129.
211. Lebon, P. (1958). [Treatment of a noma during the development of myeloblastic leukemia]. Algerie Medicale, 62(3), 347-348 passim.
212. Lee,Y.J.(2017).Nomalike Necrotizing Stomatitis in a child with Corhn's disease.
213. Lembo, S., De Leonibus, C., Francia, M. G., Lembo, C., & Ayala, F. (2011). Cancrum oris in a boy with Down syndrome. Journal of the American Academy of Dermatology, 64(6), 1200-1202.
214. Lerman, R. L., & Grodin, M. A. (1977). Necrotizing stomatitis in a pediatric burn victim. Journal of Dentistry for Children, 44(5), 388-390.
215. Levison, J. L. (1851). Cases of Cancrum Oris. The American Journal of Dental Science, 2(1), 107-113.
216. Li, M., Jiang, H. B., Tao, Z. J., & Wu, Y. N. (2015). Extended Free Forearm Flap for Treatment of Temporomandibular Joint Pseudoankylosis. Journal of Craniofacial Surgery, 26(4), e358-359.
217. Lieberman, J., Lynfield, Y. L., & Rosen, P. (1987). Noma. Cutis, 39(6), 501-502.
218. Linenberg, W. B., Schmitt, J., & Harpole, H. J. (1961). Noma. Report of a case. Oral Surgery, Oral Medicine, Oral Pathology, 14, 1138-1141.
219. Lopez, R., et al., (2002). Epidemiology of necrotizing ulcerative gingival lesions in adolescents. Journal of periodontal research, 37(6), 439–444.
220. Lourmet, J., et al.,(1969). "A case of permanent constriction of the jaws." Bulletin de la Societe medicale d’Afrique noire de langue francaise 14(4): 757-759.
221. Lu, X. G., Cai, Z. G., Zhang, Y., & Sun, Y. G. (2010). One-stage operation for noma-induced bilateral ankylosis accompanied with mouth verrucous carcinoma - a case report and review of literature. Chinese Journal of Dental Research, 13(1), 67-69.
222. Lubala, T. K., Mutombo, A. M., Mukuku, K. O., Ilunga, M. P., & Shongoya, M. P. (2012). Association of acute noma, HIV and severe malnutrition in children: about 2 cases. [French]. [Association noma aigu - VIH - malnutrition severe chez l'enfant: a propos de 2 cas.]. Pan African Medical Journal, 13(1).
223. Maccarrone, F., & Alicandri‐Ciufelli, M. (2019). Plaut‐Vincent’s Ulcerative Gingivitis and Tonsillitis. Otolaryngology–Head and Neck Surgery, 161(6), 1056-1057.
224. Mackay, D. (1949). Cancrum oris among African natives. British Medical Journal, 1(4596), 223.
225. Madabhavi I, Revannasiddaiah S, Sarkar M. Cancrum oris (noma): An early sign of acute lymphoblastic leukemia relapse. Indian J Dermatol Venereol Leprol 2018;84:373)
226. Madabhavi, I., Revannasiddaiah, S., & Sarkar, M. (2018). Cancrum oris (noma): An early sign of acute lymphoblastic leukemia relapse. Indian journal of dermatology, venereology and leprology, 84(3), 373.
227. Madsen, T., Medina, C., Jespersen, S., Wejse, C., & Honge, B. L. (2017). Noma in an HIV infected patient in Guinea-Bissau: a case report. Infection, 45(6), 897-901.
228. Magan-Fernandez, A., et al., (2015). Two cases of an atypical presentation of necrotizing stomatitis. Journal of Periodontal & Implant Science, 45(6), 252-256.
229. Majumder, D. C. (1938). A Case Note on Cancrum Oris Following Pneumonia Treated by Prontosil. Indian Medical Gazette, 73(10), 614.
230. Malberger, E. (1967). Acute infectious oral necrosis among young children in the Gambia, West-Africa. Journal of Periodontal Research, 2(2), 154-162.
231. Malden, N. (1985). An interesting case of adult facial gangrene (from Papua, New Guinea). Oral Surgery, Oral Medicine, Oral Pathology, 59(3), 279-281.
232. Malek, R., Gharibi, A., Khlil, N., & Kissa, J. (2017). Necrotizing ulcerative gingivitis. Contemporary clinical dentistry, 8(3), 496.
233. Maley, A., Desai, M., & Parker, S. (2015). Noma: A disease of poverty presenting at an urban hospital in the United States. JAAD Case Reports, 1(1), 18-20.
234. Mamadou, S., Kaka, M., Montavon, C., Noman, Y., Maty, M., Delaporte, E., & Mboup, S. (2002). About an association between HIV and noma in Niger (short report). Bulletin de la Societe de Pathologie Exotique, 95(2), 76-77.
235. Mangundjaja, S., & Arumsari, A. (2002). Correction of lip defect caused by noma. Asian Journal of Oral and Maxillofacial Surgery, 14(4), 245-249.
236. Marck, K. W., de Bruijn, H. P., Schmid, F., Meixner, J., van Wijhe, M., & van Poppelen, R. H. M. (1998). Noma: the Sokoto approach. European Journal of Plastic Surgery, 21(6), 277-280.
237. Marck, K. W., Van Der Lei, B., Spijkervet, F. K. L., Adeniyi, S. A., Meixner, J., & De Bruijn, H. P. (2000). The prefabricated superficial temporal fascia flap in noma surgery. European Journal of Plastic Surgery, 23(4), 188-191.
238. Marck, R., Huijing, M., Vest, D., Eshete, M., Marck, K., & McGurk, M. (2010). Early outcome of facial reconstructive surgery abroad: A comparative study. European Journal of Plastic Surgery, 33(4), 193-197.
239. Martos, J., et al., (2019). Clinical treatment of necrotizing ulcerative gingivitis: a case report with 10-year follow-up. General Dentistry, 67(3), 62-5.
240. Mascitti, M., et al., (2019). Noma: a reappraisal in Western countries - are HIV-negative immunocompetent adult patients safe?. Journal of biological regulators and homeostatic agents, 33(3), 957–961.
241. Masipa, J. N., Baloyi, A. M., Khammissa, R. A., Altini, M., Lemmer, J., & Feller, L. (2013). Noma (cancrum oris): a report of a case in a young AIDS patient with a review of the pathogenesis. Head and neck pathology, 7(2), 188-192.
242. McCurdy, S. L. (1907). Cancrum oris. Journal of the American Medical Association, XLIX(17), 1441.
243. Mcdonald W.P. (1891). Report of Case of Cancrum Oris. The dental registrar, 45(4),161-163.
244. Mehrotra, M. C. (1966). Cancrum oris. A clinical survey of 20 cases. Journal of the Indian Medical Association, 47(11), 555-557.
245. Mehta, D. T. (1946). Cases of cancrum oris. The Antiseptic, 43, 200.
246. Milbourne West, R. (1896). TWO CASES OF GANGRENOUS STOMATITIS, ONE ENDING FATALLY. The Lancet, 147(3785), 704-705.
247. Millogo, M., Konsem, T., Ouedraogo, D., Ouoba, K., & Zwetyenga, N. (2012). HIV and noma in Burkina Faso. Revue de Stomatologie et de Chirurgie Maxillo-Faciale, 113(6), 433-436.
248. Mishra S., et al., (2020). Lip-Nose Reconstruction Post-Cancrum Oris Defect: A Case Report. 14(4), 8700-8704.
249. Mishra, Y. C., & Manjula, D. (1986). Gangrenous stomatitis--a case report. Journal of the Indian Dental Association, 58(11), 465-468.
250. Montandon, D., Lehmann, C., & Chami, N. (1991). The surgical treatment of noma. Plastic & Reconstructive Surgery, 87(1), 76-86.
251. Moore, B. S., & Clifford, J. S. (1912). Radical Surgery in Cases of Cancrum-Oris or "Noma", with Report of a Case of Bi-Lateral Infection Complicating Typhoid Fever with Recovery. Atlanta journal-record of medicine, 59(6), 287–290.
252. Mujtaba, B., et al., (2022). Clinico-Pathological Analysis of Osteomyelitis in Cancrum Oris (Noma) Patients Seen in Noma Children Hospital, Northwest Nigeria. Nigerian Journal of Dental Research, 7(1), 29–34.
253. Mukerji, S. N. (1927). A Case of Stiff Jaw after Cancrum Oris-Surgical Interference-Cure. Indian Medical Gazette, 62(7), 387.
254. Murray, R. W. (1889). CANCRUM ORIS. The Lancet, 133(3428), 959.
255. Murrell, G. L. (2010). Trench mouth. Otolaryngology—Head and Neck Surgery, 143(4), 599-600.
256. Muzyka, B. C., & Glick, M. (1994). HIV infection and necrotizing stomatitis. General Dentistry, 42(1), 66-68.
257. Naidoo, S., & Chikte, U. M. (2000). Noma (cancrum oris): case report in a 4-year-old HIV-positive South African child. SADJ, 55(12), 683-686.
258. Nash, E. S., Cheng, L. H., & Smart, K. (1991). Cancrum oris-like lesions. British Journal of Oral & Maxillofacial Surgery, 29(1), 51-53.
259. Nath, S., & Jovic, G. (1998). Cancrum oris: management, incidence, and implications of human immunodeficiency virus in Zambia. Plastic & Reconstructive Surgery, 102(2), 350-357.
260. Nath, S., & Jovic, G. (1998). Total loss of upper and lower lips: challenges in reconstruction. British Journal of Oral & Maxillofacial Surgery, 36(6), 460-461.
261. Ndiaye, C. F., Bourgeois, D., Leclercq, M. H., & Berthe, O. (1999). Noma: Public health problem in Senegal and epidemilogical surveillance. Oral Diseases, 5(2), 163-166.
262. Noever, G., & Feurer, R. (2000). The free radial forearm flap for reconstruction of facial defects in noma patients. European Journal of Plastic Surgery, 23(1), 1-6.
263. NOMA ET INFECTION A VIH : A PROPOS D’UNE OBSERVATION AU CENTRE HOSPITALIER NATIONAL DE BOBO-DIOULASSO (BURKINA FASO)
264. Nthumba, P., & Carter, L. (2009). Visor flap for total upper and lower lip reconstruction: a case report. Journal of Medical Case Reports [Electronic Resource], 3, 7312.
265. Nwoku, A. L., & Sawyer, D. R. (1980). Cancrum oris mortalityin Lagos, Nigeria. Virginia Dental Journal, 57(2), 22-25.
266. O’Brien, N. M. (2019). Noma (cancrum oris) in a child with severe acute malnutrition in northern Nigeria. Arch Dis Child 2019;104(Suppl 2): A125.
267. Oberoi, G. S., Bithal, P. K., Saxena, N., & Kaul, H. L. (1985). Anesthesia in cancrum oris. Indian Journal of Pediatrics, 52(419), 671-672.
268. Obi, J. O. (1979). Measles in hospital practice in Nigeria. Journal of Tropical Pediatrics and Environmental Child Health, 25(1), 30-33.
269. Obiechina, A. E., Arotiba, J. T., & Fasola, A. O. (2000). Cancrum oris (noma): Level of education and occupation of parents of affected children in Nigeria. Odonto-stomatologie tropicale = Tropical dental journal, 23(90), 11-14.
270. O'Donoghue, J. J. (1931). A CASE OF CANCRUM ORIS. The Lancet, 218(5646), 1075.
271. Oginni, F. O., Oginni, A. O., Ugboko, V. I., & Otuyemi, O. D. (1999). A survey of cases of cancrum oris seen in Ile-Ife, Nigeria. International Journal of Paediatric Dentistry, 9(2), 75-80.
272. O'Gorman, P. W. (1882). Cancrum Oris in an European Soldier; Enlarged Spleen; Pancreatic Disease Post-Mortem Results. Indian Medical Gazette, 17(1), 14-15.
273. Oji, C. (2002). Cancrum oris: Its incidence and treatment in Enugu, Nigeria. British Journal of Oral and Maxillofacial Surgery, 40(5), 406-409.
274. Olasoji, H. O., Tahir, A., & Adesina, O. A. (2002). Noma in a Nigerian adult. Tropical Doctor, 32(3), 179-180.
275. Olsen, I.(2009).Cultivated and not-yet-cultivated bacteria in oral biofilms
276. Oluwasanmi, J. O., Lagundoye, S. B., & Akinyemi, O. O. (1976). Ankylosis of the mandible from cancrum oris. Plastic and Reconstructive Surgery, 57(3), 342-350.
277. Osuji, O. O. (1990). Necrotizing ulcerative gingivitis and cancrum oris (noma) in Ibadan, Nigeria. Journal of Periodontology, 61(12), 769-772.
278. Ouoba, K., Sanou, I., Dao, M., Kam, L., Ouedraogo, A., Ouedraogo, R., & Sawadogo, A. (1998). Progressive noma: apropos of 27 cases seen at the National Hospital Center of Ouagadougou. [French]. [Le noma evolutif: a propos de 27 cas observes au Centre Hospitalier National de Ouagadougou.]. Dakar Medical, 43(1), 45-48.
279. Owobu, T., et al., (2022). A review of noma cases in a tertiary hospital located in a conflict endemic region in Nigeria. Medicine, conflict, and survival, 38(4), 295–306.
280. Özberk, S. S., et al., (2018). Adjunct use of low-level laser therapy on the treatment of necrotizing ulcerative gingivitis: a case report. Journal of Lasers in Medical Sciences, 9(1), 73.
281. Palma-Perez, T. A. (1984). Noma in the 80's. JPMA. Journal of the Philippine Medical Association, 60(1), 12-14.
282. Palmer, B. B., Jr., & Carr, M. W. (1928). Medico-dental case records: Sixth report: A Clinico-Pathological Study. Journal of Dental Research, 8(5), 579-592.
283. Park, J., et al., (2022). Noma disease (cancrum oris, orofacial gangrene) in an acute myeloid leukemia patient: a case report. Journal of medical case reports, 16(1), 97.
284. Park, Y. R., et al., (2018). Restoration of an Upper Lip Affected by Necrotizing Ulcerative Stomatitis Using Bilateral Cheek Advancement with a Crescentic Perialar Excision. Archives of Aesthetic Plastic Surgery 24(2):87-90.
285. Paster, B. J., Falkler Jr, W. A., Jr., Enwonwu, C. O., Idigbe, E. O., Savage, K. O., Levanos, V. A., . . . Dewhirst, F. E. (2002). Prevalent bacterial species and novel phylotypes in advanced noma lesions. Journal of Clinical Microbiology, 40(6), 2187-2191.
286. Pedro, K., Smit, D. A., & Morkel, J. A. (2016). Cancrum Oris (noma) in an HIV-positive adult: a case report and literature review. South African Dental Journal, 71(6), 248-252.
287. Peri, G., et al.,(1978). "[Reconstruction of the lower lip for sequelae of noma (author's transl)]." Revue de Stomatologie et de Chirurgie Maxillo-Faciale 79(4): 347-348.
288. Phan Dinh, T. (1962). Noma in Viet-Nam. Indian Journal of Pediatrics, 29, 367-374.
289. Phillips, R. S., Enwonwu, C. O., & Falkler, W. A. (2005). Pro- versus anti-inflammatory cytokine profile in African children with acute oro-facial noma (cancrum oris, noma). European Cytokine Network, 16(1), 70-77.
290. Pittet, B., & Montandon, D. (1998). Nasal reconstruction in children: A review of 29 patients. Journal of Craniofacial Surgery, 9(6), 522-528.
291. Pittet, B., Jaquinet, A., & Montandon, D. (2001). Clinical experience in the treatment of noma sequelae. Journal of Craniofacial Surgery, 12(3), 273-283.
292. Pittet, B., Mahajan, A. L., Alizadeh, N., Schlaudraff, K. U., Fasel, J., & Montandon, D. (2006). The free serratus anterior flap and its cutaneous component for reconstruction of the face: a series of 27 cases. Plastic & Reconstructive Surgery, 117(4), 1277-1288.
293. Pons, R. (1925). Vaccinotherapy in Affections of Skin and Mucosa due to the Fuso-Spirochaetal Association. [Vaccinotherapie antispirillaire dans les affections a association fuso-spirochetienne de la peau et des muqueuses.]. Bulletin de la Societe de Pathologie Exotique, 18(5), 380-383.
294. Putri, I. L., Agustina, W., & Hutagalung, M. R. (2021). Columella reconstruction using double nasolabial flap and costal cartilage: A case report. Annals of medicine and surgery, 64, 102213.
295. Rajaleelan, W., & Otterman, M. (2020). Airway Management in Children with Noma Sequelae Undergoing Craniomaxillofacial Reconstructive Surgery.
296. Reynaud, J. (1950). Penicillin therapy of noma in Afghanistan. La semaine des hôpitaux : organe fondé par l'Association d'enseignement médical des hôpitaux de Paris, 26(35), 1640-1647.
297. Reynaud, J., et al.,(1965). "[On noma in children in Senegal (47 treated cases)]." Bulletin de la Societe Medicale d’Afrique Noire de Langue Francaise 10(3): 434-441.
298. Reynaud, J., Garand, G., Ployet, M. J., & Robier, A. (1978). [The pathogenesis of noma and Silvermann's syndrome: a subject for discussion (author's transl)]. Annales de Chirurgie Plastique, 23(4), 227-230.
299. Rickart, A. J., et al., (2020). Facing Africa: Describing Noma in Ethiopia. The American journal of tropical medicine and hygiene, 103(2), 613–618.
300. Rickart, A. J., Wilson, P., & Merrick, G. (2020). Single stage variation of myomucosal lip switch flap for secondary reconstruction of perioral defects. The British journal of oral & maxillofacial surgery, 58(9), 1205–1207.
301. Ritchie, R. P. (1872). Observations on the Inflammations of the Mouth in Children, with an Illustrative Case of Cancrum Oris. Transactions of the Edinburgh Obstetrical Society, 2, 418-435.
302. Rotbart, H. A., Levin, M. J., Jones, J. F., Hayward, A. R., Allan, J., McLane, M. F., & Essex, M. (1986). Noma in children with severe combined immunodeficiency. Journal of Pediatrics, 109(4), 596-600.
303. Rowe, D., McKerrow, N., Uys, A., & Winstanley, T. (2004). Cancrum oris (noma) in a malnourished HIV-positive child from rural Kwazulu-Natal. Southern African Journal of HIV Medicine(16), 45-46.
304. Royle, H. E. (1967). Use of an anticholinergic drug in the closure of an orofacial fistula due to noma. Report of a case. Oral Surgery, Oral Medicine, Oral Pathology, 24(1), 6-8.
305. Ruben, M. P., & Miller, M. (1964). Noma: Its Association with Nutritional Deprivation and Physical Debilitation. Report of a Case. Oral Surgery, Oral Medicine, Oral Pathology, 18, 167-175.
306. Ruegg, E. M., Baratti-Mayer, D., Jaquinet, A., Montandon, D., & Pittet-Cuenod, B. (2018). The surgical management of extra-articular ankylosis in noma patients. International Journal of Oral & Maxillofacial Surgery, 47(12), 1527-1533.
307. Rüegg, E. M., Gniadek, P., Modarressi, A., Baratti-Mayer, D., & Pittet-Cuénod, B. (2016). Facial bone reconstruction with prefabricated vascularized calvarium flaps in children and young adults: advantages and long-term results. Journal of Cranio-Maxillofacial Surgery, 44(12), 1880-1888.
308. Saleh, D. B., et al., (2020). Single-stage nasal reconstruction with the islanded forehead flap. Journal of plastic, reconstructive & aesthetic surgery : JPRAS, 73(9), 1692–1699.
309. Sangani, I., Watt, E., & Cross, D. (2013). Necrotizing ulcerative gingivitis and the orthodontic patient: a case series. Journal of orthodontics, 40(1), 77-80.
310. Sansom, A. E. (1878). On a Case of Noma, in which Moving Bodies were observed in the Blood during Life. Medicochirurgical Transactions, 61, 1-12.11.
311. Sen Gupta, P. C., & Chakravarty, N. K. (1945). Penicillin in Cancrum Oris complicating Kala-Azar. Indian Medical Gazette, 80(11), 542-545.
312. Shaw, J. P. (1896). Cancrum Oris. The American Journal of Dental Science, 30(6), 252-254.
313. Shaye, D. A., Rabbels, J., Adetunji, A. S., Magee, A., Vo, D., & Winters, R. (2018). Evaluation of the Noma Disease Burden Within the Noma Belt. JAMA Facial Plastic Surgery, 20(4), 332-333.
314. Shaye, D. A., Winters, R., Rabbels, J., Adentunji, A. S., Magee, A., & Vo, D. (2019). Noma surgery. Laryngoscope, 129(1), 96-99.
315. Sheiham, A. (1966). An epidemiological survey of acute ulcerative gingivitis in Nigerians. Archives of Oral Biology, 11(9), 937-942.
316. Siddiqui, A. Z., et al., (2020). Bactericidal and clinical efficacy of photochemotherapy in acute necrotizing ulcerative gingivitis. Photodiagnosis and Photodynamic Therapy, 29, 101668.
317. Simon, F., Wolfe, S. A., Guichard, B., Bertolus, C., & Khonsari, R. H. (2015). Paul Tessier facial reconstruction in 1970 Iran, a series of post-noma defects. Journal of Cranio-Maxillo-Facial Surgery, 43(4), 503-509.
318. Singh, A., Mandal, A., Seth, R., & Kabra, S. K. (2016). Noma in a child with acute leukaemia: when the 'face of poverty' finds an ally. BMJ Case Reports, 06, 06.
319. Sirol, J., Vedy, J., & Sabrie, A. (1972). [Noma or forgotten ugliness. Clinical study and pathogenesis apropos of 6 cases at Fort-Lamy (Chad)]. Annales de Dermatologie et de Syphiligraphie, 99(5), 511-516.
320. Smith, E. C. (1926). Cancrum oris in a case of leukæmia. Transactions of the Royal Society of Tropical Medicine and Hygiene, 19(7), 394-396.
321. Srour, M. L., Farley, E., & Mpinga, E. K. (2022). Lao Noma Survivors: A Case Series, 2002-2020. The American journal of tropical medicine and hygiene, 106(4), 1269–1274.
322. Srour, M. L., Watt, B., Phengdy, B., Khansoulivong, K., Harris, J., Bennett, C., . . . Newton, P. N. (2008). Noma in Laos: stigma of severe poverty in rural Asia. American Journal of Tropical Medicine & Hygiene, 78(4), 539-542.
323. ST. BARTHOLOMEW'S HOSPITAL. Cancrum Oris; Recovery. ( Under the care of Dr. BURROWS.). (1851). The Lancet, 57(1449), 621-622.
324. Stark, S. (1956). Noma or gangrenous stomatitis. Report of a case. Oral Surgery, Oral Medicine, Oral Pathology, 9(10), 1076-1079.
325. Stassen, L. F. A., Batchelor, A. G. G., Rennie, J. S., & Moos, K. F. (1989). Cancrum oris in an adult Caucasian female. British Journal of Oral and Maxillofacial Surgery, 27(5), 417-422.
326. Stewart, G. (1868). ROYAL HOSPITAL FOR SICK CHILDREN, EDINBURGH. CASE OF CANCRUM ORIS; RECOVERY. The Lancet, 92(2358), 601.
327. Stewart, M. C. (1911). Observations on the histopathology of cancrum oris. The Journal of Pathology. 16(1), 221-224.
328. Sud, A., Baylis, C., Stanley, I., & Rodney, G. (2020). Airway challenges in Noma patients undergoing facial reconstructive surgery in Ethiopia. Trends in Anaesthesia and Critical Care, 30, e132.
329. Sund, G. C., Muvunyi, P., & Harling, M. J. (2020). Airway Management Through a Facial Defect Resulting From Noma (Orofacial Gangrene): A Case Report. A&A practice, 14(11), e01319.
330. Sykes, L. M., & Essop, R. M. (2000). Combination intraoral and extraoral prosthesis used for rehabilitation of a patient treated for cancrum oris: a clinical report. Journal of Prosthetic Dentistry, 83(6), 613-616.
331. Sykes, L. M., Du Plessis, F., & Essop, R. M. (2004). Cancrum Oris--prosthetic aspects of treatment. SADJ, 59(1), 14-17.
332. Tang Chen, Y. B., Chen, H. C., & Randall, P. (1991). Reconstruction of a compound facial deformity involving the columella, nasal base, and upper lip. Plastic & Reconstructive Surgery, 87(5), 950-953.
333. Tassonyi, E., Lehmann, C., Gunning, K., Coquoz, E., & Montandon, D. (1990). Fiberoptically guided intubation in children with gangrenous stomatitis (noma). Anesthesiology, 73(2), 348-349.
334. Tauber, E. B., & Goldman, L. (1936). Fusospirillary gangrenous stomatitis. Archives of Dermatology and Syphilology, 34(4), 630-634.
335. Tkacz, K., Gill, J., & McLernon, M. (2021). Necrotising periodontal diseases and alcohol misuse - a cause of osteonecrosis?. British dental journal, 231(4), 225–231.
336. Tonna, J. E., Lewin, M. R., & Mensh, B. (2010). A case and review of noma. PLoS Neglected Tropical Diseases, 4(12), 1-2.
337. Toure, S., Siriki, S., Radi, E., & Assi, K. (1991). Surgical prosthetic approach to sequelae of a case of noma involving half the face. [French]. [Approche chirurgico-prothetique des sequelles d'un cas de noma interessant l'hemi-face.]. Odonto-stomatologie tropicale = Tropical dental journal, 14(1), 27-32.
338. Traore, H., et al., (2021). Case report: a rare case of NOMA (cancrum oris) in a Malian woman. New microbes and new infections, 42, 100907.
339. Tungotyo, M. (2017). Noma as a complication of false teeth (Ebiino) extraction: a case report. Journal of Medical Case Reports [Electronic Resource], 11(1), 112.
340. Umeizudike, K. A., et al., (2011). Severe presentation of necrotizing ulcerative periodontitis in a Nigerian HIV-positive patient: a case report. Medical Principles and Practice, 20(4), 374-376.
341. Uri, J. (1947). The role of shedding teeth in the propagation of gangrenous stomatitis. Dental items of interest, 69(5), 401-405.
342. Usher, S. J., & Ross, D. E. (1931). A Case of Cancrum Oris Following Typhoid Fever; with Plastic Repair. Canadian Medical Association Journal, 25(4), 446-449.
343. Vaidya, S., Agrawal, A., & Sharma, V. K. (2006). Cancrum Oris: A case report. Indian Journal of Otolaryngology and Head and Neck Surgery, 58(4), 411-413.
344. Vaidyanathan, S., Tullu, M. S., Lahiri, K., & Deshmukh, C. (2005). Pseudomonas sepsis with Noma: An association? Indian Journal of Medical Sciences, 59(8), 357-360.
345. Vaillant, J. M. and G. Peri (1970). "[A noma which is hard to repair]." Revue de Stomatologie et de Chirurgie Maxillo-Faciale 71(3): 212-214.
346. Vaillant, M. M., et al.,(1967). "[Permanent para-articular constriction of the jaws. Sequels of noma]." Annales de Chirurgie Plastique 12(3): 212-216.
347. Vaizey, J. M. (1946). NOMA TREATED WITH PENICILLIN. British Medical Journal, 2(4461), 14-14.
348. Valadas, G., & Leal, M. J. (1998). Cancrum oris (noma) in children. European Journal of Pediatric Surgery, 8(1), 47-51.
349. van Niekerk, C., Khammissa, R. A., Altini, M., Lemmer, J., & Feller, L. (2014). Noma and cervicofacial necrotizing fasciitis: clinicopathological differentiation and an illustrative case report of noma. AIDS Research & Human Retroviruses, 30(3), 213-216.
350. Van Nitsen, R. (1922). "[English title not available] [not specified]." Annales de la Societe Belge de Medecine Tropicale 2(2-3): 133-134.
351. Venter, T. H., et al., (2023). The Africa Temporal Scalp Flap: A Novel Flap for Facial Reconstruction. Journal of burn care & Research: official publication of the American Burn Association, 44(3), 618–623.
352. Verhoeff, F. H. (1901). A Case of Noma of the Auricles Due to the Streptococcus Pyogenes, and Its Bearing on the Etiology of Noma in General. Journal of the Boston Society of Medical Sciences, 5(10), 465-478.
353. Ver-Or, N., et al., (2022). Retrospective Characterization of Noma Cases Found Incidentally across Nigeria during Outreach Programs for Cleft Lip from 2011-2020. The American journal of tropical medicine and hygiene, 107(5), 1132–1136.
354. Vikram, V., & Rajwanshi, A. (1981). A case of noma with membrane on the throat. Indian Journal of Pediatrics, 48(392), 333-336.
355. Vinzenz, K., Holle, J., & Wuringer, E. (2008). Reconstruction of the maxilla with prefabricated scapular flaps in noma patients. Plastic and Reconstructive Surgery, 121(6), 1964-1973.
356. Weinstein, R. A., Choukas, N. C., & Wood, W. S. (1974). Cancrum oris-like lesion associated with acute myelogenous leukemia. Oral Surgery, Oral Medicine, Oral Pathology, 38(1), 10-14.
357. Weledji, E. P., & Njong, S. (2015). Cancrum Oris (Noma): The Role of Nutrition in Management. The journal of the American College of Clinical Wound Specialists, 7(1-3), 50-52.
358. Wells, R. G., Sty, J. R., & Starshak, R. J. (1988). CT findings in noma. Journal of Computer Assisted Tomography, 12(4), 711-712.
359. White, N. S. (1968). Noma: report of case. Journal of Oral Surgery, 26(6), 418-419.
360. Whiteson, K. L., Lazarevic, V., Tangomo-Bento, M., Girard, M., Maughan, H., Pittet, D., . . . Schrenzel, J. (2014). Noma Affected Children from Niger Have Distinct Oral Microbial Communities Based on High-Throughput Sequencing of 16S rRNA Gene Fragments. PLoS Neglected Tropical Diseases, 8 (12) (no pagination)(e3240).
361. Woon, C. Y., Sng, K. W., Tan, B. K., & Lee, S. T. (2010). CASE REPORT Journey of a Noma Face. Eplasty [Electronic Resource], 10, e49.
362. Xu, L., et al., (2019). Noma in a boy with septic shock: a case report. BMC pediatrics, 19(1), 200.
363. Yates, P., & Kingsford, E. C. (1889). CANCRUM ORIS, AND ITS SUCCESSFUL TREATMENT BY THE LOCAL APPLICATION OF CORROSIVE SUBLIMATE. The Lancet, 133(3427), 880-882.
364. Yuca, K., Yuca, S. A., Cankaya, H., Caksen, H., Calka, O., & Kiris, M. (2004). Report of an infant with noma (cancrum oris). Journal of Dermatology, 31(6), 488-491.
365. Zawar, V., Pawar, M., Kumavat, S., Shah, M., & Chaddha, C. (2018). A progressive ulcer in immunocompetent man: cancrum oris. Tropical Doctor, 48(1), 60-62.
366. Zee, Z. U., & Paty, R. M., Jr. (1937). Gangrenous Stomatitis with Special Reference to Subtertian Malaria as an Etiological Factor. Chinese Medical Journal, 52(1), 95-100.

### 7. PRISMA CHECKLIST

| **Section and Topic** | **Item #** | **Checklist item** | **Location where item is reported** |
| --- | --- | --- | --- |
| **TITLE** | | |  |
| Title | 1 | Identify the report as a systematic review. | Title |
| **ABSTRACT** | | |  |
| Abstract | 2 | See the PRISMA 2020 for Abstracts checklist. | Abstract |
| **INTRODUCTION** | | |  |
| Rationale | 3 | Describe the rationale for the review in the context of existing knowledge. | Introduction |
| Objectives | 4 | Provide an explicit statement of the objective(s) or question(s) the review addresses. | Introduction; Protocol Publication^ |
| **METHODS** | | |  |
| Eligibility criteria | 5 | Specify the inclusion and exclusion criteria for the review and how studies were grouped for the syntheses. | Methods; Supplementary File 1; Protocol Publication^ |
| Information sources | 6 | Specify all databases, registers, websites, organisations, reference lists and other sources searched or consulted to identify studies. Specify the date when each source was last searched or consulted. | Methods; Supplementary File 1 |
| Search strategy | 7 | Present the full search strategies for all databases, registers and websites, including any filters and limits used. | Supplementary File 1 |
| Selection process | 8 | Specify the methods used to decide whether a study met the inclusion criteria of the review, including how many reviewers screened each record and each report retrieved, whether they worked independently, and if applicable, details of automation tools used in the process. | Supplementary File 1; Protocol Publication^ |
| Data collection process | 9 | Specify the methods used to collect data from reports, including how many reviewers collected data from each report, whether they worked independently, any processes for obtaining or confirming data from study investigators, and if applicable, details of automation tools used in the process. | Methods; Protocol Publication^ |
| Data items | 10a | List and define all outcomes for which data were sought. Specify whether all results that were compatible with each outcome domain in each study were sought (e.g. for all measures, time points, analyses), and if not, the methods used to decide which results to collect. | Supplementary File 2_Variable Dictionary; Protocol Publication^ |
|  | 10b | List and define all other variables for which data were sought (e.g. participant and intervention characteristics, funding sources). Describe any assumptions made about any missing or unclear information. | Supplementary File 2_Variable Dictionary; Protocol Publication^ |
| Study risk of bias assessment | 11 | Specify the methods used to assess risk of bias in the included studies, including details of the tool(s) used, how many reviewers assessed each study and whether they worked independently, and if applicable, details of automation tools used in the process. | Methods; Supplementary File 3_Section 9; Protocol Publication^ |
| Effect measures | 12 | Specify for each outcome the effect measure(s) (e.g. risk ratio, mean difference) used in the synthesis or presentation of results. | Not applicable. Largely descriptive synthesis. See Methods and Protocol Publication^ |
| Synthesis methods | 13a | Describe the processes used to decide which studies were eligible for each synthesis (e.g. tabulating the study intervention characteristics and comparing against the planned groups for each synthesis (item #5)). | Methods; Protocol Publication^; Supplementary File 1_Table2; Supplementary File 2_Variable Dictionary |
|  | 13b | Describe any methods required to prepare the data for presentation or synthesis, such as handling of missing summary statistics, or data conversions. | Methods; Protocol Publication^; Supplementary File 2_Variable Dictionary |
|  | 13c | Describe any methods used to tabulate or visually display results of individual studies and syntheses. |  |
|  | 13d | Describe any methods used to synthesize results and provide a rationale for the choice(s). If meta-analysis was performed, describe the model(s), method(s) to identify the presence and extent of statistical heterogeneity, and software package(s) used. | Methods; Protocol Publication^ |
|  | 13e | Describe any methods used to explore possible causes of heterogeneity among study results (e.g. subgroup analysis, meta-regression). | Not applicable |
|  | 13f | Describe any sensitivity analyses conducted to assess robustness of the synthesized results. | Not applicable |
| Reporting bias assessment | 14 | Describe any methods used to assess risk of bias due to missing results in a synthesis (arising from reporting biases). | Methods; Protocol Publication^; Supplementary File 3_Section 9 |
| Certainty assessment | 15 | Describe any methods used to assess certainty (or confidence) in the body of evidence for an outcome. |  |
| **RESULTS** | | |  |
| Study selection | 16a | Describe the results of the search and selection process, from the number of records identified in the search to the number of studies included in the review, ideally using a flow diagram. | Supplementary File 1 |
|  | 16b | Cite studies that might appear to meet the inclusion criteria, but which were excluded, and explain why they were excluded. | Supplementary File 2_Excluded at Full-text |
| Study characteristics | 17 | Cite each included study and present its characteristics. | Supplementary File 2_Data |
| Risk of bias in studies | 18 | Present assessments of risk of bias for each included study. | Supplementary File 2_Data |
| Results of individual studies | 19 | For all outcomes, present, for each study: (a) summary statistics for each group (where appropriate) and (b) an effect estimate and its precision (e.g. confidence/credible interval), ideally using structured tables or plots. | Supplementary File 2_Data |
| Results of syntheses | 20a | For each synthesis, briefly summarise the characteristics and risk of bias among contributing studies. | Results; Supplementary File 3; |
|  | 20b | Present results of all statistical syntheses conducted. If meta-analysis was done, present for each the summary estimate and its precision (e.g. confidence/credible interval) and measures of statistical heterogeneity. If comparing groups, describe the direction of the effect. |  |
|  | 20c | Present results of all investigations of possible causes of heterogeneity among study results. |  |
|  | 20d | Present results of all sensitivity analyses conducted to assess the robustness of the synthesized results. |  |
| Reporting biases | 21 | Present assessments of risk of bias due to missing results (arising from reporting biases) for each synthesis assessed. | Results; Supplementary File 3_Section 9 |
| Certainty of evidence | 22 | Present assessments of certainty (or confidence) in the body of evidence for each outcome assessed. |  |
| **DISCUSSION** | | |  |
| Discussion | 23a | Provide a general interpretation of the results in the context of other evidence. | Discussion; SF3 |
|  | 23b | Discuss any limitations of the evidence included in the review. | Risk of Bias; SF3; Discussion; Limitations |
|  | 23c | Discuss any limitations of the review processes used. | Discussion; Limitations |
|  | 23d | Discuss implications of the results for practice, policy, and future research. | Discussion; Box 1; Conclusion; Panel: research in context |
| **OTHER INFORMATION** | | |  |
| Registration and protocol | 24a | Provide registration information for the review, including register name and registration number, or state that the review was not registered. | Methods |
|  | 24b | Indicate where the review protocol can be accessed, or state that a protocol was not prepared. |  |
|  | 24c | Describe and explain any amendments to information provided at registration or in the protocol. |  |
| Support | 25 | Describe sources of financial or non-financial support for the review, and the role of the funders or sponsors in the review. | Funding Information |
| Competing interests | 26 | Declare any competing interests of review authors. | Declaration of interests |
| Availability of data, code and other materials | 27 | Report which of the following are publicly available and where they can be found: template data collection forms; data extracted from included studies; data used for all analyses; analytic code; any other materials used in the review. | Data Sharing |

PRISMA CHECKLIST From: Page MJ, McKenzie JE, Bossuyt PM, Boutron I, Hoffmann TC, Mulrow CD, et al. The PRISMA 2020 statement: an updated guideline for reporting systematic reviews. BMJ 2021;372:n71. doi: 10.1136/bmj.n71

^ PROTOCOL PUBLICATION: Maguire BJ, Shrestha P, Rashan S et al. Protocol for a systematic review of the evidence-based knowledge on the distribution, associated risk factors, the prevention and treatment modalities for noma. Wellcome Open Research 2023, 8:125 https://doi.org/10.12688/wellcomeopenres.19033.1
