## Supplementary material for "A systematic review of the noma evidence landscape: current knowledge and gaps": SF3. Supplementary File 3_Extended Results

### 1. Study Characteristics

##### a. Spatial distribution

Figure 1 and Figure 2 map the spatial distribution of noma studies between 2000-2009 and 2010-2022, respectively. Additional maps for the periods pre-1900, 1900 to 1940 (prior to widely accessible antibiotics), and 1941 to 1999 can be found in Figure 3.

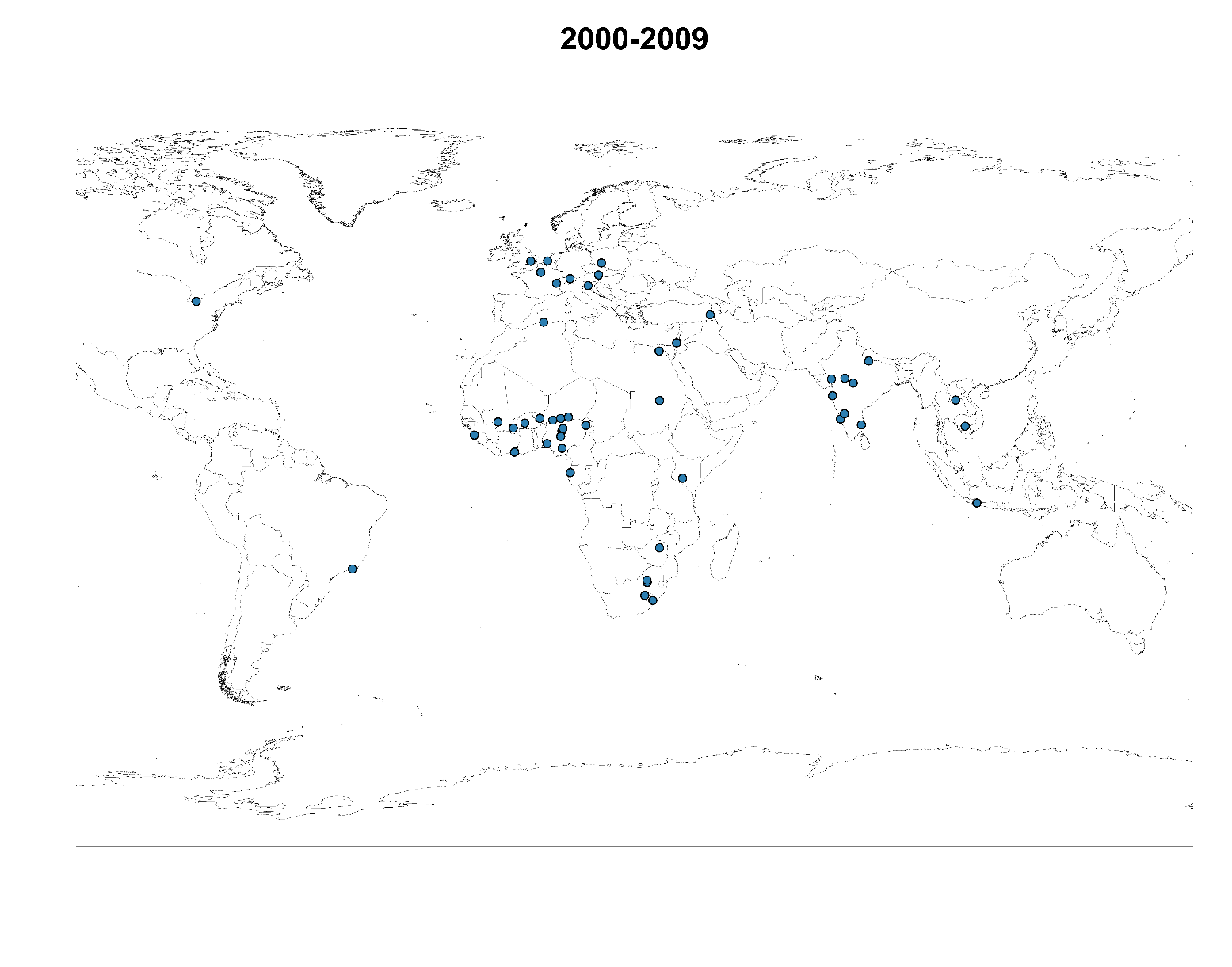

Figure 1	Spatial distribution of noma studies between 2000 and 2009

Legend: The dots simply represent the study sites and are not weighted for the number of studies or the number of patients reported. Refer to Figure 3 for additional time periods.

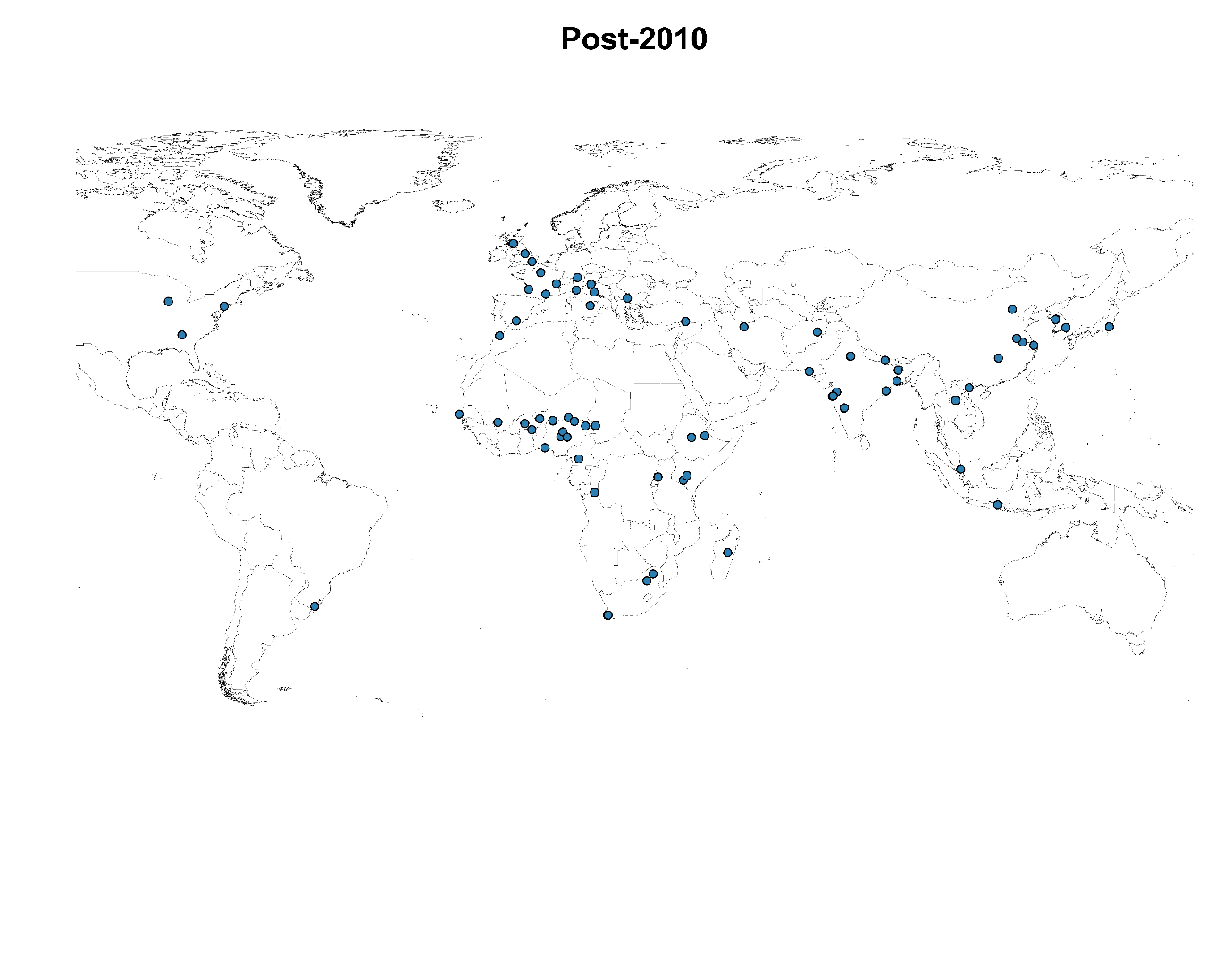

Figure 2	Spatial distribution of noma studies between 2010 and 2022^*^

Legend: The dots simply represent the study sites and are not weighted for the number of studies or the number of patients reported. Refer to Figure 3 for additional time periods.

* Until 07 December 2022

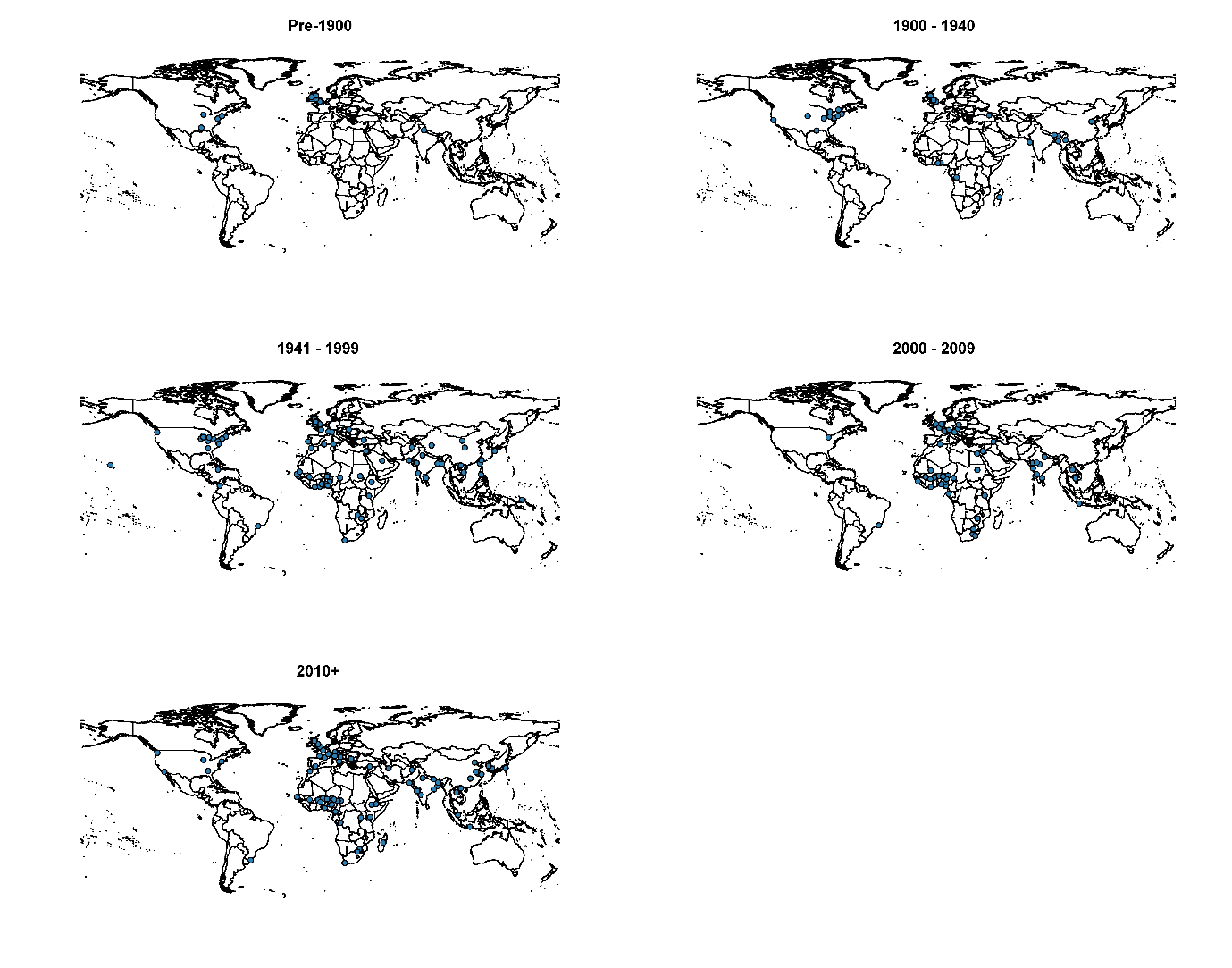

Figure 3	Spatial distribution of noma studies 1839 to 7 December 2022

Legend: The dots simply represent the study sites and are not weighted for the number of studies or the number of patients reported

##### b. Patient numbers by country

The number of patients enrolled or reported per country of study and by study design are presented in Figure 4 for all countries where more than 10 patients were reported. The number of patients by study design and region for all studies are presented in Figure 5. The number of noma patients using the WHO regional classification is provided in Figure 6 only. Included patient numbers for all countries are also reported in Table 1.

Some of the largest numbers of noma patients per country reported in the included studies were from the “noma belt” region, deemed in the literature as the most prevalent region for cases (Brattström-Stolt *et al.,* 2019). Nigeria (n=10,643), Niger (n=887), Burkina Faso (n=580), Ethiopia (n=381) and Senegal (n=324). Yet, there was limited (Ghana, Mali, Chad, Cameroon, Togo, Benin) or no data (Central Africa Republic, South Sudan) reported from other “noma belt” countries. This could be explained by the weak health surveillance system in the region and/or lack of reporting of diseases, in addition to limited knowledge of noma among the healthcare providers, primarily due to a lack of exposure in the medical training programs (Brattström-Stolt *et al.,* 2019; Ahlgren *et al.,* 2017; Farley *et al.,* 2022)

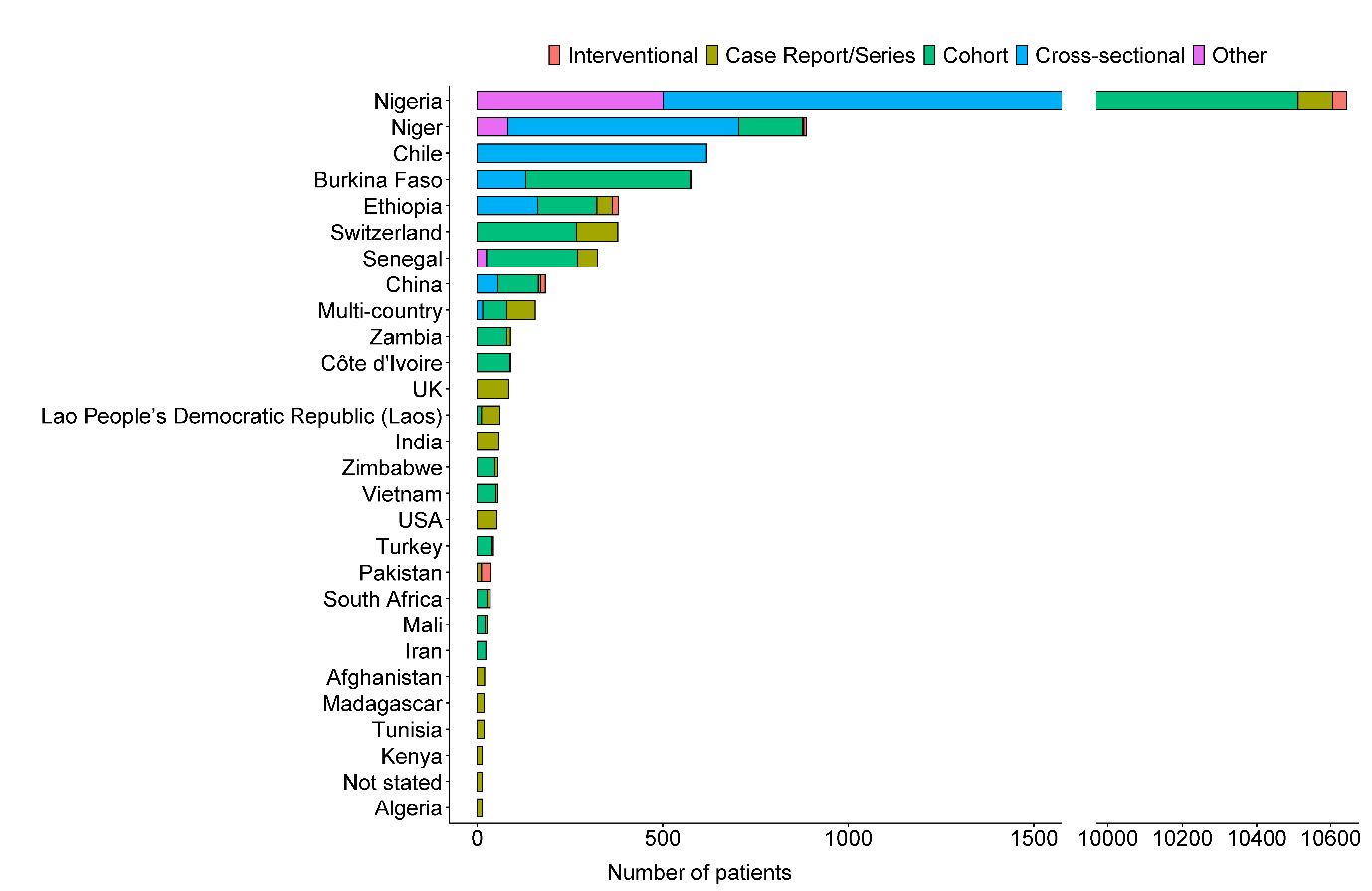

Figure 4	Number of patients (n= 15,082 patients) reported per country of study and by study design (note that the X axis is discontinued to account for the representation of a very large number of patients from Nigeria; data is shown only when there were >10 patients reported per country)

Refer to Table 1 for further detail; Some or all of the patients from Benin, Ethiopia, UK, Togo, Mali, Nigeria, Niger, Guinea-Bissau, Republic of Congo, Madagascar, Cameroon, and Senegal are presented under multi-country

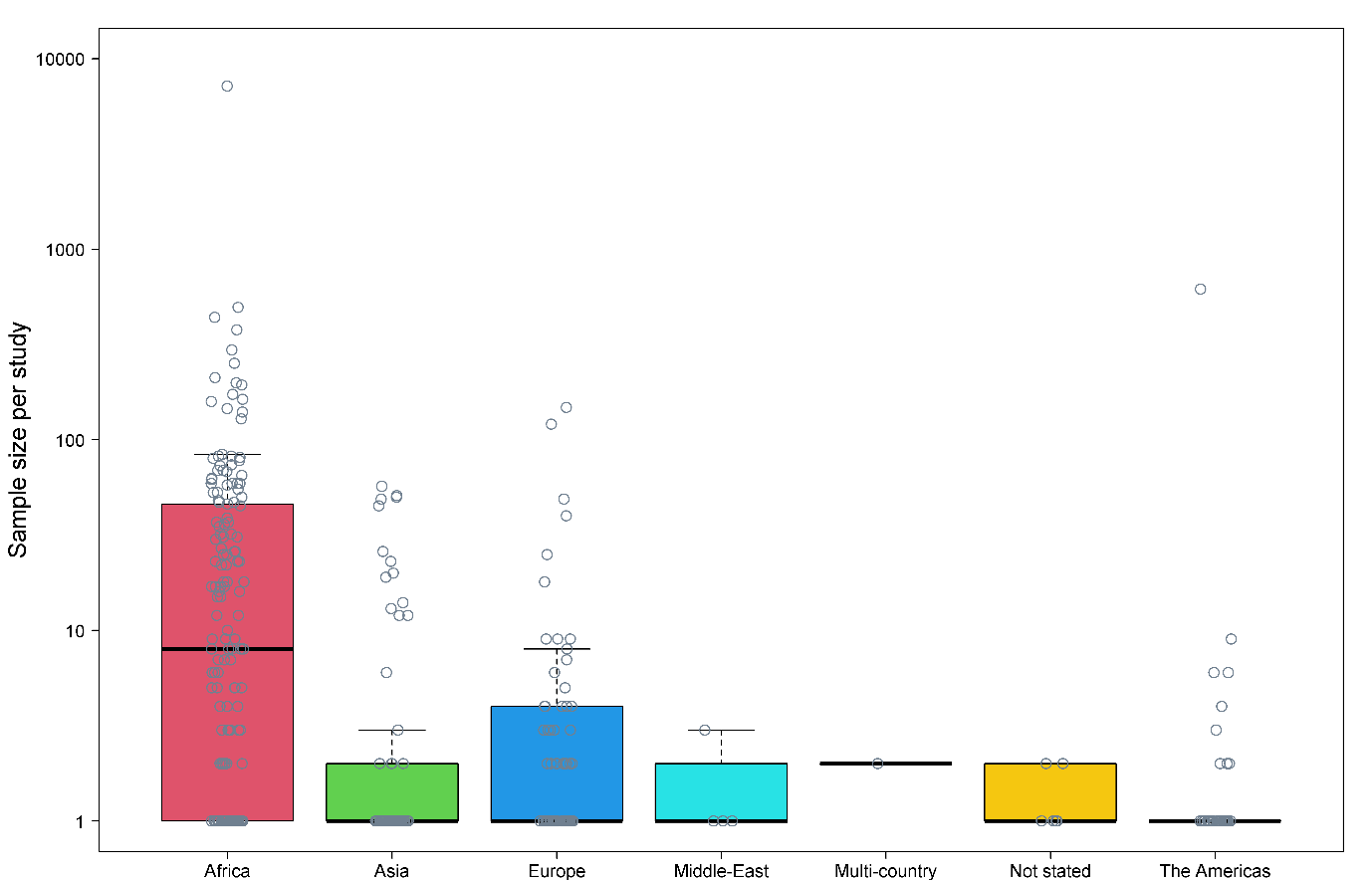

Figure 5	Number of included noma patients per study by region

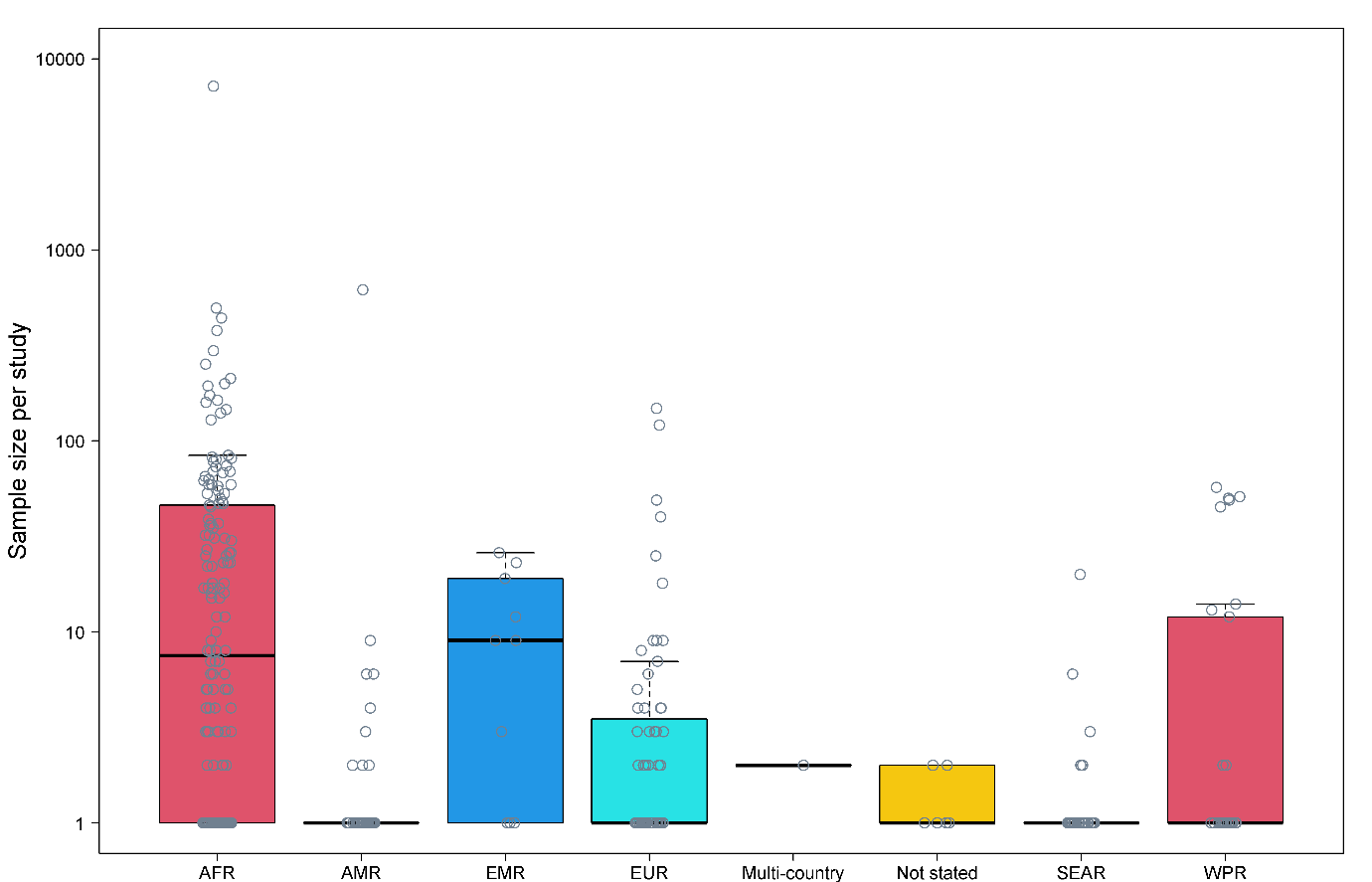

Figure 6	Number of included noma patients per study by WHO region classification

AFR = African region, AMR = Region of the Americas, EMR = Eastern Mediterranean region; EUR= European region, SEAR = South-East Asian region, WPR= Western Pacific region; Not stated = country not specificied; one multi-country study with locations in UK and Ethiopia

Table 1	Total number of patients reported per country from 1839 to 7 December 2022

| **Country** | **Number of studies** | **Number of patients** |
| --- | --- | --- |
| Afghanistan | 2 | 20 |
| Algeria | 6 | 13 |
| Austria | 1 | 9 |
| Bangladesh | 2 | 2 |
| Brazil | 3 | 8 |
| Bulgaria | 1 | 1 |
| Burkina Faso | 9 | 580 |
| Burundi | 1 | 1 |
| Cambodia | 1 | 1 |
| Cameroon | 1 | 1 |
| Chad | 1 | 6 |
| Chile | 1 | 618 |
| China (1 case in Taiwan) | 12 | 185 |
| Colombia | 1 | 9 |
| Côte d'Ivoire | 3 | 90 |
| Croatia | 1 | 1 |
| Democratic Republic of Congo | 2 | 3 |
| Ethiopia | 10 | 381 |
| France | 5 | 8 |
| Gabon | 1 | 1 |
| Gambia | 1 | 6 |
| Germany | 1 | 4 |
| Ghana | 1 | 1 |
| Guinea | 1 | 5 |
| Guinea-Bissau | 1 | 1 |
| Hungary | 1 | 1 |
| India | 31 | 59 |
| Indonesia | 2 | 2 |
| Iran | 1 | 23 |
| Ireland | 1 | 1 |
| Israel | 3 | 3 |
| Italy | 5 | 7 |
| Jamaica | 1 | 1 |
| Japan | 2 | 2 |
| Kenya | 5 | 14 |
| Lao People's Democratic Republic (Laos) | 2 | 62 |
| Lesotho | 1 | 2 |
| Madagascar | 2 | 19 |
| Mali | 4 | 27 |
| Micronesia | 1 | 2 |
| Morocco | 2 | 2 |
| Multi-country | 6 | 157 |
| Myanmar | 1 | 1 |
| Namibia | 1 | 1 |
| Nepal | 1 | 1 |
| Niger | 14 | 887 |
| Nigeria | 62 | 10643 |
| Not stated | 7 | 13 |
| Pakistan | 2 | 38 |
| Papua New Guinea | 1 | 1 |
| Philippines | 1 | 1 |
| Portugal | 1 | 1 |
| Saudi Arabia | 1 | 3 |
| Senegal | 10 | 324 |
| Singapore | 1 | 1 |
| Slovenia | 1 | 1 |
| South Africa | 11 | 36 |
| South Korea | 3 | 3 |
| Spain | 1 | 2 |
| Switzerland | 8 | 379 |
| Togo | 2 | 2 |
| Tunisia | 2 | 18 |
| Turkey | 4 | 44 |
| Uganda | 1 | 1 |
| UK | 39 | 85 |
| USA | 41 | 54 |
| Vietnam | 5 | 56 |
| Zambia | 3 | 90 |
| Zimbabwe | 3 | 57 |

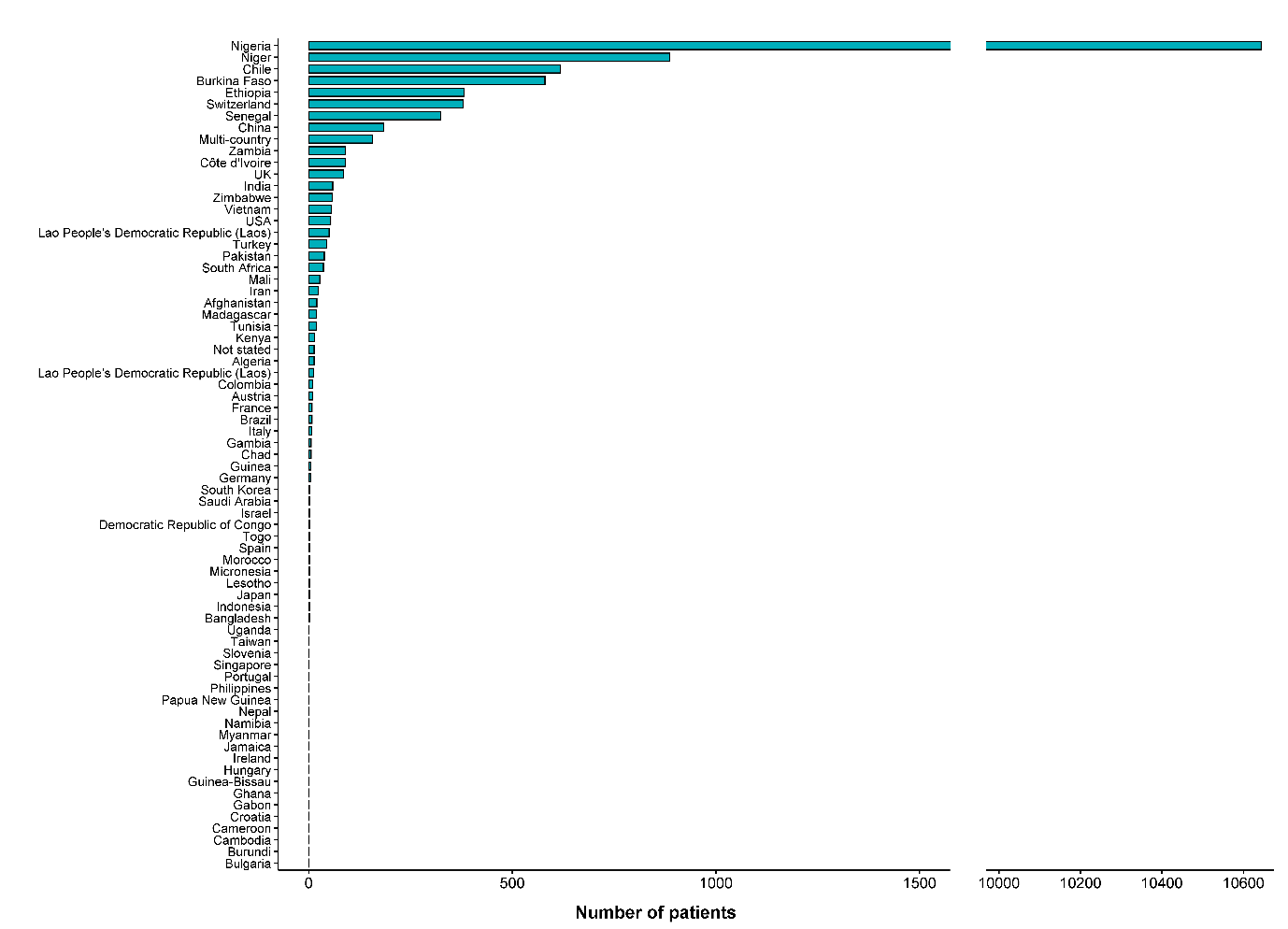

Figure 7 Number of patients (n=15,082 patients) reported per country

Refer to Table 1 for further detail

Table 2	Noma cases reported before 1950 by country

| **Country** | **Number of studies** | **Number of patients** |
| --- | --- | --- |
| Bangladesh | 2 | 2 |
| China | 6 | 179 |
| Democratic Republic of Congo | 1 | 1 |
| Ethiopia | 1 | 3 |
| Hungary | 1 | 1 |
| India | 6 | 14 |
| Ireland | 1 | 1 |
| Madagascar | 1 | 18 |
| Myanmar | 1 | 1 |
| Nigeria | 1 | 1 |
| Not stated | 5 | 10 |
| Togo | 1 | 1 |
| Turkey | 1 | 40 |
| UK | 29 | 70 |
| USA | 18 | 26 |
| Vietnam | 1 | 1 |

Table 3	Noma cases reported between 1950 and 2000 by country

| **Country** | **Number of studies** | **Number of patients** |
| --- | --- | --- |
| Nigeria | 17 | 693 |
| Senegal | 9 | 308 |
| Zambia | 3 | 90 |
| Vietnam | 3 | 54 |
| Niger | 1 | 50 |
| Switzerland | 3 | 30 |
| India | 7 | 27 |
| South Africa | 1 | 26 |
| Ethiopia | 1 | 25 |
| Mali | 1 | 22 |
| USA | 17 | 22 |
| Afghanistan | 1 | 19 |
| Tunisia | 2 | 18 |
| Pakistan | 1 | 12 |
| Kenya | 2 | 11 |
| Algeria | 5 | 10 |
| Colombia | 1 | 9 |
| Multi-country | 1 | 8 |
| Zimbabwe | 1 | 8 |
| Brazil | 1 | 6 |
| Chad | 1 | 6 |
| Gambia | 1 | 6 |
| UK | 4 | 5 |
| France | 1 | 4 |
| Not stated | 2 | 3 |
| Saudi Arabia | 1 | 3 |
| Israel | 2 | 2 |
| Micronesia | 1 | 2 |
| Turkey | 1 | 2 |
| Côte d'Ivoire | 1 | 1 |
| Ghana | 1 | 1 |
| Italy | 1 | 1 |
| Jamaica | 1 | 1 |
| Japan | 1 | 1 |
| Morocco | 1 | 1 |
| Papua New Guinea | 1 | 1 |
| Philippines | 1 | 1 |
| Portugal | 1 | 1 |
| China (case in Taiwan) | 1 | 1 |

Table 4	Noma cases reported after 2000 by country

| **Country** | **Number of studies** | **Number of patients** |
| --- | --- | --- |
| Nigeria | 44 | 9,949 |
| Niger | 13 | 837 |
| Chile | 1 | 618 |
| Burkina Faso | 9 | 580 |
| Ethiopia | 8 | 353 |
| Switzerland | 5 | 349 |
| Multi-country | 5 | 149 |
| Côte d'Ivoire | 2 | 89 |
| Lao People's Democratic Republic (Laos) | 2 | 62 |
| Zimbabwe | 2 | 49 |
| Pakistan | 1 | 26 |
| Iran | 1 | 23 |
| India | 18 | 18 |
| Senegal | 1 | 16 |
| South Africa | 10 | 10 |
| UK | 6 | 10 |
| Austria | 1 | 9 |
| Italy | 4 | 6 |
| USA | 6 | 6 |
| China | 5 | 5 |
| Guinea | 1 | 5 |
| Mali | 3 | 5 |
| France | 4 | 4 |
| Germany | 1 | 4 |
| Algeria | 1 | 3 |
| Kenya | 3 | 3 |
| South Korea | 3 | 3 |
| Brazil | 2 | 2 |
| Democratic Republic of Congo | 1 | 2 |
| Indonesia | 2 | 2 |
| Lesotho | 1 | 2 |
| Spain | 1 | 2 |
| Turkey | 2 | 2 |
| Afghanistan | 1 | 1 |
| Bulgaria | 1 | 1 |
| Burundi | 1 | 1 |
| Cambodia | 1 | 1 |
| Cameroon | 1 | 1 |
| Croatia | 1 | 1 |
| Gabon | 1 | 1 |
| Guinea-Bissau | 1 | 1 |
| Israel | 1 | 1 |
| Japan | 1 | 1 |
| Madagascar | 1 | 1 |
| Morocco | 1 | 1 |
| Namibia | 1 | 1 |
| Nepal | 1 | 1 |
| Singapore | 1 | 1 |
| Slovenia | 1 | 1 |
| Togo | 1 | 1 |
| Uganda | 1 | 1 |
| Vietnam | 1 | 1 |

##### c. Patient age

Patients aged less than 19 years were enrolled in 174 studies (n=3,346 patients); these include 4 studies in infants (<1 year), 107 studies in children between 1-10 years, 41 studies that stated children (0-19 years) as the study population, and 22 studies in adolescent population (10-19 years). Adults (>19 years) were enrolled in 98 studies (n=143 patients), patients of all ages were reported in 73 studies (n=11,070 patients), and the age-distribution of the patients was not reported in 21 (n=523 patients). The number of studies by age-distribution is presented in Figure 8.

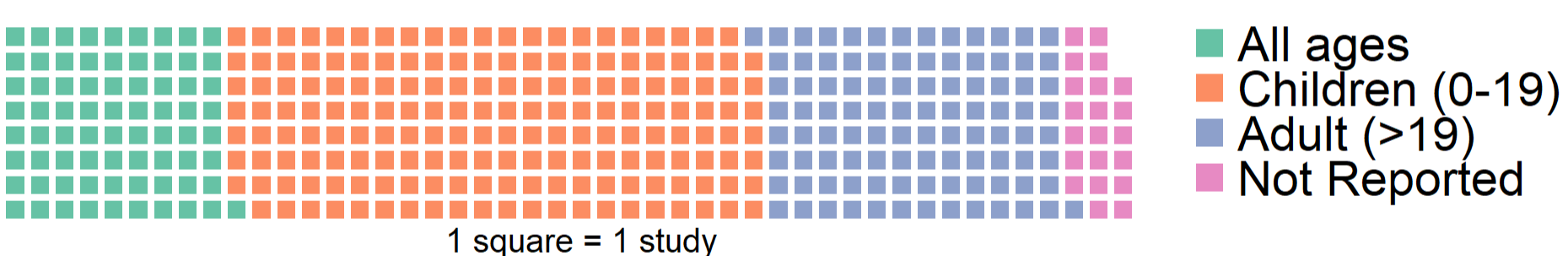

Figure 8	Number of included studies by age group

##### d. Noma stage & study design

The study design for the majority of publications were case reports (270 studies, 884 patients). The greatest volume of individual noma patients reported in the literature were from 29 cross-sectional studies (n=10,601 patients) and 53 cohort studies (n=2,888 patients). A further 8 studies had other observational designs such as case-control or mixed methods studies and included 608 patients. The median [range] number of patients included was: 1 [1-69] for case report/case series, 40 [2-212] for cohort studies, 57 [2-7195] for cross-sectional studies, 16[7-26] for interventional studies, and 65 [8-296] for other study designs. There were 42 (11.4%) studies that included more than 50 patients with cohort and cross-sectional studies from Nigeria (n=15), Niger (n=3), Burkina Faso (n=7), China (n=1), Senegal (n=1), Switzerland (n=2) and Zambia (n=1).

Publications primarily reporting on patients in the acute necrotising gingivitis (ANG) stage formed 9.6% (35/366) of all included studies. Most patients in this stage are attributed to 2 cohorts and 2 cross-sectional studies (Table 5).

Publications primarily reporting patients in the oedema stage represented 7.4% (27/366) of all included studies. However, as all studies in this stage were case reports/series, only 33 noma patients are described.

Studies with patients reported in the gangrenous stage represent 15.6% of all included studies (57/366). Apart from a single cohort study, all other studies were case reports/series. Therefore, total number of patients with gangrenous stage of the disease was 94 and the median number of patients in each study was 1 (range: 1-18).

Of the WHO categories for noma, few studies were attributed to the scarring stage, which only formed 0.8% of the included studies (3/366). These 3 studies were case reports, and all included information on the administered treatment.

Studies in the sequalae stage account for 20.8% of all included studies (76/366). Of the 76 studies, 55 are case reports/series, 12 are cohort studies, 5 are cross-sectional. The sequalae stage also contains 4 of the 6 interventional studies included in this review (Table 5).

Studies assigned to the multiple stages of noma category represented 34.7% of all included studies (127/366). This category contained 84/127 case reports/series, 27/127 cohort studies, 12/127 cross-sectional studies, 1/127 interventional study, and 1 study with other design (Table 5).

A further breakdown of the disease staging by time-period and geographical region is presented in Figure9. There was a distinct variation in noma stages across the regions. In Africa, of the 13,372 patients, 290 were in the acute necrotising gingivitis stage, 51 patients were between stages oedema/gangrenous/scarring stage, 684 in the sequelae stage, 10,567 patients were reported as being in multiple stages, and a further 1,780 patients with unclear disease staging. In Asia, the majority of studies and patients were categorised across multiple stages (216/398 patients). In Europe, most of the patients presented with sequelae stage (344/544 patients) whereas in the Americas, majority of the patients were in ANG stage (626/690 patients). Across the time-periods, the majority of the patients presented with multiple stages of the disease, although number of studies reporting the sequelae stages has steadily increased from 1975 onwards (Figure 9A and 9B).

Table 5	Study design by noma stage

| **Noma Stage** | **Case Report/Series** | | **Cohort** | | **Cross-sectional** | | **Interventional** | | **Other** | |
| --- | --- | --- | --- | --- | --- | --- | --- | --- | --- | --- |
|  | No. of studies | No. of cases | No. of studies | No. of cases | No. of studies | No. of cases | No. of studies | No. of cases | No. of studies | No. of cases |
| 1. Acute necrotizing gingivitis | 28 | 42 | 2 | 150 | 4 | 742 | 1 | 26 | - | - |
| 2. Oedema Stage | 27 | 33 | - | - | - | - | - | - | - | - |
| 3. Gangrenous Stage | 56 | 85 | 1 | 9 | - | - | - | - | - | - |
| 4. Scarring Stage | 3 | 3 | - | - | - | - | - | - | - | - |
| 5. Sequelae Stage | 55 | 320 | 12 | 467 | 5 | 271 | 4 | 62 | - | - |
| Multiple Stages | 84 | 362 | 27 | 1,667 | 12 | 8,775 | 1 | 13 | 3 | 164 |
| Unclear | 17 | 39 | 11 | 595 | 8 | 813 | - | - | - | 444 |

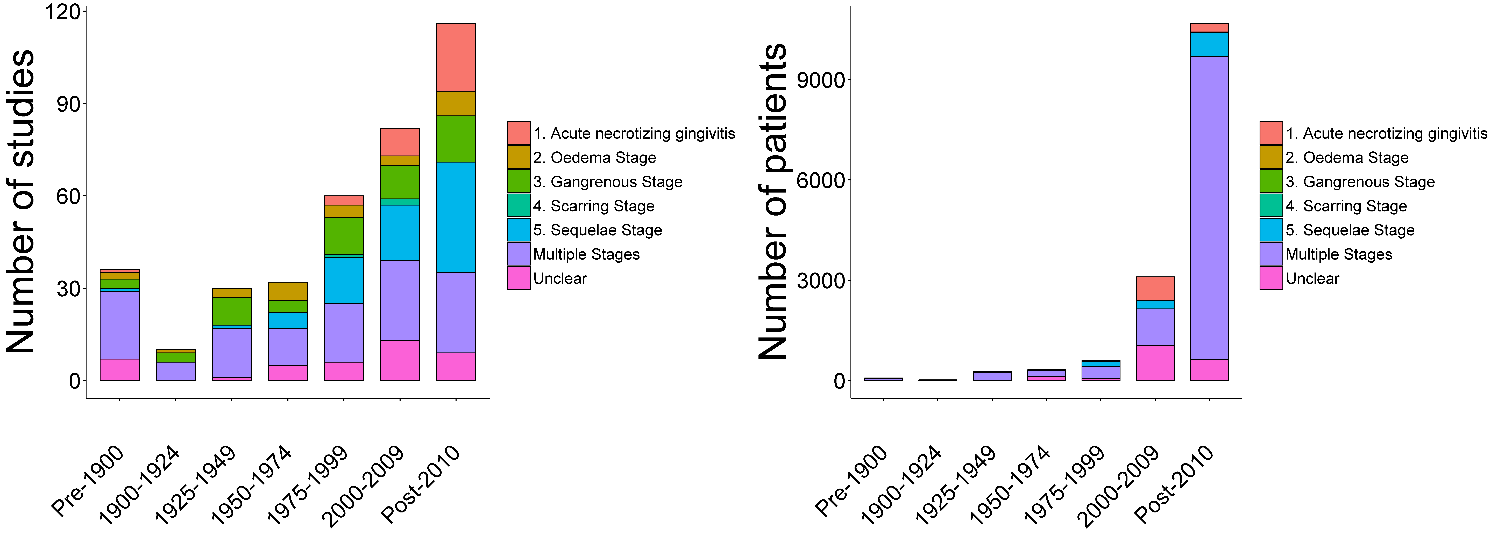

A. B.

Figure 9	A. Number of studies / B. Number of patients – by publication date and noma stage

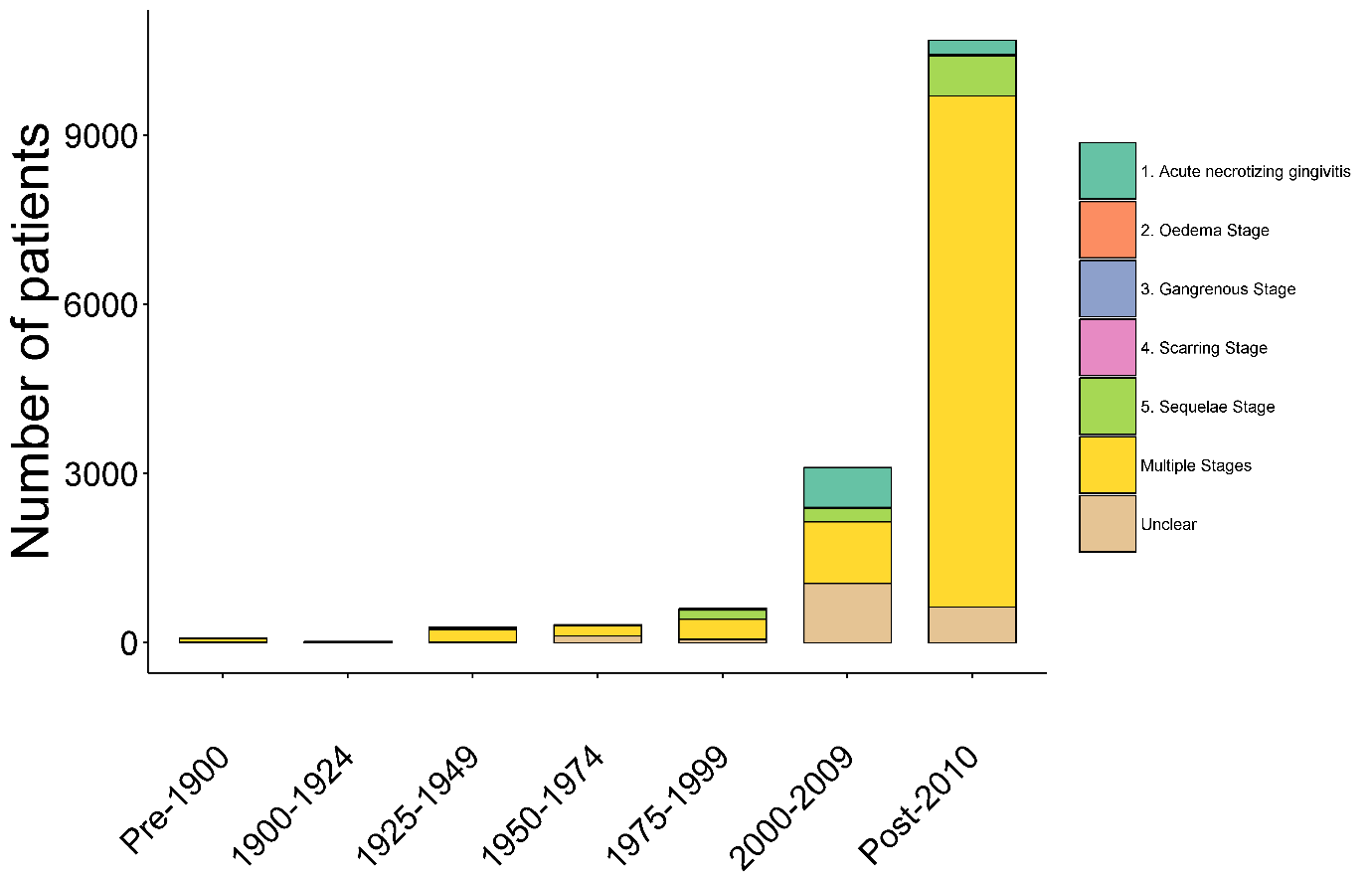

Figure 10	Number of included patients by date of publication and reported noma stage

### 3. Risk Factors

The total number of studies that observed risk factors for noma within each of these categories is presented in Table 6.

Table 6	Number of studies reporting risk factors

| **Category of risk factor** | **Number of studies** |
| --- | --- |
| Immunosuppression^a^ | 61 |
| Nutritional deficit | 142 |
| Poor oral hygiene | 75 |
| Infectious diseases comorbidities | 152 |
| Respiratory infections | 53 |
| Other diseases or clinical comorbidities | 149 |
| Physical environment | 39 |
| Socio economic factors | 65 |
| Lifestyle factors | 41 |

^a^ non-infectious comorbidities causing immunosuppression.

Table 7 provides a de-duplicated and exhaustive list of all potential risk factors associated with reported cases or the noma study populations in the included studies. Each risk factor is listed as it directly appears in the publications. Figure 11 illustrates the frequency of reported risk factors.

Table 7	List of reported risk factors from included Noma studies

| Acute myeloid leukaemia | Gaucher's disease | Pertussis |
| --- | --- | --- |
| advanced pnuemonic phthisis | general malaise | Plaque |
| age | genitourinary gonococcal infection | Pneumococcal disease |
| age of mother | gonorrhea | Pneumonia |
| Agranulocytosis | gum disease | Polio |
| alcoholism | halitosis | polyadenopathies |
| anaemic | headaches | polymorphic reticulosis |
| anorexia | height | poor living conditions |
| ariboflavinosis | hemoptysis | poor oral hygeine |
| ascites | hepatitis | poor ventilation in house |
| atrial flutter | hermes simple virus | poorly built |
| bacillary dysentry | Herpes Simplex Virus | poverty |
| bilateral mandibular hypoplasis | Herpes Zoster | pre-kwoshiorkor |
| bronchiectasis | HIV | presence of disease in last three months |
| bronchiolitis | human cytomegalovirus | presence of larvae in mouth |
| bronchitis | human herpes virus 6 | proptosis |
| broncho-pnuemonitis | hyperlipidemia | pruritus |
| Burkitt tumour | hypertension | psuedomonas |
| Burkitt's lymphoma | hypertensive cardiovascular renal disease | Pyelonephritis |
| cachexia | hyrocephalus internus during her infancy | quinsy |
| cancer | icterus | rachitic rosary |
| candidiasis bucco-pharyngeal | Immunosuppression | Recurrent UTI |
| cardiac condition | impacted teeth | retroviral infection |
| Celiac disease | infantile remittent fever | Rheumatoid arthritis |
| cerebrovascular accident | insect bite | rhuematic fever |
| Chickenpox | intestinal parasitosis | rhuematism |
| child>7 months when first given water | intravenous drug abuse | ring worms |
| Chlamydia | kwashiorkor | rubella |
| chronic lung disease | large family unit | rubeola |
| Chronic Lymphocytic leukemia | length of pregnancy and breastfeeding | scarlatina |
| chronic obstructive pulmonary disease | lethargy | scarlet fever |
| chronic sinusitis | limited toilet facilities | SCID |
| colustrum | lived next to animals | scoliosis |
| constipation | liver disease | sepsis |
| contaminated water supply | living with someone with typhoid | septicaemia |
| cough | lobar pnuemonia | severe burns |
| crepitus | loss of teeth | skin infection |
| croup | low protein diet | small pox |
| cystic fybrosis | luekemia | smoking |
| degenerative disc disease | lymphatic disease | splenomegaly |
| dehydration | malaria | stomatitis |
| Dental caries | malaria | Stress |
| depression | malnourished | strumous |
| diabetes | marasmus | swelling of feet |
| diarrhea | measles | syphilis |
| difficulties with deglutition | medication in past year | T cell acute lymphoblastic leukaemia |
| diffuse peritonitis | meningitis | tachycardia |
| diptheria | mixed non-labeled connective tissue disease | Tetanus |
| disorded dental arrangement | Monolateral acute necrotizing tonsillitis | thrombocytopenic |
| Diverticular disease | mother being primary caretaker | tongue furred |
| Down syndrome | mucormycosis | tool used to clean teeth |
| dyspnea | myelodysplastic syndrome | tooth decay |
| E.coli | myeloid metaplasia | tooth extraction |
| elephantiasis | myocarditis | trismus |
| endstage renal disease | nephritis | tuberculosis |
| entercolitis | no vaccinations | typhoid fever |
| eosinophilia | nocturia | typhus |
| Epigastralgia | nomadic tribal environment | ulcer |
| Epstein–Barr virus type 1 | number of mother's past pregnancies | upper and lower airway infections |
| Eruptive fever | obstructive deep apnea | upper respiratory tract infection |
| erysipelas | one of many children | variola |
| ethanol abuse | oral thrush | venous disease |
| exanthematous fever | osteoarthritis | vertigo |
| exposure to livestock | osteogenesis imperfecta | visceral leishmaniasis |
| fever | otalgia | wasting |
| fish bone trauma | otitis media | weight |
| foetor oris | otorrhoea | weight loss |
| food debris present in teeth | palpitations | wet season |
| frontal headaches | Patent ductus arteriosus | whooping cough |
| gastritis | peptic ulcer disease | yaws |
| gastroenteritis | periodontitis |  |

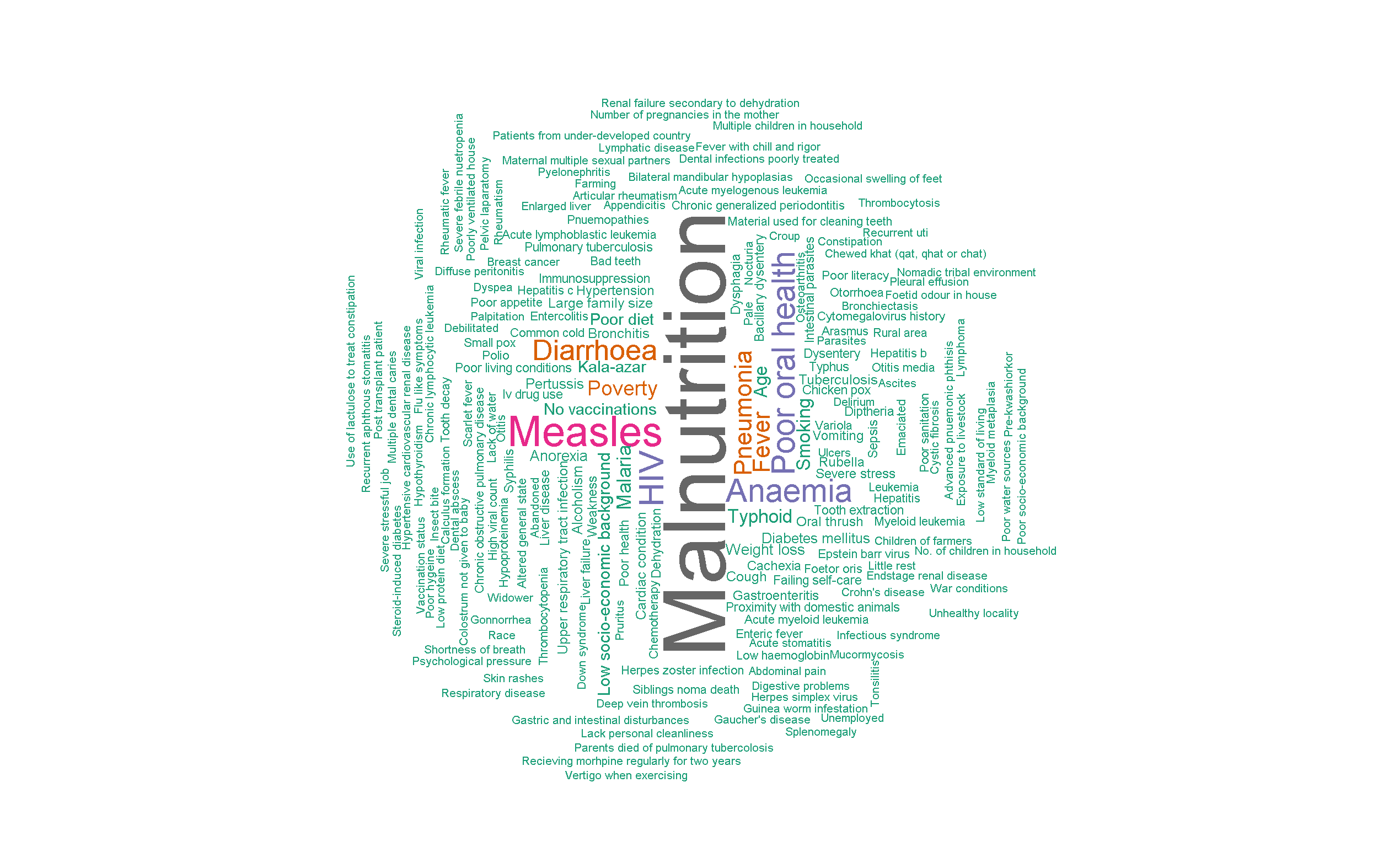

Figure 11	All risk factors as reported in the included publications

Size of the word is proportional to the number of times this word or string of words is reported across multiple publications.

##### a. Summary by risk factor category

*Nutritional deficit:* Nutritional deficit was identified as a risk factor in 142 (38.8%) studies, not identified as a risk factor in 36 (9.8%) and there was no report of nutritional status in the remaining 188 (51.4%) studies. 52.7% of studies (142/269) reporting risk factors observed some aspect of nutritional deficit (marasmic-kwashiorkor, stunting, wasting). Overall, 9.3% (25/269) of these studies reported a nutritional deficit in the context of HIV.

*Immunosuppression:* Overall, 22.7% of the studies (61/269) of those reporting risk factors observed some form of immunosuppression due to non-infectious comorbidity(ies). Cancer was reported in 13 of the 61 studies. Leukaemia was the most commonly reported form of cancer in 12 studies, with 67% of cases occurring in adult patients (8 out of 12).

*Infectious diseases comorbidities* as a possible risk factor for noma were detailed in 56.5% of the publications (152/269). Measles was the most frequently named disease, accounting for 35.5% (54/152). Other infectious diseases included malaria, HIV, visceral leishmaniasis, syphilis, dysentery, viral infections (CMV, EBV), enteric fevers, meningitis, plague, gonorrhoea, herpes zoster, chlamydia, tuberculosis and polio.

*Respiratory infections* were recorded in 53 publications with unspecified pneumopathies as the most cited illness (30%).

*Poor oral hygiene* was reported in 27.8% of the publications (75/269). Identifying poor oral hygiene as a risk factor posed some challenges due to its similarity with poor oral health as a consequence of noma. Nonetheless, the risk factor of poor oral hygiene was identified in 75 studies, with the years of publication ranging from 1841 to 2022. As with the year of publication, the country reported for studies identifying poor oral hygiene, varied widely. The countries reported were: Afghanistan, Bangladesh, Brazil, Burkina Faso, Colombia, England, France, Gambia, Ghana, India, Israel, Italy, Kenya, Micronesia, Morocco, Nepal, Niger, Nigeria, Pakistan, Senegal, South Africa, South Korea, Turkey, Uganda, UK, USA, Vietnam, Zambia, and Zimbabwe.

*Physical environment factors* were defined as close proximity to livestock, scant sanitation and unsafe drinking water. Only 39 publications detailed these risk factors in 1841 patients of whom 1,797 were from Africa. While a majority (58%, 23/39) of the studies reported were from Africa, a small proportion (8/39, 20.5%) was reported from the UK and USA during a period spanning from 1841 to 1893.

*Socio-economic factors* were only detailed in 24.2% of the publications (65/269) despite the common knowledge that noma manifests itself mainly in populations enclosed by extreme poverty. However, other risk factors such as lack of safe drinking water, limited access to good health services and malnutrition could possibly serve as proxy of poverty.

*Lifestyle factors* such as smoking, alcoholism, and intravenous drug use were captured in 41 publications (737 patients) with the 51% of the studies referring to adult patients (21/41).

##### b. Publications reporting risk factors per year

*Publications before 1950:* A total of 24.5% of the studies (66/269) of those reporting risk factors were published before 1950. More than half during this time period were from the UK (n=25) and USA (n=16) contrasting with a small number from Africa (n=4). Infectious diseases constituted the main risk factor category accounting for 75% (50/66), measles being the most frequently reported illness. Other risk factor categories included nutritional deficit and respiratory infections accounting for 28.8% (19/66) and 30.3% (20/66) respectively.

*Publications between 1950 and 2000:* Of the 269 studies reporting risk factors, 28.3% (76/269) were published between 1950-2000. Of these, 48.7% (37/76) were from Africa. The main risk factor categories reported are nutritional deficit (63.2%, n=48/76), infectious diseases (50.0%, n=38/76) and immunosuppression (23.7%, n=18/76).

*Publications after 2000*: 47.2% of the studies (127/269) that reported risk factors were published after 2000 with 56.0% originating from Africa (71/127). Nutritional deficit, immunosuppression and infectious diseases remained the main risk factors categories accounting for 59.1%, 38.6% and 40.1% respectively (75/127, 49/127, 51/127). Measles constituted the main risk factor reported in the infectious diseases category across the three periods (18 before 1950, 18 between 1951-2000, and 17 from 2000 onwards). Unsurprisingly, HIV as a risk factor was most reported after 2000 (in 32 out of 154 publications during this period) in comparison to 6 out of 96 publications from 1950 to 2000.

Although additional synthesis of risk factors and sub-group analyses were initially intended, the heterogeneity of data prevented further analysis.

### 4. Microbiology

Of the 366 included studies, 113 (30.9%) publications reported varying level of details on the microbiology of the noma cases. Of the 113 studies reporting microbiological agents associated with noma (both primary and secondary reports), 78 (69.0%) reported identifying multiple microorganisms, 20 (17.6%) reported a single microorganism and 15 (13.3%) did not list any specific organism (Table 8).

Table 8	Reporting of microbiological agents associated with noma in the included studies

|  | **Number of studies** |
| --- | --- |
| Single microorganism reported | 20 (16.8%) |
| Multiple microorganisms reported | 78 (72.6%) |
| No specific organism reported | 15 (10.6%) |
| Total number of studies reporting microbiology | 113 |

Note: There was substantial heterogeneity in the culture techniques associated with this reporting of all microbiology

Table 9 provides an exhaustive list of all microorganisms for noma which had been cultured through the blood and lesion smears. Each microorganism is listed as it directly appears in the publications.

Table 9	List of reported microorganisms from included noma studies

| staphylococcus aureus | Peptostreptococcus Genus | Bacilli species |
| --- | --- | --- |
| enterobacter aerogenes | Actinomyces Species | Cocci species |
| candida species | Streptococcus Pyogens | Prevotella species |
| Pseudomonas Aeruginosa | Streptococcus Viridins | enterococcus species |
| fusiform bacillus | Streptococcus Pneumoniae | Group B hemolytic streptococcus |
| fusobacterium necrophorum | Streptococcus Species | gram positive streptococci |
| Klebsiella Pneumoniae | Staphylococccus Albus | gram negative bacilli |
| Spirochetes | Group D Streptococcus | staphylococcus species |
| Escherichia Coli | Spirochaeta Vincenti | coliforms species |
| Haemophilus Influenzae | Streptococcus Salivarius | proteus |
| Pervotella Intermedia | Borrelia Vincenti | Veillonella parvula |
| Beta-Hemolytic Streptococci | Neisseria species | Clostridiales |
| Prevotella Melaninogenica | Enterobacter species | Nitrospina |
| Alpha-Hemolytic Streptococci | Bacteroides | Haliangium |
| staphylococcus aureus | Fusiform species | Saprospiraceae |
| spirochaeta dentium | Enterobacter Agglomerans | Streptococcus sanguinis |
| Proteus Mirabilis | Bacterium antatum | nonhemolytic streptococci |
| Acinetobacter baumanni | Candida athicans | diphtheroid bacilli |
| Staphylococcus epidermis | Providencia stuartii | fusiform spirilla |
| Citrobacter koseri | Vincent's organisms | Klebs-Loeffler organisms |
| Bacteroides fragilis | Streptococcus Faecalis | Streptococcus Group G |
| Bacillus cereus | serratia marcescens | Anaerococcus prevotii |
| Streptococcus sanguinis | citrobacter species | Leptotrichia Bucalis |
| nonhemolytic streptococci | Lactobacillus species | spirochaeta dentium |
| diphtheroid bacilli | Streptococcus Bovis | Streptococcus sanguinis |
| fusiform spirilla | capnocytophaga | nonhemolytic streptococci |
| Klebs-Loeffler organisms | Corynebacterium diptheriae | diphtheroid bacilli |
| Streptococcus Group G | Enterobacter Agglomerans | fusiform spirilla |
| Anaerococcus prevotii | Bacterium antatum | Klebs-Loeffler organisms |
| Leptotrichia Bucalis | Candida athicans | Streptococcus Group G |
| spirochaeta dentium | Providencia stuartii | Bacillus cereus |
| Proteus Mirabilis | Vincent's organisms | Streptococcus Bovis |
| Acinetobacter baumanni | Streptococcus Faecalis | capnocytophaga |
| Staphylococcus epidermis | serratia marcescens | Streptococcus Bovis |
| Citrobacter koseri | citrobacter species | Lactobacillus species |
| Bacteroides fragilis | Rhinovirus | Enterovirus |
| Streptococcus intermedius | vancomycin-resistance enterococci (VRE) | Escherichia coli Extended Spectrum Beta Lactamase. |
| Mycoplasma Pneumoniae | Neisseria Cattarhalis | Treponema microdentium |
| Fusobacterium nucleatum | Porphyromonas Species | Micrococcus species |

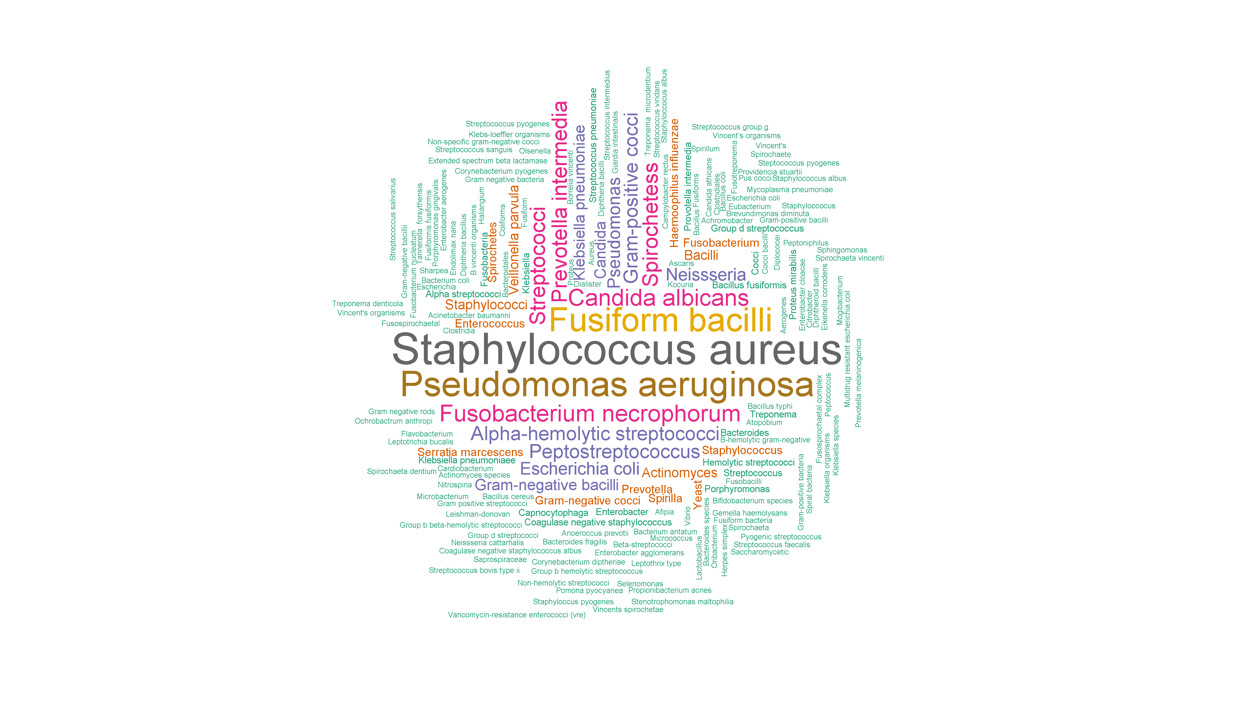

Figure 12	Frequency of microorganisms reported in the included publications

Size of the word is proportional to the number of times this word or string of words is reported across multiple publications.

*Microbiology by region:*

To assess the difference in observed microorganisms by region, publications reporting microbiology prior to 1990 were excluded from further analysis. Of the 67 studies conducted in the last 30 years, 5 studies reported microbiology from patients in the Americas, 21 from Asia, 13 from Europe and 28 studies from Africa. During this time period, the most common organisms reported from Europe were *Candida albicans* documented in 5 studies and *Streptococcus* species reported in 4 studies. The most common organism observed from the Americas was *Prevotella* species reported in 2 studies. The most common organisms identified from Asia were *Klebsiella pneumoniae* reported in 4 studies and *Candida albicans,* *Pseudomonas aeruginosa, Fusobacterium and Prevotella* species each documented in 3 studies. The most common organisms reported from Africa were *Staphylococcus* species listed in 7 studies, *Streptococcus* species and *Prevotella* species reported in 6 studies each, *Pseudomonas aeruginosa* reported in 5 studies, and *Fusobacterium* species reported in 4 studies. Among the publications from countries within the region of Africa known as the noma belt which reported any data on microorganisms (Burkina Faso, Ethiopia, Ghana, Mali, Niger, Nigeria, and Senegal) the most common bacteria reported from these areas were *Fusobacterium necrophorum, Klebsiella pneumonia, Prevotella intermedia,* and *Staphylococcus aureus*.

*Evidence of antimicrobial resistance:*

More recent case studies have reported cultures of resistant organisms. A single case from Afghanistan reported multidrug resistant *Escherichia coli* prior to any use of antibiotics (Barrera *et al.,* 2012). A case in South Korea reported a single microorganism, vancomycin-resistance enterococci (VRE) (Park *et al.,* 2022). Cultures from another case in Mali identified a polymicrobial infection and among the bacteria isolated an Extended Spectrum Beta Lactamase *Escherichia coli* (Traore *et al.,* 2021).

Given the number of organisms, heterogeneity of reporting and limited information on culture techniques, further comparison, sub-group analysis or examination of microorganisms at the individual patient level was not possible.

### 5. Prevention

This review found little evidence on noma prevention. Good nutrition practices, promotion of exclusive breastfeeding for the first three to six months of life, immunisation against endemic communicable diseases, oral hygiene practices, separation of livestock from human living areas, and noma education were recommended (Enwonwu *et al.,* 2006). Others suggested early detection and antibiotic treatment of intra-oral necrotising disorders may prevent noma, however no included research tested this hypothesis (Feller *et al.,* 2012). While “recommended interventions” are likely based on suspected noma risk factors, there is no solid evidence supporting them in the literature and they appear to be based on general health best practices.

### 7. Treatment

The table below provides a deduplicated and exhaustive list of all treatments for noma (medical, surgical, traditional/local practices) associated with reported cases or the noma study populations in the included studies. Each treatment is listed as it directly appears in the publications. For the frequency of reported treatments see Figure 13.

Table 10	List of reported treatment from included noma studies

| sodium ricinoleate solution | dental wax mould | one stage lip reconstruction with Gillies fan flap |
| --- | --- | --- |
| "Enzymol" plug | dentoline cautery | opiates |
| 11 fold cheiloplasty | denture | oral hygiene prophylaxis |
| 16-fold coral reconstruction | dequalinium chloride | osteotomy |
| Abbe flaps | Dextro propoxyphene hydrochloride (Darvon) | palliative treatment |
| acetate of ammonia | dextrose | paracodin |
| acriflavine | diazepam | parascapular free flap |
| acyclovir | diptheria antitoxin | parenteral rehydration |
| adriamycin | Dover's powder | partial flap necrosis |
| aggressive fluid therapy | drainage of abscess | pedical flaps |
| albumin | electrolytic chlorine | Pedicled latissimus dorsi myocutaneous flap |
| alphachymotrypsin | electrolytic re-equilibration | pedicled supraclavicular island flap |
| amikacin | erythromycin | penicillin |
| aminoglycosides | escharotics | penicillin G |
| amoxicillin | Estlander flap | perchloride of murcury |
| ampicillin | Eusol solution | Peridex |
| angle ostectomy | excision of fibrous tissue | physiotherapy |
| antibiotic therapy | excision of parts of internal pterygoid muscle | piperacillin/tazobactam |
| antifungals | excision of parts of the masseter muscle | plastic surgery |
| antispirill vaccine | fermenting poultice | potassium chlorate |
| antiviral therapy | fibre optic assisted intubation | potassium permanganate solution |
| appendectomy | flap prefabrication | potassium sulphate |
| aqua rosae | flap trimming | potassium suplement |
| aqueous penicillin | flavine solution | povidone iodine |
| arsenic treatment | flucanozole | powdered rhubarb |
| arsenicaux | flucloxacillin | prefabricated superficial fascia flaps |
| ascorbic acid | fluconazole | Pregamal |
| augmentin | free flap | prelamination |
| aureomycin | free serratus anterior flap | Prontosil album |
| beef tea | frontotemporal flap | proper nutrition |
| benzoyl peroxide powder | gargarisma Condi | purinethol |
| benzydamine hydrochloride spray | gargle of carbolate of glycerine | quick cutting intubation |
| benzylpenicillin | gargle with solution of chloride of lime | quinine |
| beta lactums | genioplasty | realimentation |
| biborate of soda | gentalline | reconstructive surgery |
| bichloride of murcury | gentian violet | rectal paracetamol |
| bilatcostochondral flap | glucose | rehabilitiation with prosthesis |
| bilateral buccal scar lysis | glycerium acidi tannici | rehydration |
| bipedicled chin flap | gum paint | release of bony ankylosis |
| bipenicillin | hemotherapy | removal of gangritis stomatitis |
| bismarsen | heparin | removal of necrotic bone |
| bismuth | hepatic extracts | removal of slough |
| bismuth pastes | hydrargum cum creta | renal replacement therapy |
| Bisthmus iodoform paraffin paste (BIPP) on gauze | hydrocortisone | resection of affected area |
| blind intubation | hydrogen peroxide | restoration of oral cavity |
| blood transfusion | imidazole | rhubarb |
| bone bridge excision | imipenem | saline |
| bone graft | indigenous treatment | salvarsen |
| bone reconstruction | infliximab | scalp flap |
| bony sequestrectomy | intrathecal methotrexate | scammony powder |
| boric acid solution | iodoform | Seldinger minitracheostomy |
| boric compress | iron sulphate | sequestrectomies |
| brandy | iron with bark | skeletal restoration |
| broad spectrum antibiotics | irrigation of peritoneal cavity | skin graft |
| bromine | jalap powder | sloughectomy |
| butter of antimony | kaolin | sodium perborate |
| calcium gluconate | leeches | soft tissue reconstruction |
| calomel | lidocaine gel | soluceptacin |
| carbenicillin | lincocin | spirometry |
| carbolic dressing | linseed poultice | starch and opium enema |
| carbonate of ammonia | liquor of sarsae | steeple flap |
| carbonate of ammonia | lockjaw treatment | streptomagma |
| cartilage graft | lycomicin | streptomycin |
| castor oil | macrolidi | submental intubation |
| Catasplana Lini topically | mandibular ostectomy | sulfanilamide |
| cautery | mapharsen | sulfathiozol |
| cedilanid | mecanotherapy | sulphapyridine |
| cefalosporin | mercury | sulphate of quina |
| ceftazidime | meropenem | sulphonamide |
| cefuroxime | methicillin | sulphur |
| cephalexin monohydrate | methotrexate | sultirene |
| cervical rotation flap | methyl-glucamine | suphale of zinc lotion |
| cheek rotation flap | metrogyl | supraclavicular tubed flap |
| chemotherapy | metronidazole | syrup of ginger |
| chinese herbal covering | microsurgical reconstruction | temporalis muscle and temporalis fascia flaps |
| chloramphenicol | microvascular forearm flap | terramycin |
| chlorhexidine and hydrogen peroxide mouthwashes | Milton lotion | testosterone |
| chloride of zinc | mitoxantrone | tetanus antitioxin |
| clindamycin | mixture of chalk mixture and tincture of catechu and ticture of cinnamon | tetracycline |
| clindamycin | mixture of sulphocarbolate of sodium, sulphite of sodium with Decoctum Cinchona | Therapuetic B complex |
| clindamycinazathiprine | modified eggnog | three stage reconstruction |
| clomicine | morniflumate | three-fold oral opening and canthopexy |
| co-amoxiclav | morphia | thromboplastin |
| commissuroplasty. | morphine | tincture of aconite |
| commisuroplasty | morphine sulphate | tooth powder |
| Condy's fluid | moutache reconstruction | topical bacitracin |
| condylectomy | murcurial purgative | tracheostomy |
| contralateral coronoidectomy | murcurochrome | tube flap |
| convers flap | muriate iron tincture | tube graft |
| coronoid process excision | muriatic acid | turnover flap |
| coronoidectomy | muriatic beef juice | turpentine stupes |
| cortancyl | musculocutaneous latissimus dorsi flap | tyrothrycin |
| corticosteroids | myocutaneous flaps | ultra-violet radiation |
| cough mixture | nasal reconstruction | urea stibamine |
| cranial flaps | nasogenal flap technique | vancomycin |
| crude live extract | near-total conservative right parotidectomy | Vascularized calvarium flap |
| cyclophosphamide | neoarsphenamin | vestibuloplasty |
| cytarabine | neostibosan | vincristine |
| cytoxan | netilmicin | vitamin supplementation |
| daily rectal irrigations | nitrate of silver | waltzing flap |
| Dakin Cooper | nitric acid locally | warfarin |
| debridement of wound | nutritous enema | Webster flap |
| dehydration therapy | nux vomica | whisky |
| deltopectoral flap | nystatin | wine |
| demerol | ofloxacin | yeast |
| dental avulsions | oncovin |  |

NB: It is important to remember here, as a warning from history, that noma research of extraordinary unethical barbarity was conducted, in World War 2 led by Mengele at Auschwitz Birkernau on Sinti and Roma extermination camp prisoners, with treatment with sulphanilamido-ethyl thiodiazole and nicotinic acid. These 'data' were never published (Halioua & Halioua 2021).

Table 11	Publications and patient numbers reported by noma treatment modality received

| **Treatment modality** | **Number of publications** | **Number of patients** |
| --- | --- | --- |
| Medical | 108 | 654 (4.3%) |
| Surgical | 94 | 1,554 (10.3%) |
| Both medical and surgical | 119 | 1,999 (13.3%) |
| Neither medical nor surgical | 31 | 2,665 (17.7%) |
| Unspecified^*^ | 14 | 8,210 (54.4%) |
| **Overall** | **366** | **15,082** |

* Treatment or intervention of any kind was not reported in the publication

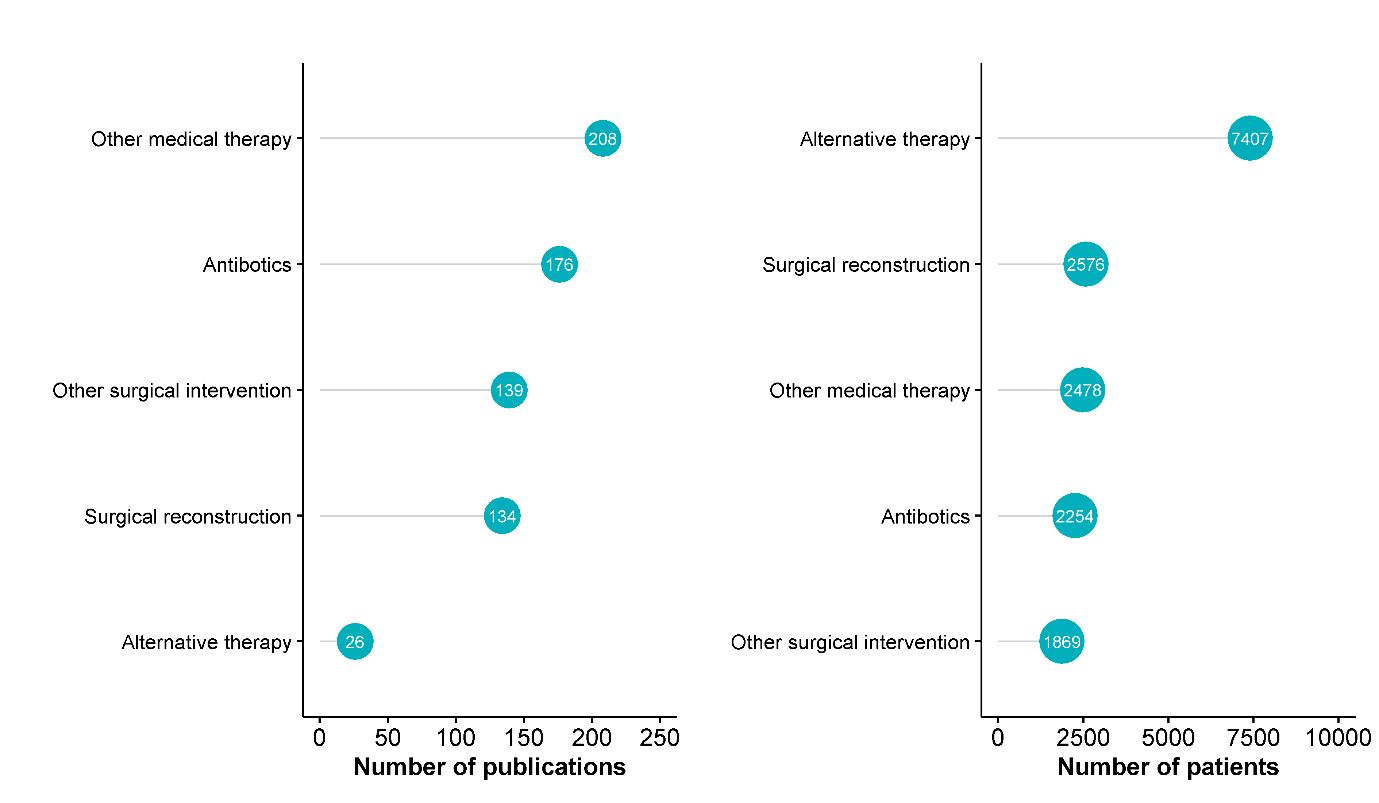

Figure 13	Number of studies and patients reported to have received treatment for noma within each intervention category

Note: the intervention categories were designated for the purposes of this systematic review; each study can have more than one intervention and hence the number of publications will add to >100%.

Definitions: ‘Other surgical intervention’ = any surgical procedure that was not reconstruction (e.g., wound management during the acute phase, wound debridement); ‘Other medical therapy’ = any medical intervention that was not surgical or the administration of antibiotics; ‘Alternative therapy’ = non-medical or surgical intervention (e.g., complementary medicines, nutritional supplements, traditional medicine and practices). See SF2_VariableDictionary.

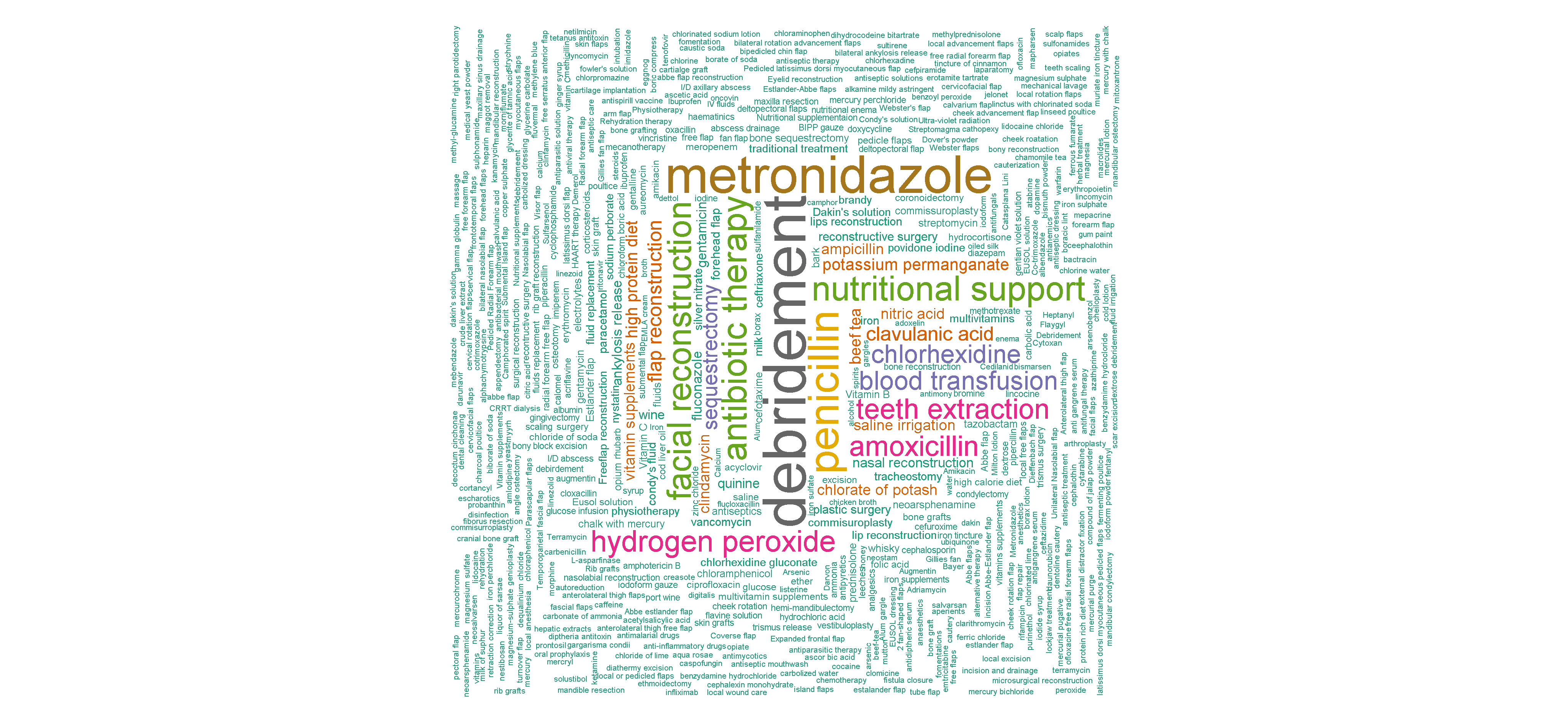

Figure 14	Frequency of treatments reported in the included publications

Size of the word is proportional to the number of times this word or string of words is reported across multiple publications.

##### a. Study design & time period

For each study design, the number of patients and studies by the additional sub-group categorisation of intervention type (antibiotic, other medical therapy, alternative therapy, surgical reconstruction, other surgical procedure) in Table 13. Specific observations by study design are reported below:

*Case Reports:*

The included studies which reported interventions for noma were predominantly case reports or series (270 studies) which included 884 patients and of these, the use of a combination of surgical and medical interventions was the most common course of management for the noma patients with 103 case reports/series with 244 patients. Thirty five percent of the studies (95/270) reported the use of medical treatment only and 23% of the publications (63/270) reported the use of surgical treatment only for noma patient management with 188 patients and 426 patients respectively.

When observing the sub-categories of intervention type, the predominant form of noma therapy as reported by case reports was other medical therapies (182 case reports/series representing 423 patients). These other medical therapies (i.e., not antibiotics) included opiates, tincture of chloride, topical silver nitrate, topical zinc peroxide, vitamin therapy and blood transfusion. The case reports reporting treatment of noma with antibiotics, where antibiotics were prescribed to 381 patients, 456 patients reported surgical reconstruction, 377 patients reported other surgical procedures and 25 patients received an alternative therapy.

A number of case reports (Chaubal et al. 2017, Malek et al. 2017, Murrell et al. 2010, Feller et al. 2012, Fetterolf et al. 2015, Umeizudike et al. 2011, Hu et al. 2015, Kato et al. 2017, Özberk et al. 2018, Martos et al.2019, Maccarrone et al. 2019) have shown that early intervention, during the acute necrotizing gingivitis stage, with proper oral hygiene, debridement, antibiotics therapy and chlorhexidine mouthwash leads to proper wound healing and no long-term adverse effects. This shows that early diagnosis would be the best route to preventing more severe stage progression.

Table 12	Number of studies and patients by intervention and study design

|  | **Study design** | | | | | | | | | |
| --- | --- | --- | --- | --- | --- | --- | --- | --- | --- | --- |
| **Intervention** | **Case series/reports** | | **Cohort** | | **Cross-sectional** | | **Interventional** | | **Other** | |
|  | No. of  case series/ reports | No. of  patients | No. of cohort studies | No. of  patients | No. of cross sectional  studies | No. of  patients | No. of interventional studies | No. of  patients | No. of  studies | No. of  patients |
| Medical | 95 | 188 | 6 | 262 | 4 | 157 | 2 | 39 | 1 | 8 |
| Surgical | 63 | 426 | 23 | 822 | 3 | 179 | 4 | 62 | 1 | 65 |
| Both | 103 | 244 | 13 | 934 | 3 | 821 | - | - | - | - |
| Neither | 5 | 21 | 7 | 616 | 15 | 1,814 | - | - | 4 | 214 |
| Unspecified* | 4 | 5 | 4 | 254 | 4 | 7,630 | - | - | 2 | 321 |
| **Total** | 270 | 884 | 53 | 2,888 | 29 | 10,601 | 6 | 101 | 8 | 608 |

Both = Medical and surgical, Neither = Neither Surgical nor medical.

* Treatment or intervention of any kind was not reported in the publication

Table 13	Number of studies and patients who received noma interventions by sub-group for each study design

|  | **Study design** | | | | | | | | | |
| --- | --- | --- | --- | --- | --- | --- | --- | --- | --- | --- |
| **Sub-group of  intervention type** | **Case series/reports** | | **cohort studies** | | **cross-sectional** | | **Interventional** | | **Other** | |
|  | No. of  case series/ reports | No. of  patients | No. of cohort studies | No. of  patients | No. of cross sectional  studies | No. of  patients | No. of interventional studies | No. of  patients | No. of  studies | No. of  patients |
| Antibiotics | 150 | 381 | 15 | 881 | 6 | 921 | 4 | 63 | 1 | 8 |
| Other medical therapy | 182 | 423 | 19 | 1177 | 4 | 828 | 3 | 50 | - | - |
| Alternative therapy | 21 | 25 | 2 | 71 | 3 | 7311 | - | - | - | - |
| Surgical Reconstruction | 94 | 456 | 31 | 1374 | 4 | 619 | 4 | 62 | 1 | 65 |
| Other surgical Procedure | 118 | 377 | 16 | 694 | 2 | 748 | 3 | 50 | - | - |

*Overlap is attributed to studies which report more interventions from more than one category. These studies will be counted more than once.

Table 14	Noma management over different periods of time by intervention type

| Period | Medical | | Surgical | | Both | | Neither | | Unspecified* | |
| --- | --- | --- | --- | --- | --- | --- | --- | --- | --- | --- |
|  | No. of  studies | No. of  patients | No. of  studies | No. of  patients | No. of  studies | No. of  patients | No. of  studies | No. of  patients | No. of  studies | No. of  patients |
| Pre-1900 | 21 | 46 | 3 | 7 | 11 | 21 | - | - | 1 | 2 |
| 1900-1924 | 2 | 2 | 4 | 5 | 3 | 12 | - | - | 1 | 1 |
| 1925-1949 | 15 | 166 | 2 | 7 | 11 | 98 | 2 | 2 | - | - |
| 1950-1974 | 11 | 129 | 9 | 78 | 9 | 104 | 2 | 8 | 1 | 1 |
| 1975-1999 | 11 | 48 | 18 | 197 | 28 | 327 | 1 | 22 | 2 | 13 |
| 2000-2009 | 19 | 182 | 25 | 519 | 23 | 619 | 11 | 1,517 | 4 | 267 |
| 2010 onwards | 29 | 81 | 33 | 741 | 34 | 818 | 15 | 1,116 | 5 | 7,926 |
| Overall | 108 | 654 | 94 | 1,554 | 119 | 1,999 | 31 | 2,665 | 14 | 8,210 |

Both = Medical and surgical, Neither = Neither Surgical or medical.

* Treatment or intervention of any kind was not reported in the publication

Table 15	Noma management over different periods of time by sub-group of intervention type

| Period | Antibiotic | | Other medical therapy | | Alternative therapy | | Surgical Reconstruction | | Other surgical procedure | |
| --- | --- | --- | --- | --- | --- | --- | --- | --- | --- | --- |
|  | No. of  studies | No. of  patients | No. of  studies | No. of  patients | No. of  studies | No. of  patients | No. of  studies | No. of  patients | No. of  studies | No. of  patients |
| Pre-1900 | - | - | 31 | 60 | 9 | 13 | 3 | 5 | 13 | 30 |
| 1900-1924 | - | - | 6 | 11 | 1 | 1 | 2 | 3 | 8 | 18 |
| 1925-1949 | 11 | 51 | 27 | 258 | 1 | 57 | 2 | 6 | 16 | 111 |
| 1950-1974 | 21 | 239 | 17 | 167 | - | - | 13 | 107 | 9 | 81 |
| 1975-1999 | 41 | 301 | 38 | 377 | 1 | 1 | 32 | 499 | 28 | 236 |
| 2000-2009 | 41 | 713 | 34 | 681 | 7 | 134 | 34 | 731 | 26 | 634 |
| 2010 onwards | 62 | 950 | 55 | 924 | 7 | 7201 | 48 | 1225 | 39 | 759 |
| Overall | 176 | 2,254 | 208 | 2,478 | 26 | 7,407 | 134 | 2,576 | 139 | 1,869 |

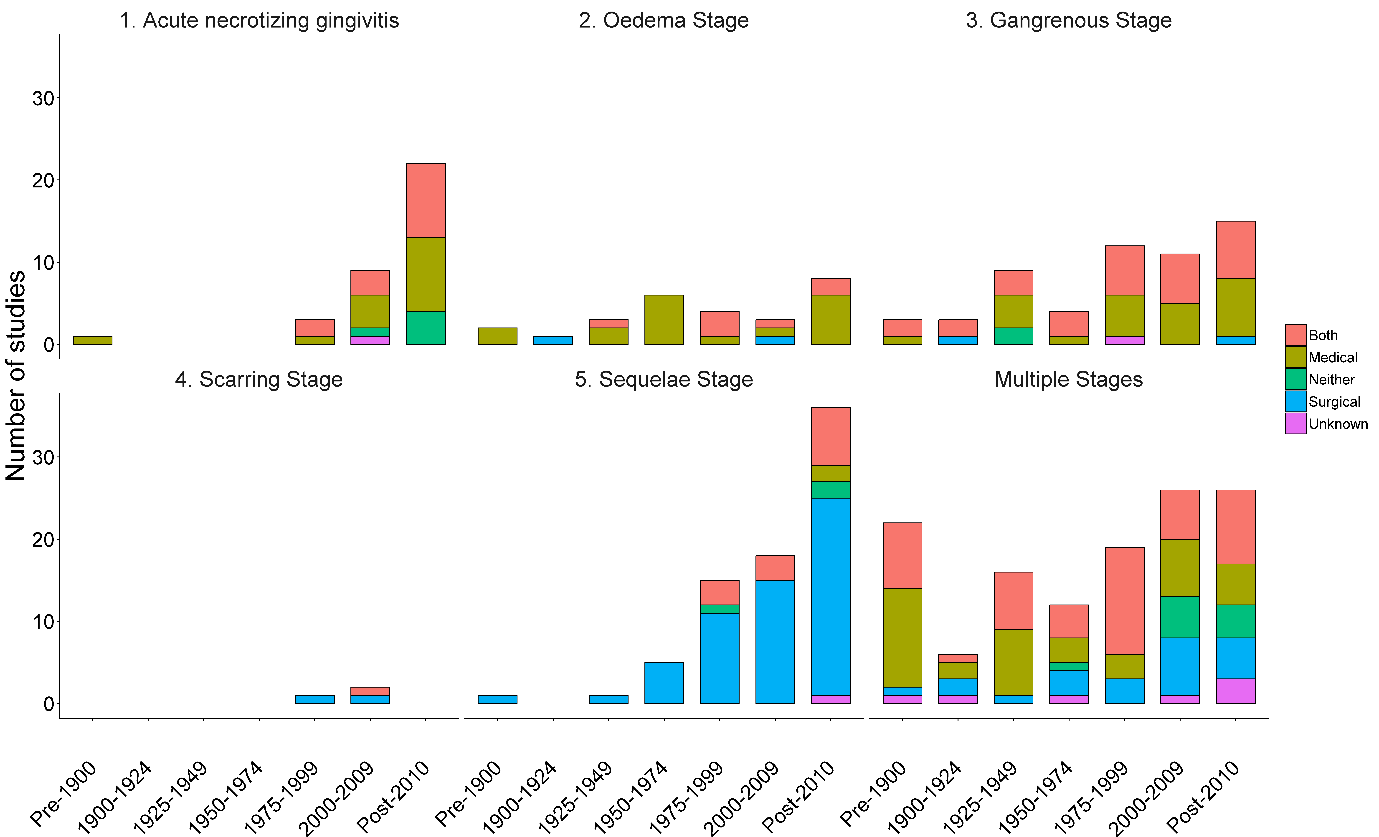

Figure 15	Noma studies and intervention type grouped by disease stage and date of publication

Note: This figure does not display the included noma publications for which the disease stage was unknown or unclear.

##### b. Disease Stage

Where possible the 366 included studies were categorised by noma disease stage. However, few studies explicitly reported the noma stage of included patients using the WHO definitions. While stage was judged by reviewers based on available information provided on symptoms and diagnosis, many did not provide sufficient information to allow an identification of noma stage. Furthermore, many publications reported on patients included at different stages of noma or detailed patients who progressed from one stage to another. Even more challenging in synthesising available information was the stage at which individuals received treatment, particularly if multiple modalities received for those patients observed progressing through multiple phases. Similar challenges were faced in observing outcomes and treatment modalities across patients within a single study who were recruited at different phases of the disease. Accordingly, the following results should be interpreted with caution.

For each noma stage, the number of patients and studies are reported by intervention type (medical, surgical, both, unspecified) in Table 16. This information is also presented by the additional treatment sub-categories (antibiotic, other medical therapy, alternative therapy, surgical reconstruction, other surgical procedure) in Table 17. Temporal evolution of treatment modalities and sub-interventions are presented in Figure 14, Figure 15 and Figure 16. Results of the observations of treatment modalities by noma stage are described below.

*Acute necrotising gingitvitis (ANG):*

Of the 35 ANG studies, 15 studies (n=65 patients) reported medical treatment, 2 studies (n=2 patients) reported surgical, 14 (n=25 patients) reported both medical and surgical, 5 (n=802 patients) reported no treatment or intervention, and in 1 study (n=68 patients) treatment was unspecified. When observing the sub-categories of intervention type, the most commonly reported therapy for this stage is other medical therapy (28 studies; 89 patients), which included vitamins, high-protein diets, blood transfusions, and oral rinsing.

Antibiotics (including ampicillin, metronidazole, amoxicillin, penicillin, and kanamycin) were administered in 26 studies (n=87 patients). The majority of the antibiotics usage in this stage occurred during the 2000-2009 period (See Figure 15). Other surgical procedures (including excision of necrotic areas, removal of loose teeth and bone, and debridement of the mouth) for management of the disease, were delivered in 17 studies (n=53 patients); most of these occurred after 2000 (Figure 15).

The final sub-category of intervention reported for patients presenting in the ANG stage was surgical reconstruction, in 2 case reports (n=2 patients). Barrios et al., 1995 reported a single case of noma in an HIV-coinfected patient who received surgical reconstruction in the form of tooth removal, and closure of the intraoral and extraoral defects, over 6 months after presenting with noma and receiving antibiotic therapy. The second case was reported by Evans et al in 2001 and included a man with the comorbidity of diabetes mellitus. This patient received oro-facial reconstruction following progressive destruction of his lips.

*Oedema stage:*

Overall, 27 studies reported this stage in 33 patients (Table 8). All the studies in the oedema stage reported details of interventions, with the majority of studies categorised as receiving medical treatment alone (18/27 studies; n=23 patients). Of the remaining studies, 2 (n=2 patients) reported only surgical interventions, and 7 studies (n=8 patients) reported a combination of both medical and surgical interventions.

The most commonly reported sub-category of intervention type for the oedema stage was, similar to the ANG stage, other medical therapies (23 studies, n=29 patients). The treatments reported within this category included sodium chloride gargles, boric compress, potassium chlorate gargles, diluted hydrogen chloride, high calorie diets, nutritional support, and blood transfusions. Antibiotics, among which were penicillin, lincomycin, streptomycin, metronidazole, flucloxacillin, ampicillin, and were administered in 22 studies (n=25 patients); there was a relatively limited number of studies reporting antibiotics usage between 1975-1999 (Figure 15). The use of alternative therapy was reported in 2/20 studies (n=2 patients), published in 1852 and 1923, and included the following treatments: leeches, mercurial purgative, wine, beef-tea, ammonia and bark.

In the oedema stage, surgical treatments were also administered, although most frequently in conjunction with medical therapy. Debridement of affected sites, drainage of abscesses, and the removal of necrosed tissue and bone, were among the other surgical interventions reported in 8 studies (n=10 patients). Surgical reconstruction occurred in 3 studies (n=3 patients). The first, Hefferman et al., 1923, reported a tube graft. The second, Daya and Nair, 2009, reported the performance of a bilateral mandibular condylectomy and vascular anastomosis. A third case report, Xu 2019, reported using continuous renal replacement therapy (CRRT) for septic shock arising from noma, following which at ten days after admission, surgery was performed to remove the scar crust and the necrotic tissue. No further details of the surgical techniques were reported.

*Gangrenous stage:*

The gangrenous stage was reported in 57 studies (n=94 patients). For this stage, no treatment was reported in 2 studies (n=2 patients), and undefined in a further 1 study (n=9 patients). Medical treatment only was reported in 23 studies (n=44 patients), with surgical treatment only reported in 2 studies (n=2 patients), and a combination of both medical and surgical treatment reported in 29 studies (n=37 patients).

Antibiotics were the most common treatment in this stage of the disease, with 44 studies (n=68 patients) reporting use of them. Antibiotic usage for this stage was reported between 1925-1950 and most of the studies reporting antibiotic usage for this stage were published after 2000 (Figure 15). The named antibiotics include penicillin, clindamycin, amoxicillin, aureomycin, gentamicin, cefuroxime, metronidazole, and oxacillin. Other medical therapies were administered in 41 studies (n=64 patients), and included clavulanic acid, mitoxantrone, antifungals, amphotericin-b, magnesium sulphate solution, neoarsphenamine, and nutritional support. Alternative therapy in the form of herbal therapy was administered in 2 studies (n=2 patients).

At this stage of noma, surgical treatment was used in support of medical therapy; 11 studies (n=15 patients) reported surgical reconstruction, and other surgical treatment was reported in 29 studies (n=38 patients). As with the previous phases, surgical management was primarily comprised of the excision of necrotic areas and wound debridement. The surgical reconstruction commonly entailed resection of the affected areas like the mandible and plastic reconstructive surgery. There were relatively few studies describing surgical reconstruction prior to the 1950s, whereas the usage of other surgical procedures have been adopted consistently across the time-period (Figure 15).

*Scarring stage:*

Of the WHO categories for noma, few were attributed to the scarring stage, which only formed approximately 0.8% of the included studies (3/366). These 3 studies were case reports, and all included information on the administered treatment.

Two studies reported receiving surgical interventions only. Both included surgical reconstruction (bipedicled chin flap for lip reconstruction, osteotomy, and flap reconstruction). While this was the sole intervention in one study, the other also reported mechanotherapy and massage which were categorised as alternative therapy.

The third case report in this category, Chidzonga and Mahomva, 2008, reported medical and surgical interventions. This report was the first documented case of recurrent noma in an HIV/AIDS patient, with surgical reconstruction. The patient initially received antibiotics, nutritional support, debridement of necrotic tissue, and wound irrigation and dressing before surgical reconstruction. However, the patient relapsed and was re-admitted approximately one year later when the same management plan was followed.

*Sequalae stage:*

The sequalae stage was reported in 76 studies in 1,120 patients. Surgical intervention was reported in 22 studies (n=530 patients), medical interventions in 40 studies (n=440 patients), and both medical and surgical interventions in 13 studies (n=50 patients). In one study, interventions were unspecified (n=163 patients) and 3 studies (n=75 patients) reported no information on intervention or treatment.

Of the intervention sub-categories, surgical reconstruction was most commonly reported in 65 studies (n=772 patients). There has been a steady increase in the number of studies reporting surgical reconstruction since the 1950s (Figure 14). A wide range of reconstructive surgery was performed including surgical release of ankylosis, lip reconstruction, and facial reconstruction. For the latter, a wide range of flap procedures were reported. Other surgical interventions including excision and cauterization were reported in 24 studies (n=290 patients), and commonly undertaken as a precursor for reconstructive surgery.

Antibiotic therapy, including augmentin, penicillin, and metronidazole, was reported in 17 studies (n=151 patients). Other medical therapies, including plasma infusions and nutritional support, were reported in 20 (n=135 patients) sequelae stage studies. Alternative healing was reported in 6 studies (n=6 patients), and included a Chinese herbal covering, indigenous treatment, and treatment at noma onset by a traditional health practitioner.

*Multiple stages:*

Overall, 127 studies were assigned to multiple stages and included the majority of the patients identified in this review (n=10,981 patients). The majority of studies in this category were published after 2010 (See Figure 15). No treatment was reported in 10 studies (n=998 patients), and treatment was mentioned but undefined in 7 studies (n=7,496 patients). Of the remaining studies, 10 (n=80 patients) reported medical interventions, 9 (n=188 patients) reported surgical interventions, and 48 (n=1,517 patients) reported both intervention types.

Within the sub-categories, other medical interventions were reported in 80 studies (n=1,800 patients), antibiotics were reported in 54 studies (n=1,500 patients), and 14 studies (n=7,384 patients) reported alternative therapy. Other surgical procedures were described in 50 studies (n=1,139 patients), surgical reconstruction was described in 39 studies (n=1,489 patients).

Table 16	Noma management by disease stage and intervention type

| **Noma Stage** | **Type of Intervention** | | | | | | | | | |
| --- | --- | --- | --- | --- | --- | --- | --- | --- | --- | --- |
|  | **Medical** | | **Surgical** | | **Both** | | **Neither** | | **Unspecified** | |
|  | No. of studies | No. of cases | No. of studies | No. of cases | No. of studies | No. of cases | No. of studies | No. of cases | No. of studies | No. of cases |
| 1. Acute necrotizing gingivitis | 15 | 65 | 2 | 2 | 14 | 25 | 5 | 802 | 1 | 68 |
| 2. Oedema Stage | 18 | 23 | 2 | 2 | 7 | 8 |  |  |  |  |
| 3. Gangrenous Stage | 23 | 44 | 2 | 2 | 29 | 37 | 2 | 2 | 1 | 9 |
| 4. Scarring Stage | 2 | 2 | 57 | 830 | 1 | 1 |  |  |  |  |
| 5. Sequelae Stage | 40 | 440 | 22 | 530 | 13 | 50 | 3 | 75 | 1 | 163 |
| Multiple Stages | 10 | 80 | 9 | 188 | 48 | 1,517 | 10 | 998 | 7 | 7,496 |
| Unclear |  |  |  |  | 7 | 361 | 11 | 788 | 4 | 474 |

Both = Medical and surgical, Neither = Neither Surgical or medical.

* Treatment or intervention of any kind was not reported in the publication

Table 17	Noma management by disease stage and sub-group of intervention type

| **Noma Stage** | **Sub-group of intervention type** | | | | | | | | | |
| --- | --- | --- | --- | --- | --- | --- | --- | --- | --- | --- |
|  | **Antibiotics** | | **Other medical therapy** | | **Alternative therapy** | | **Surgical reconstruction** | | **Other surgical procedure** | |
|  | Number of studies | Number of patients | Number of studies | Number of patients | Number of studies | Number of patients | Number of studies | Number of patients | Number of studies | Number of patients |
| 1. Acute necrotizing gingivitis | 26 | 87 | 28 | 89 | - | - | 2 | 2 | 17 | 53 |
| 2. Oedema Stage | 22 | 25 | 23 | 29 | 2 | 2 | 3 | 3 | 8 | 10 |
| 3. Gangrenous Stage | 44 | 68 | 41 | 64 | 2 | 2 | 11 | 15 | 29 | 38 |
| 4. Scarring Stage | 1 | 1 | 1 | 1 | 1 | 1 | 3 | 3 | 2 | 2 |
| 5. Sequelae Stage | 17 | 151 | 20 | 135 | 6 | 6 | 65 | 772 | 24 | 290 |
| Multiple Stages | 54 | 1,500 | 80 | 1,800 | 14 | 7,384 | 39 | 1,489 | 50 | 1,139 |
| Unclear | 12 | 422 | 15 | 360 | 1 | 12 | 11 | 292 | 9 | 337 |

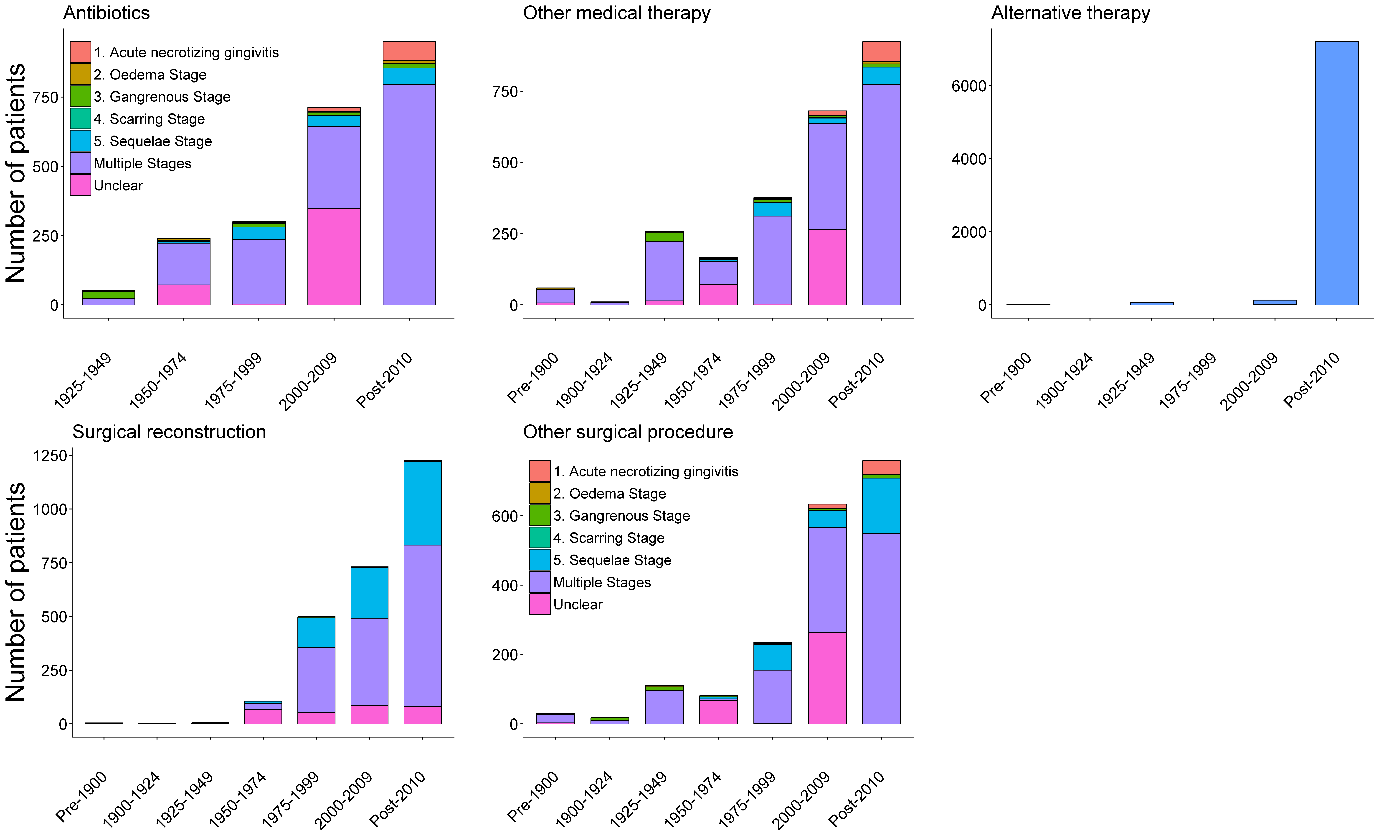

Figure 16	Noma management by disease stage and sub-group of intervention type (Number of patients)

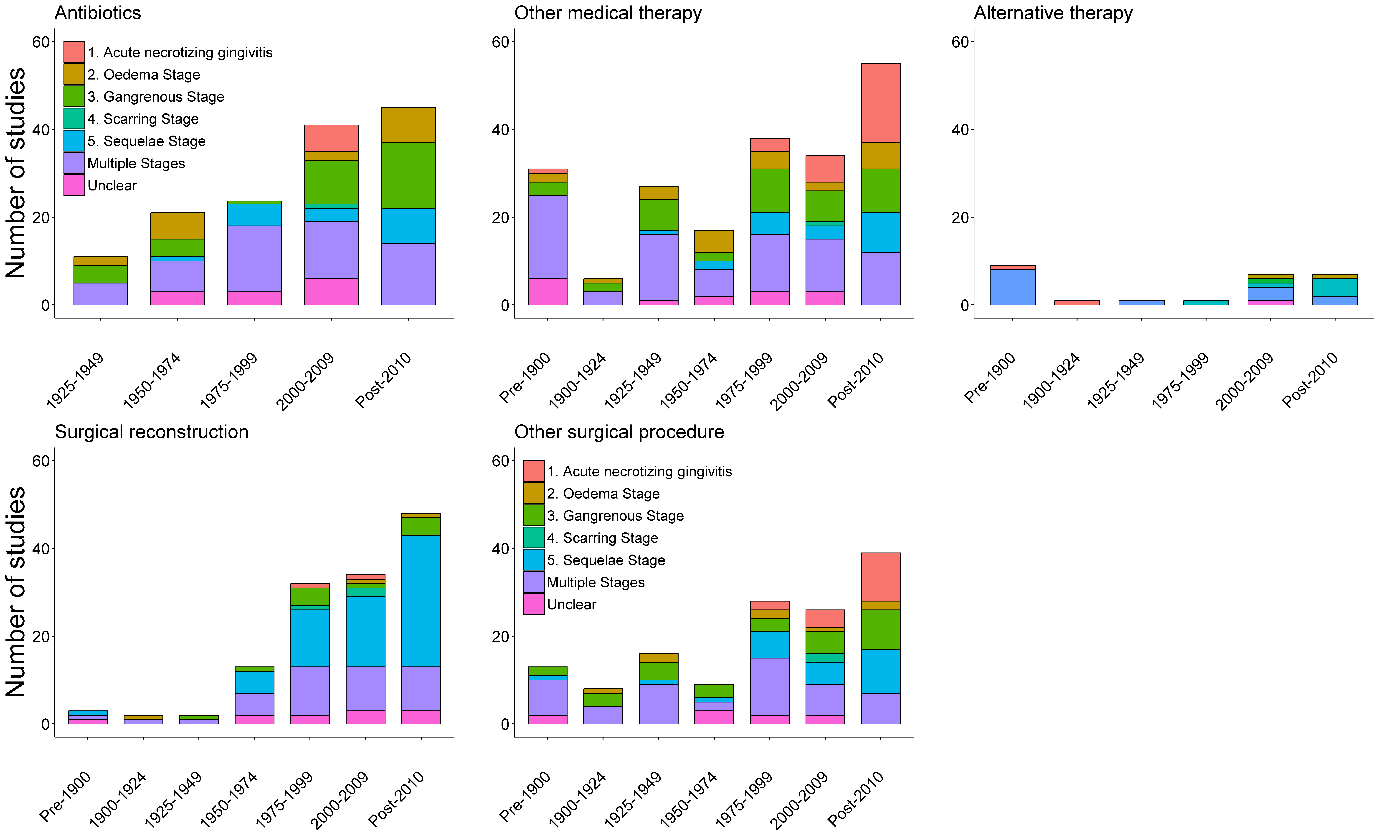

Figure 17	Noma management by disease stage and sub-group of intervention type (Number of studies)

### 8. Deaths

***Disclaimer:***

*Most noma experts report that only 10% (although ranges between <5% to 30%) of acute noma patients access medical care, due to their remote location, extreme poverty, and lack of free medical care. In the literature, it is often estimated that there is between 70 to 90% untreated mortality, which contributes significantly to the currently reported disease burden (WHO 1998; Barmes et al., 1997). Therefore, the included literature in this review reflects the small proportion of noma patients who attend healthcare facilities and are subsequently reported, and is hardly representative of all noma cases. Necessarily, mortality rate cannot be appropriately extrapolated and generalised from this systematic review.*

*However, death and post-noma complications (as broadly categorised further below) were the only treatment outcomes that could be extracted in a consistent way across the heterogeneous literature included in this review.*

***To provide a baseline of reported information on patient outcomes in the current evidence base, results and details of the deaths and groups of post-noma complications that are described in the literature are provided for comprehensiveness below. As detailed further in the discussion there are serious limitations to any interpretations of reported deaths and the representativeness and applicability of mortality.***

Among the 366 included studies, 283 (77.3%) reported information on mortality. This included 187 studies where it was explicitly reported that there were no deaths in noma patients. Across the 283 publications which reported mortality and included 4,181 noma participants, a total of 291 deaths were reported. On examination of the reported deaths, 88 (2.1%) of all included patients (representative of only those who attended a health facility/had access to medical attention) had died due to the acute effects of noma, which was determined on the basis that these patients died shortly after admission into a health facility. The remaining 203 specified that the cause of death was not related to noma or no clear cause was reported.

The age-wise distribution of deaths as documented in Table 19 were as follows; 3 deaths among infants (<1 year), 116 deaths among children (1-10 years), 35 deaths among adults (>19 years) with no death reported in the adolescent (10-19 years) age group. An additional age category for children not specified in the publications by the contiguous groups was the 0 to 19 year olds. There were 18 deaths in the 0-19 years age category. Publications which included patients falling under multiple of the aforementioned age groups were categorised under “all ages” with 90 deaths occurring in this group. Age of noma patients was not reported for 8 deaths.

Death was a common outcome of noma prior to the discovery of antibiotics in 1929, although widespread use and availability of antibiotics was limited until the late 1940s. Reported deaths in the included studies was therefore examined over a number of time periods as seen in Table 18. There were 47 studies in the first group with 95 patients in total, among which 48 deaths were reported. The second group comprised of 236 studies with 4,086 patients in total, among which 236 deaths were reported. In the era prior to antibiotic treatment, the percentage of death was considerably higher in the first group compared to the second group. Though considering the selection bias of noma cases reported in the publications, it is very difficult and as described in the discussion, cautioned not to extrapolate the current fatality rate of noma from these results.

Table 13 presents the number of deaths and patients in studies reporting mortality, by noma stage. Zero deaths were reported in the 3 publications with patients in the scarring stage of noma and the 58 studies with participants in the sequelae noma stage. There were 17 deaths (850 patients) reported in 30 studies describing cases of acute necrotizing gingivitis. The highest rates of reported deaths were represented in patients described as being within the gangrenous phase (25/79; 31.6% patients) and oedema phase (8/29; 27.6% patients).

Table 18	Number of deaths recorded in included articles (n=283 unique studies with explicit reporting of deaths)

| **Study design** | **Time period** | **No. of studies** | **No. of patients** | **No. of deaths** |
| --- | --- | --- | --- | --- |
| Case Report/Series  (overall: 237 studies, 717 patients, 127 deaths) | Pre-1900 | 32 | 70 | 37 |
|  | 1900-1924 | 10 | 20 | 8 |
|  | 1925-1949 | 23 | 52 | 23 |
|  | 1950-1974 | 22 | 81 | 11 |
|  | 1975-1999 | 42 | 152 | 28 |
|  | 2000-2009 | 38 | 100 | 12 |
|  | Post-2010 | 70 | 242 | 8 |
| Cohort  (overall: 31 studies; 1664 patients; 152 deaths) | Pre-1900 | - | - | - |
|  | 1900-1924 | - | - | - |
|  | 1925-1949 | 4 | 148 | 65 |
|  | 1950-1974 | 3 | 106 | 22 |
|  | 1975-1999 | 4 | 115 | 23 |
|  | 2000-2009 | 10 | 730 | 14 |
|  | Post-2010 | 10 | 565 | 28 |
| Cross-sectional  (Overall:7 studies; 1560 patients, 6 deaths) | Pre-1900 | - | - | - |
|  | 1900-1924 | - | - | - |
|  | 1925-1949 | - | - | - |
|  | 1950-1974 | - | - | - |
|  | 1975-1999 | 1 | 22 | 0 |
|  | 2000-2009 | 3 | 943 | 3 |
|  | Post-2010 | 3 | 595 | 3 |
| Interventional  (Overall: 6 studies, 101 patients, 0 deaths) | Pre-1900 | - | - | - |
|  | 1900-1924 | - | - | - |
|  | 1925-1949 | 1 | 13 | 0 |
|  | 1950-1974 | - | - | - |
|  | 1975-1999 | 1 | 23 | 0 |
|  | 2000-2009 | 2 | 22 | 0 |
|  | Post-2010 | 2 | 43 | 0 |
| Other  (overall 2 studies; 139 patients; 6 deaths) | Post-2010 | 2 | 139 | 6 |

Table 19	Number of deaths reported in studies before 1950 by age group of included participants

|  | Pre-1900 | | 1900-1924 | | 1925-1949 | |
| --- | --- | --- | --- | --- | --- | --- |
|  | No. of studies**^c^** | No. of  deaths/ patients | No. of studies**^c^** | No. of  deaths/patients | No. of studies**^c^** | No. of  deaths/patients |
| Infant (<1) | 1 | 0 / 1 | - | - | - | - |
| Child (1 - <10) | 18 | 27 / 44 | 6 | 6 / 15 | 11 | 10 / 28 |
| Children (0-19)^a^ | 3 | 0 / 5 | 1 | 0 / 2 | - | - |
| Adolsecent (10 - 19) | - | - | 1 | 0 / 1 | - | - |
| Adult (>19) | 4 | 4 / 4 | 1 | 1 / 1 | 8 | 9 / 10 |
| All age groups^b^ | 3 | 3 / 8 | - | - | 6 | 65 / 167 |
| Not Reported | 3 | 3 / 8 | 1 | 1 / 1 | 3 | 4 / 8 |

a Children (0-19): This group was added for studies that did not specify sub-category of child’s age.

b All age groups denotes any study for which included patients ranged across multiple age categories (i.e., from infants through to adults)

c Total number of patients included in the 283 publications which reported mortality (deaths observed or zero deaths explicitly reported)

Table 20	Number of deaths reported in studies after 1950 by age group of included participants

|  | 1950-1974 | | 1975-1999 | | 2000-2009 | | Post-2010 | |
| --- | --- | --- | --- | --- | --- | --- | --- | --- |
|  | No. of studies**^c^** | No. of  deaths/ patients | No. of studies**^c^** | No. of  deaths/ patients | No. of studies**^c^** | No. of  deaths/ patients | No. of studies**^c^** | No. of  deaths/ patients |
| Infant (<1) | - | - | 2 | 3/4 | 1 | 0/1 | 17 | 12/150 |
| Child (1 - <10) | 11 | 23/120 | 18 | 32/141 | 9 | 6/14 | 4 | 6/108 |
| Children (0-19)^a^ | 1 | 3/19 | 3 | 0/52 | 5 | 9/606 | 7 | 0/10 |
| Adolsecent (10 - 19) | - | - | 6 | 0/6 | 6 | 0/6 | 41 | 4/70 |
| Adult (>19) | 10 | 4/10 | 12 | 8/15 | 13 | 5/15 | 15 | 23/1144 |
| All age groups^b^ | 2 | 3/32 | 5 | 8/28 | 19 | 9/1153 | 3 | 0/102 |
| Not Reported | 1 | 0/6 | 2 | 0/12 | - | - | - | - |

a Children (0-19): This group was added for studies that did not specify sub-category of child’s age.

b All age groups denotes any study for which included patients ranged across multiple age categories (i.e., from infants through to adults)

c Total number of patients included in the 283 publications which reported mortality (deaths observed or zero deaths explicitly reported)

Table 21	Number of deaths reported by noma stage of included participants

| **Noma Stage** | **No. of studies** | **No. of patients** |  | **No. of deaths** |
| --- | --- | --- | --- | --- |
| 1. Acute necrotizing gingivitis | 30 | 850 |  | 17 |
| 2. Oedema Stage | 24 | 29 |  | 8 |
| 3. Gangrenous Stage | 51 | 79 |  | 25 |
| 4. Scarring Stage | 3 | 3 |  | 0 |
| 5. Sequelae Stage | 58 | 583 |  | 1 |
| Multiple Stages | 98 | 2209 |  | 225 |
| Unclear | 19 | 428 |  | 15 |

a Total number of patients included in the 283 publications which reported mortality (deaths observed or zero deaths explicitly reported)

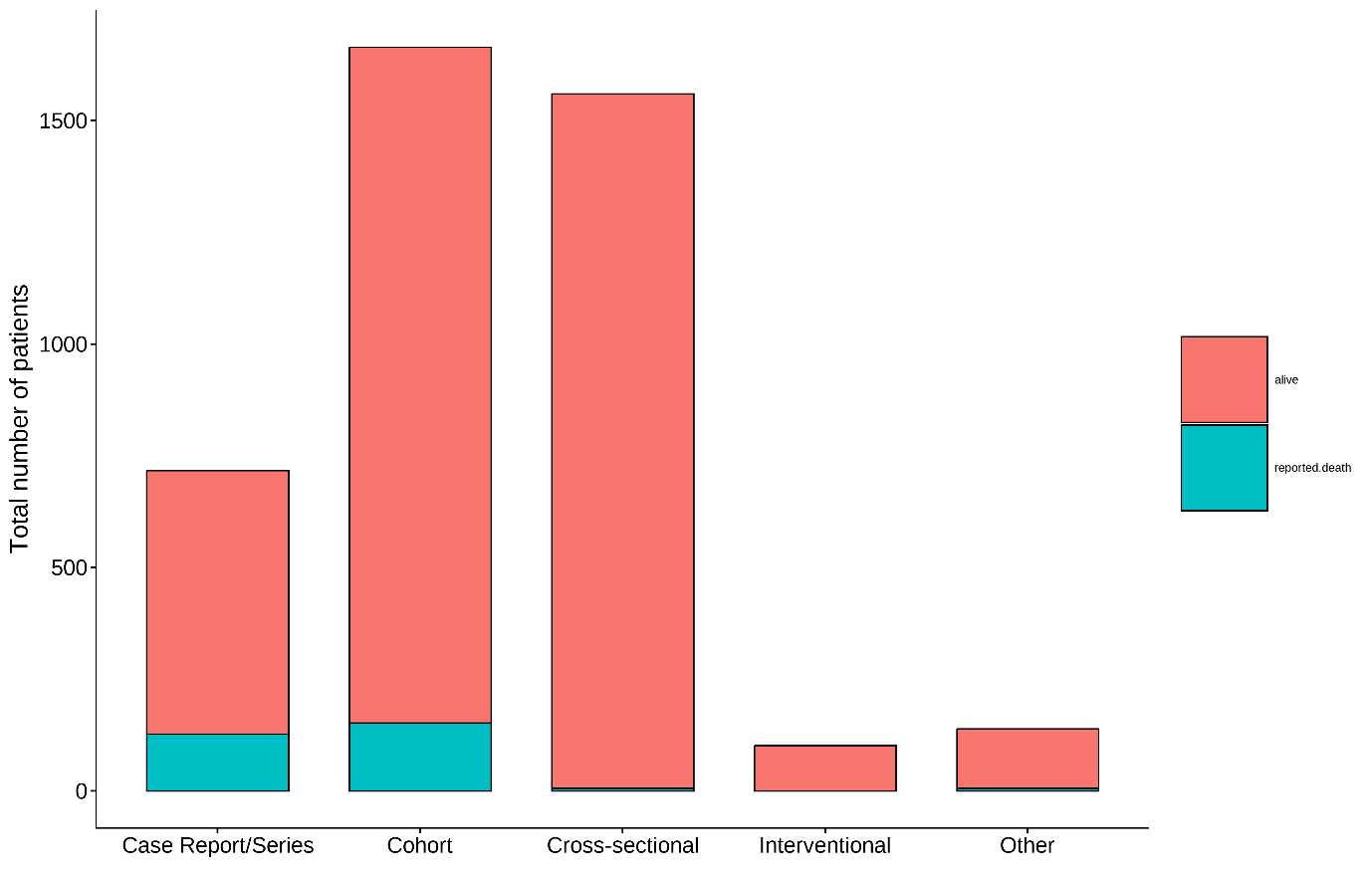

Figure 18	Reported number of deaths as a proportion of total patients by study design

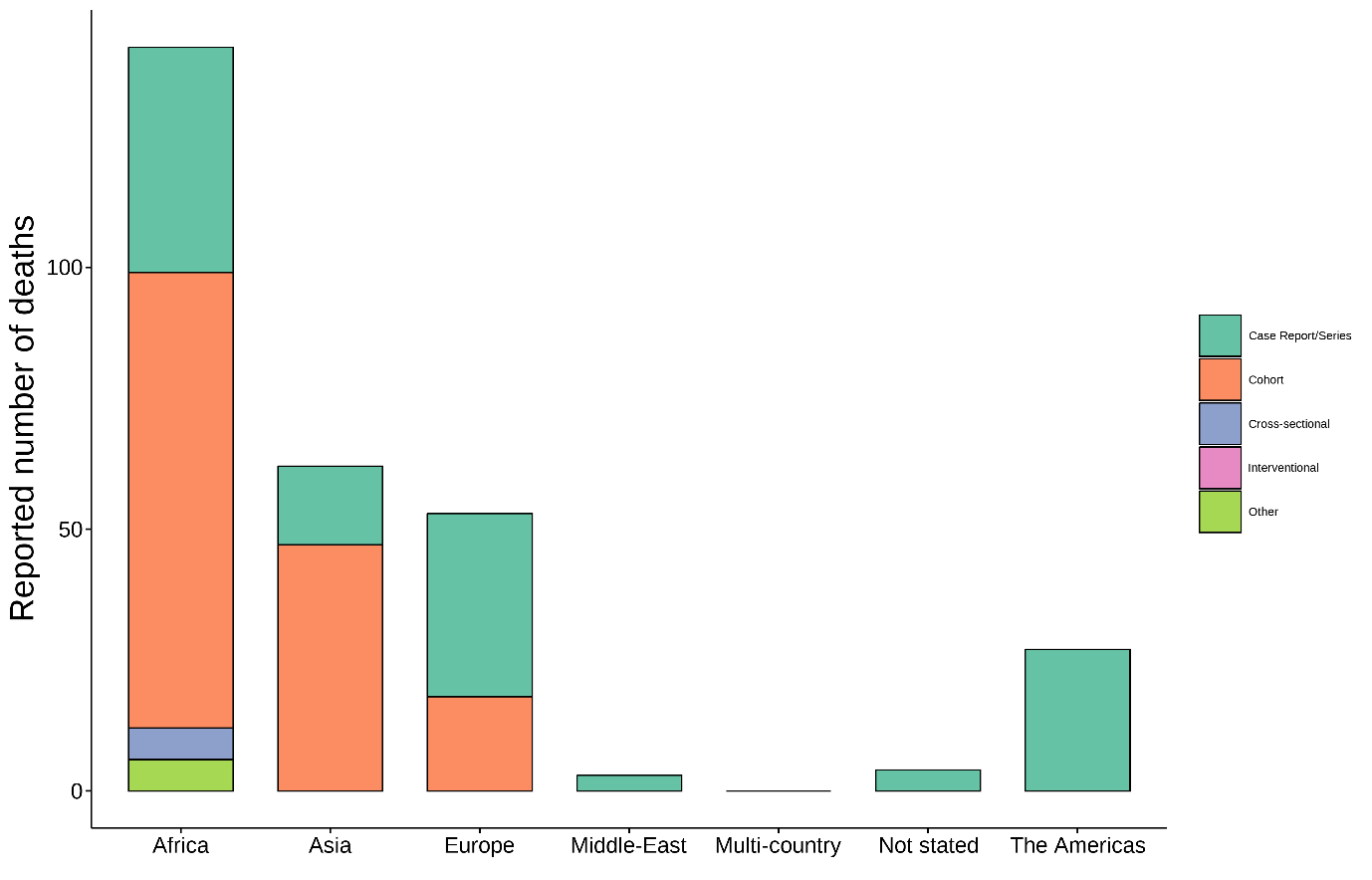

Figure 19	Reported number of patient deaths by region and study design

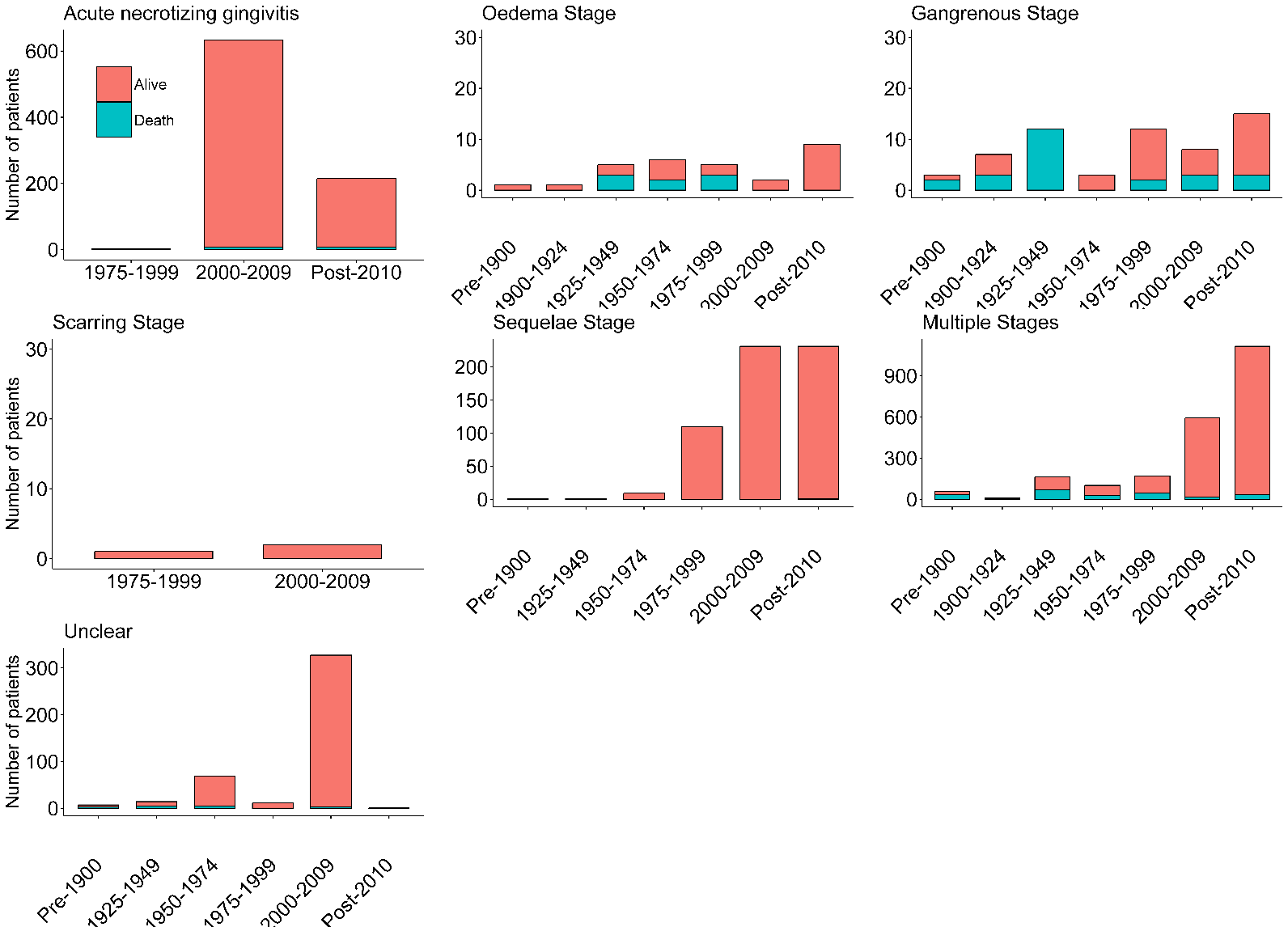

Figure 20	Number of patients by mortality status and disease stages across different time period (red: alive, death: blue)

Note: Different y-scales across the panels

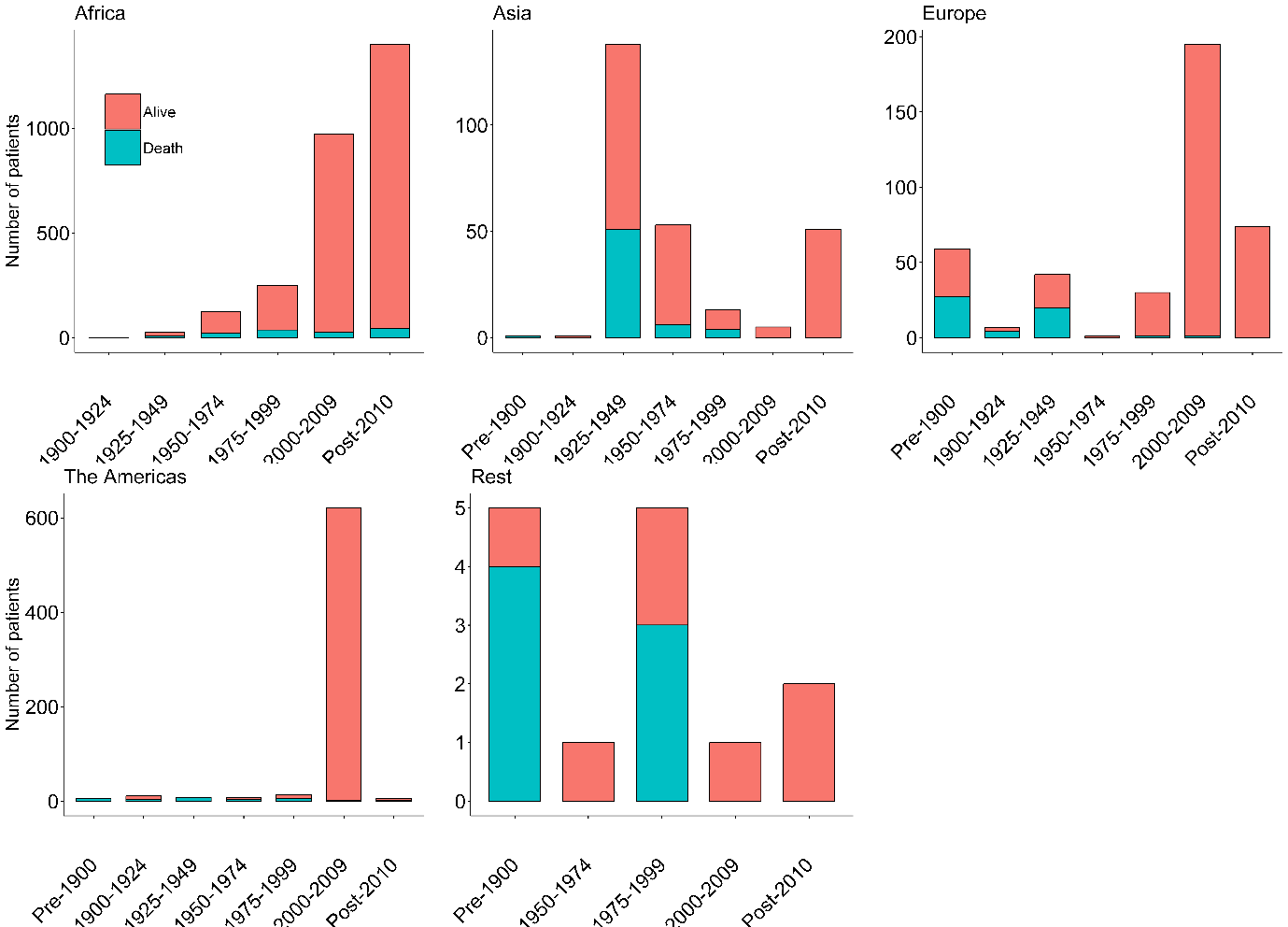

Figure 21	Number of patients reported deaths across regions by time-period

Rest includes Multi-country studies and studies with unclear country names and middle east.

##### a. Case-fatality of cohort and interventional studies

The observed case-fatality rate from the 37 cohort or interventional studies are presented in Figure 21. The median proportion of deaths reported from these 37 studies was 0% [inter-quartile range: 0-9.7%, range: 0-83.3%] (Table 22). However, this analysis is challenged as it includes those studies reporting on surgical intervention and includes noma survivors and those undergoing reconstruction. After excluding studies providing surgical intervention (for facial reconstruction or any other forms of surgery), there were 12 studies without surgical intervention. The median case-fatality rate reported from these 12 studies was 4.6% [IQR: 0-28.4%; range: 0-83.3%] (Figure 22). Using a formal meta-analysis of these 12 studies, the estimate of case-fatality was 9.9% [95% CI: 7.9%-12.2%; *I^2^*=87.3%] using a common-effect meta-analysis, and 5.5% [95% CI:0.9%-26.7%] from random-effects meta-analysis. Estimated 95% prediction interval was wide (0-94.2%) indicating large heterogeneity of the estimated effect. There was insufficient number of studies to undertake further sub-group meta-analysis. Summary estimates are presented in Table 23.

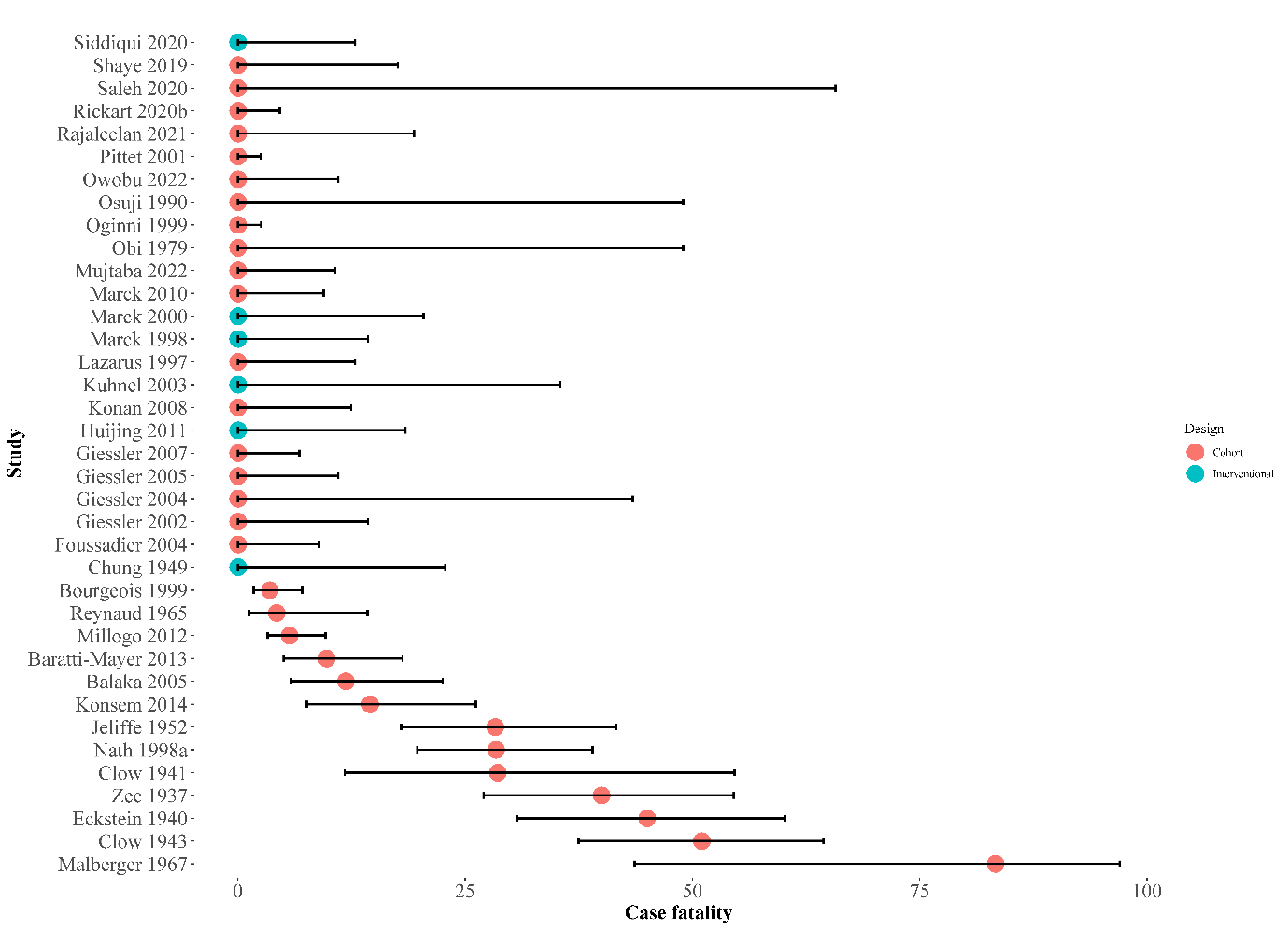

Figure 22	Forest plot depicting case-fatality rate in individual cohort and interventional studies

95% confidence interval for individual studies computed using Wilson’s method. The number of patients in each of the studies is shown in parenthesis.

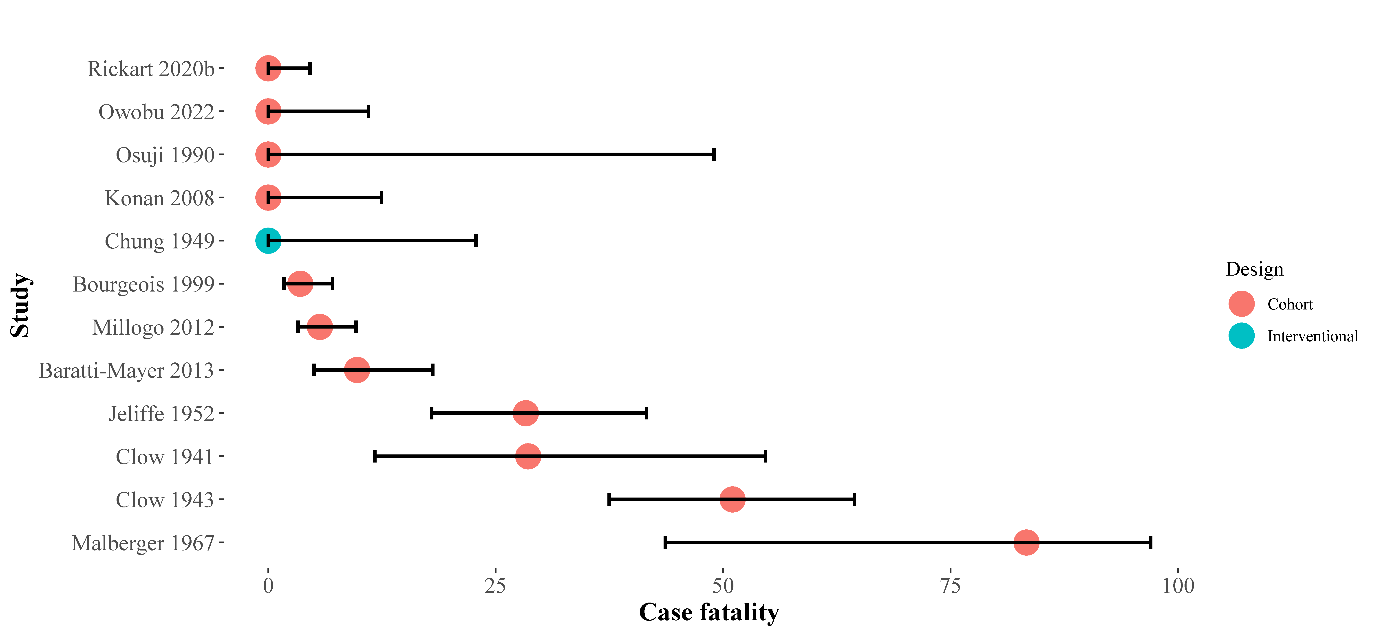

Figure 23	Forest plot depicting case-fatality rate in individual cohort and interventional studies that did not use surgical intervention (n=12 studies)

95% confidence interval for individual studies computed using Wilson’s method

Table 22	Case-fatality rate reported in prospective cohort and interventional studies (n=37 studies)

| **Time-period** | **Number of**  **Studies** | **Number of**  **Patients/deaths** | **Median reported case-fatality rate**  **[IQR; min-max]** |
| --- | --- | --- | --- |
| 1925-1949 | 5 | 161/65 | 40% [IQR =28.6%-45.0%; 0%-51%] |
| 1950-1974 | 3 | 106/22 | 28.3% [16.3%-55.8%;4.3%-83.3%] |
| 1975-1999 | 5 | 138/23 | 0% [0%-0%;0%-28.4%] |
| 2000-2009 | 12 | 752/14 | 0% [0%-0%;0%-11.9%] |
| Post-2010 | 12 | 608/28 | 0% [0%-1.4%;0%-14.9%] |

IQR = interquartile range

Table 23	Case-fatality rate reported in prospective cohort and interventional studies excluding studies of surgical interventions (n=12 studies*)

| **Time-period** | **Number of**  **Studies** | **Number of**  **Patients/deaths** | **Median reported case-fatality rate**  **[min-max]** |
| --- | --- | --- | --- |
| 1925-1949 | 3 | 29/76 | 28.6% (0%-51.0%) |
| 1950-1974 | 2 | 20/59 | 55.8% (28.3%-83.3%) |
| 1975-1999 | 1 | 0/4 | 0% |
| 2000-2009 | 2 | 7/226 | 1.8% (0%-3.5%) |
| Post-2010 | 4 | 20/405 | 2.8% (0%-9.8%) |

*Meta-analysis estimate of case-fatality: 9.9% [95% CI: 7.9%-12.2%] using a common-effect model; 5.5% [95% CI:0.9%-26.7%] using random-effects model; 95% prediction interval 0-94.2%

### 9. Post Noma Complications

Among the 366 included articles, 205 (56.0%) reported on post-noma complications. Post-noma complications related to the head region were observed in 142 articles. The most common complications in the head regions were trismus, sequestration of the jaws, bony ankylosis of temporomandibular joints, severe scarring, fistulisation of cheeks, and oro-nasal fistula. For the 6 publications which reported other or multiple (i.e., head and other) complications, the observed other complications post-noma included: mental defect, psychological, anaemia, albuminuria, loss of palmaris longus tendon, left pleural effusion, pulmonary emphysema, gangrenous pneumonia, “metastatic complications”, chronic pain, as well unclear or non-descript “major sequelae”. A total of 28 articles explicitly reported at the time of publication no complications occurred, while 133 studies did not report on complications. The number of studies reporting post-noma complications by noma stage are presented below.

Table 24	Number of studies reporting post noma complications

| **Post Noma Complications** | **No. of studies** |
| --- | --- |
| Reported any complication | 205 |
| Not Applicable – Death^a^ | 53 |
| Head Complications | 142 |
| Other Complications | 4 |
| Multiple Complications | 6 |
| No Complications^b^ | 28 |
| Not Reported | 133 |

a Reported post-noma complications however the patient subsequently died and was therefore not included for synthesis with post-noma complications in survivors.

b No complications related to those studies which had early treatment and no subsequent complications were reported.

Table 25	Number of studies reporting post noma complications by noma stage

| **Noma Stage** | **Head complications** | **Multiple complications** | **No complications** | **Not applicable -death** | **Other** | **Not reported** |
| --- | --- | --- | --- | --- | --- | --- |
| 1. Acute necrotizing gingivitis | 4 | 0 | 0 | 2 |  | 29 |
| 2. Oedema Stage | 9 | 1 | 7 | 5 |  | 5 |
| 3. Gangrenous Stage | 19 | 0 | 5 | 12 | 2 | 19 |
| 4. Scarring Stage | 1 | 0 | 1 | - |  | 1 |
| 5. Sequelae Stage | 57 | 1 | 6 | - |  | 12 |
| Multiple Stages | 44 | 4 | 7 | 31 | 2 | 39 |
| Unclear | 8 | 0 | 2 | 3 |  | 28 |

### 10. Burden of disease

Of the 366 studies included in this review, 117 (31.9%) reported indicators related to the burden of disease. Indicators defined as representative of the burden of disease for the purposes of this review included incidence, prevalence, quality-adjusted life years (QALYs), disability-adjusted life years (DALYs), financial costs, morbidity or mortality rates. Primary estimates (i.e., rates or costs calculated directly from the primary research reported in the publication) were captured in addition to burden of disease estimates from secondary sources (i.e., rates reported as sourced from other literature and not calculated from the primary data collected in the publication).

Of the 118 publications that reported burden of disease indicators, only 17 reported primary source estimates. Secondary sources were cited to report the burden of disease in 91 studies and 10 cited both primary and secondary source estimates. Further details are provided in Table 26.

Table 26	Number of studies reporting burden of disease

|  | **Number of studies** |
| --- | --- |
| **Primary reports** | **17** |
| Incidence | 8 |
| Prevalence | 6 |
| Both incidence and prevalence | 2 |
| Other | 1 |
| **Secondary reports** | **91** |
| **Both Primary and secondary reports** | **10** |
| **Total** | **118** |

##### a. Before 1950

From the included literature prior to 1950, no estimates of the worldwide or even national incidence or prevalence estimates of noma were published. Table 27 below presents the average yearly incidence of noma at individual sites as obtained from hospital records from before 1950.

Studies included in this review report 62 noma patients in Europe from 1839 until 1950. The true burden is expected to have been considerably higher. Secondary sources within the included papers report a further 779 patients in Europe over the same period.

In China, between 1936 and 1950, included studies report 179 cases of noma, often associated with visceral leishmaniasis. An additional 100 patients admitted to a Chinese hospital with noma from 1921-1935 were reported from a secondary source (Mehrotra, 1966).

There were few cases of noma from Africa and other regions of Asia. This is almost certainly due to the absence of published records, rather than the absence of the disease itself. The total number of cases reported in the published literature before 1950 by country is presented in Table 2.

Table 27	Prior to 1950: Incidence of noma obtained from retrospective hospital records

| **Study** | **Primary Source of estimate^a^** | **Country** | **Years** | **Number of patients** | **Average annual incidence at study site^b^** |
| --- | --- | --- | --- | --- | --- |
| Mehrotra 1966 | Fu-tang 1936 | China | 1921-1935 | 100 | 7.1 |
| Fan 1934 | Yes | China | 1927-1933 | 57 | 11.4 |
| Zee 1937 | Yes | China | 1933-1936 | 45 | 15 |
| Fleming 1893 | Yes | England | 1876-1888 | 6 | 0.5 |
| Fleming 1893 | Yes | England | 1881-1887 | 5 | 0.8 |
| Fleming 1893 | Ranke (citation unclear) | Germany | Unknown (21 years) | 2 | 0.1 |
| Ritchie 1872 | Yes | Scotland | 1863-1871 | 8 | 0.7 |
| Kittredge 1872 | Yes | USA | 1863-1864 | 67 | 67 |

a If the study in question provides an estimate based on a calculation from primary data collected in that study “Yes” is entered. Where a secondary source is reported, the citation of the primary source (First author, date of publication) is provided

b Calculated for the purposes of this review by dividing the total number of incident cases by the total number of years of observation at a given study site.

##### b. Between 1950 and 2000

From 1950 onwards, noma began to appear more frequently in reports from the region of Sub-Saharan Africa now described as the noma belt, whilst almost disappearing from Europe and the USA.

Reported estimates of the incidence of noma in countries within the Sub-Saharan region differ. The WHO estimated that, in 1997, the annual incidence for Sub-Saharan Africa was 2-10 cases per 10,000 children.

In the 1997 publication by Barmes *et al*, a higher national case incidence was estimated in Senegal of 7-12 cases per 10,000 in children aged 0-6. Two years later, in Senegal, the incidence was estimated by Bourgeois *et al* 1999 to range from 2.8-8.4/10,000 cases nationally in children between the ages of 1-4. This estimate was based on the 1981-1993 clinical records of the National Hospital Le Dantec in Dakar, the tertiary reference hospital for all maxillofacial diseases until 1994.

In Niger, annual incidence was reported between 7-14 cases per 10,000 children between the ages of 0-6.

The included literature also highlighted discrepancies of estimated incidence rates within a country versus reported cases observed within that country. As an example, a reported national case incidence of 1:1,250 in Nigeria (Barmes 1997) differed from presentation of cases at individual study sites, with an annual incidence ranging from 0.3 cases (Oji 2002; 3 cases in 10 years) to 3.3 cases (Fieger 2003; 46 cases over 14 years).

Worldwide, the annual incidence of noma was estimated to be 500,000 cases by the WHO in 1994 (Costini 1995, as cited in Marck 1998). Four years later, in 1998, the WHO published their latest estimate of annual incidence as 140,000 acute noma patients worldwide. The 1998 report also estimated the global prevalence of noma survivors to be 770,000.

The total number of cases reported in the literature from 1950 to 2000 by country is presented in Table 3.

Table 28	From 1950 to 2000: Incidence of noma obtained from retrospective hospital records

| **Study** | **Primary source of estimate^a^** | **Country** | **Years** | **Number of patients** | **Average annual incidence at study site^b^** |
| --- | --- | --- | --- | --- | --- |
| Balaka 2006 | Yes | Burkina Faso | 1986-1996 | 70 | 7 |
| Tall 2006^c^ | Yes | Burkina Faso | 1987-1996 | 59 | 5.9 |
| Balaka 2006 | N'Gouoni 1995 | Congo | 1987-1996 | 11 | 1.6 |
| Malberger 1967 | Yes | Gambia | 1964-1965 | 6 | 6 |
| Mehrotra 1966 | Yes | India | 1956-1963 | 20 | 2.9 |
| Balaka 2006 | Gadegbeku 1994 | Ivory Coast | Unknown (20 years) | 49 | 2.5 |
| Diombana 1998 | Yes | Mali | 1981-1993 | 22 | 1.8 |
| Sheiham 1966 | Emslie 1963 | Nigeria | Unknown (3 months) | 50 | 200 |
| Oji 2002 | Tempest 1966 | Nigeria | 1962-1965 | 250 | 83.3 |
| Adekeye 1983 | Yes | Nigeria | 1978-1982 | 140 | 35 |
| Oji 2002 | Sawyer 1981 | Nigeria | Unknown (1 year) | 26 | 26 |
| Ojinni 1999 | Yes | Nigeria | 1982-1996 | 146 | 10.4 |
| Obiechina 2000 | Yes | Nigeria | 1982-1998 | 173 | 10.8 |
| Osuji 1990 | Yes | Nigeria | 1983-1984 | 5 | 5 |
| Oji 2002 | Yes | Nigeria | 1990-1999 | 3 | 0.3 |
| Diop 1972 | Yes | Senegal | 1959-1976 | 277 | 16.3 |
| Diombana 1998 | Lozes 1973 | Senegal | 1960-1982 | 155 | 7 |
| Bourgeois 1999 | Yes | Senegal | 1981-1993 | 199 | 16.6 |
| Lazarus 1997 | Yes | South Africa | 1960-1995 | 26 | 0.7 |
| Nath 1998 | Yes | Zambia | 1979-1993 | 117 | 7.8 |

a If the study in question provides an estimate based on a calculation from primary data collected in that study “Yes” is documented. Where a secondary source is reported, the citation of the primary source (First author, date of publication) is provided.

b Calculated for the purposes of this review by dividing the total number of incident cases by the total number of years of observation at a given study site.

c Tall et al 2006 is a subset of the population assessed in the Balaka et al., 2006 study

Table 29	From 1950 to 2000: Incidence & prevalence estimates of noma obtained from all other sources

| **Study** | **Primary source of estimate^a^** | **Year** | **Country / Region** | **Estimate** | **Notes** |
| --- | --- | --- | --- | --- | --- |
| WHO 1994 | WHO 1994 | 1994 | Some regions of Africa | 2-7 cases per 10,000 children up to the age of 6 | Regions unspecified |
| Mamadou 2002 | WHO 1997 | 1997 | Sub-Saharan Africa | 2-10 cases per 10,000 |  |
| Bah 2009 | WHO 1998 | 1998 | Some regions of Africa | 2-4 cases per 10,000 children aged 2-6 | Regions unspecified |
| Whiteson 2014 | Bourgeois and Leclercq 1999 (WHO) | 1999 | Sub-Saharan Africa | 100,000 new cases every year in children aged 1-7 |  |
| Fieger 2003 | Barmes 1997 | 1997 | Niger | 1.34 cases per 1,000 children aged 0-6 |  |
| WHO 1998 | Yes | 1991-1992 | Niger | 7-14 cases per 10,000 children up to the age of six | Estimates obtained from a national study of cases |
| Fieger 2003 | Barmes 1997 | 1997 | Nigeria | 1:1250 cases | The estimate was built on data from 3 hospitals in Nigeria |
| Idigbe 1999 | Yes | 1999 | South-west Nigeria | 10 cases of noma were identified | Survey undertaken to examine risk factors |
| Idigbe 1999 | Yes | 1999 | North-west Nigeria | 129 cases of noma were identified | Survey undertaken to examine risk factors |
| Bourgeois 1999 | Yes | 1999 | Senegal | 2.8-8.4 cases per 10,00 children aged 1-4 |  |
| Marck 1998 | WHO 1994 | 1994 | Worldwide | 500,000 new cases every year |  |
| Enwonwu 2005 | WHO 1998 | 1998 | Worldwide | Prevalence of 770,000 noma survivors (30,000 with severe limitations) |  |
| Chiandussi 2009 | WHO 1998 | 1998 | Worldwide | 140,000 new cases every year |  |

a If the study in question provides an estimate based on a calculation from primary data collected in that study “Yes” is documented. Where a secondary source is reported, the citation of the primary source (First author, date of publication) is provided

##### c. After 2000

Since 2000, studies citing a secondary source of the burden of noma most commonly used the WHO’s 1998 estimate of global incidence which estimates an incidence of 140,000 new cases a year (Shaye 2019, Madsen 2017, Tungotyo 2017, Lembo 2011, O'Brien 2019, Park 2022, Rajaleelan 2021, Rickart 2020b, Ver-Or 2022, Bouassalo 2019, Hartmann 2006).

The latest global estimate was published in 2003. Fieger *et al.,* reported a lower global incidence of 30,000-40,000 cases per year. Of these, 25,600 cases were expected to originate from Sub-Saharan Africa. It’s important to note this figure was calculated using the numbers of noma patients in north-west Nigeria which were used to calculate national incidence before being extrapolated to a global incidence. This global estimate from Fieger *et al*., was referenced in 6 recent publications. Global prevalence of noma survivors was estimated to be 210,000 noma survivors worldwide, based on the 15% survival rate and a life expectancy of 40 years following the disease (Srour 2017, Ruegg 2018).

More recent incidence and prevalence estimates calculated using primary data have been localised to specific geographic regions or territories within the countries of Nigeria, Burkina Faso and one study in Chile (Table 30 and Table 31). Estimates vary greatly within and across countries. For example in Nigeria, in the northeast a prevalence of 6.4 per 1,000 population is reported by Fieger et al., a prevalence of 7 per 1,000 from a hospital-based study in Ibadan by Denloye et al., Bello et al., reported a prevalence rate of 8.3 to 100,000 (0.083 per 1,000 population) in the north–central zone of the country, whereas Farley et al., 2020a documented a substantially higher prevalence of 3.3% in two north-western Nigerian states. The substantially higher prevalence is on account of 181 (93.3%) of these 194 cases being classified as Stage 0, presenting with simple gingivitis.

Included literature also suggests the number of HIV positive noma patients are increasing. A study conducted in 2008 by Chidzonga et al., explored HIV-positive patients who presented with noma in Zimbabwe between 2002 and 2006. They saw 3, 7, 9, 13, and 16 cases in the years 2002, 2003, 2004, 2005, and 2006 respectively. This study concluded female children (average age: 9.2 years; range: 1 to 36 years) appear to be more commonly affected than their male counterparts (average age: 14.2 years; range, 3 to 30 years).

The total number of cases reported in the literature from 2000 to December 2022 by country is presented in Table 4.

Table 30	After 2000: Incidence of noma obtained from retrospective hospital records.

| **Study** | **Country** | **Years** | **Number of patients** | **Average annual incidence at study site^a^** |
| --- | --- | --- | --- | --- |
| Bonkoungou 2006 | Burkina Faso | 1986 - 1996 | 70 | 7 |
| Millogo 2012 | Burkina Faso | 1998-2007 | 212 | 23.5 |
| Konsem 2014 | Burkina Faso | 2003-2012 | 55 | 5.5 |
| Adeola 2004 | Nigeria | 1991-2001 | 252 | 25.2 |
| Adeniyi 2019 | Nigeria | 1999-2011 | 159 | 12.2 |
| Bello 2019 | Nigeria | 2010-2018 | 78 | 8.3 |

a Calculated for the purposes of this review by dividing the total number of incident cases by the total number of years of observation at a given study site.

Table 31	After 2000: Prevalence of noma obtained from prospective and retrospective surveys

| **Study** | **Country** | **Years** | **Prospective/ retrospective** | **Sample size** | **Number of patients with noma** | **Notes and reported prevalence** |
| --- | --- | --- | --- | --- | --- | --- |
| Lopez 2002 | Chile | 1999 | Prospective | 9,203 | 618^a^ | Point prevalence of 6.7% (95% CI = [6.2; 7.3]) |
| Farley 2020a | Nigeria | 2018 | Prospective | 7,122 | 194 | The prevalence of any stage of noma was identified in 3.3% of sampled children (0-15years). |
| Bello 2019 | Nigeria | 2010-2018 | Retrospective | Not reported^b^ | 78 | Period prevalence of noma which incorporated all cases seen within the study period was 1.6 per 100,000 |

a Students who showed signs of NUG

b Period prevalence calculated and reported in the publication. Denominator or total number of all records reviewed from all surgical outreach programmes organized in north central Nigeria was not explicitly reported in the publication

Table 32	After 2000: Incidence & prevalence estimates of noma as reported in the literature from all other sources

| **Study** | **Primary source of estimate** | **Year** | **Country / Region** | **Estimate** | **Notes** |
| --- | --- | --- | --- | --- | --- |
| Fieger 2003 | Fieger 2003 | 2003 | Noma belt | 24,600 new cases every year. | Numbers of noma patients in Nigeria were used to calculate national incidence before extrapolated to incidence in the noma belt. |
| Cocquempot 2014 | WHO 2006 | 2006 | Africa region | 42,800 new cases |  |
| Fieger 2003 | Fieger 2003 | 2003 | North-west Nigeria | 6.4 cases per 1000 children (<15 years age) | Numbers of noma patients in Nigeria were used to calculate national incidence before extrapolated to incidence in the noma belt. |
| Enwonwu 2006 | Enwonwu 2006 | 2006 | Gambia | 1.9 cases per 1,000 children (age range unspecified) | Calculated using hospital records obtained from Bickler 2000 |
| Fieger 2003 | Barmes 1997 | 1997 | Senegal | 0.7-1.2 cases per 1000 children aged 0-6 | Estimates obtained from a national study of cases. |
| Fieger 2003 | Fieger 2003 | 2003 | Worldwide | 30-400 new cases every year | Numbers of noma patients in Nigeria were used to calculate national incidence before extrapolated to global incidence |
| Srour 2017 | Srour 2017 | 2017 | Worldwide | 210,000 noma survivors | Accounts for survival rate and life expectancy |
| Bello 2019 | Bello 2019 | 2019 | Nigeria | Incidence of 8.3 per 100,000 in north central  Nigeria.  Period prevalence of 1.6 per 100,000 population at risk. | With a range of 4.1–17.9 per 100,000 observed across different states within the geopolitical zone.  Which incorporated all cases seen within the 9-year study period was |
| Bello 2019 | Fieger 2003 | 2017  2003 | Worldwide  Nigeria | 30,000–40,000 cases  Incidence rates between 0.8–6.4 per 1,000 children | With 75% occurring in sub-Saharan Africa  Reported in the last two decades mostly according to data provided by foreign non-governmental organizations or surgical missions. |
| Braun 2020 | Srour 2017 | 2017 | Worldwide | “The global incidence per year may be approximately 30,000-40,000 cases” | Paper simply references Srour 2017. Likely Fieger 2003 estimate. |
| Ding 2020 | Srour 2017 | 2017 | Worldwide | “There are an estimated  30,000–40,000 cases per year worldwide.” | Paper simply references Srour 2017. Likely Fieger 2003 estimate. |
| Farley 2020a | Farley 2020 | 2020 | Nigeria | The prevalence of any stage of noma 3.3% of sampled children. | Prospective survey children 0-15 years |
|  | Bello 2019; Fieger 2003 | 2003  2019 | Nigeria | 7 cases per 1,000 children aged between 1 and 16 years (2003) to 6.4 per 1,000 children (2003) to 1.6 per 100,000 population at risk (2010–2018). | Northwest Nigeria and North central Nigeria |
|  | WHO 1998 | 1998 | Worldwide | The WHO estimates 770,000 people are currently living with noma globally. |  |
| Gebretsadik 2022a | Bello 2019 | 2019 | Nigeria | Incidence of 8.3 per 100,000 in north central  Nigeria.  Period prevalence of 1.6 per 100,000 population at risk. | With a range of 4.1–17.9 per 100,000 observed across different states within the geopolitical zone.  Which incorporated all cases seen within the 9-year study period was |
| Isah 2021 | Farley 2020 | 2018 | Northwest Nigeria | 3,300 out of every 100,000 children in the 0–15-year age | In northwest Nigeria in 2018, estimated to have any stage of noma at the time of the survey. |
| O'Brien 2019 | WHO | - | Worldwide | Incidence of  140,000 annually, and the prevalence in northern Nigeria is estimated at 6.4/1000 children. | Abstract: no source provided |
| Park 2022 | - | - | Worldwide | The incidence and prevalence of noma is reported as 140,000 and 770,000 cases, respectively. | Primary source unclear. Paper later cites Auluck 2005 and Oji 2002. |
| Putri 2021 | - | - | Africa | The condition is predominantly suffered in Africa, with an estimation of 20 instances per 100,000 individuals. | No citation or primary source provided. |
| Rajaleelan 2021 | - | - | Worldwide | Affecting up to 1,40,000 children annually. | Abstract: no citation or primary source provided. |
| Rickart 2020b | [WHO](https://apps.who.int/iris/handle/10665/254579) | 2017 | Worldwide | 140,000 new cases a year |  |
| Ver-Or 2022 | Srour 2017; Fieger 2003; WHO 1998 | 1998-2017 | Worldwide | Annual global incidence 30,000 to 140,000 | Range and multiple secondary sources |
|  | Oji | 2002 | Nigeria | 3 patients in 10 years | Enugu |
|  | Fieger 2003 | 2003 | Nigeria | prevalence of 6.4 per 1,000 population | Northeast Nigeria |
|  | Denloye | 2003 | Nigeria | prevalence of 7 per 1,000 | hospital-based study in Ibadan |
|  | Bello 2019 | 2018 | Nigeria | Prevalence rate of 8.3 to 100,000 (0.083 per 1,000 population) | from north–central zone of Nigeria |
|  | Farley 2020 | 2020 | Nigeria | 3.3% prevalence rate | Northeastern Nigeria |
| Bouassalo 2019 | - | - | Worldwide | Incidence of 140,000 cases and prevalence at 770,000 cases. | Publication cites secondary sources Ashok 2015, Enwonwu 2005 and Tonna 2010 |
| Hartmann 2006 | Fieger 2003 | 2003 | Worldwide | Annual incidence of 25,600–140,000 cases |  |

Only a single calculation of the disability adjusted life-years (DALYs) of noma was encountered in this systematic review. In 2013 a preliminary study, only identified as a conference abstract (Haesen 2013) but never subsequently published, estimated the global burden of noma to range from 1-10 million DALYs, once premature mortality and post-noma disability was accounted for. This preliminary estimate was reported not to include the burden of the disease to the household or take into consideration the impact of the stigma or psychological effects of noma. No subsequent study with details of the calculations nor an update to this “preliminary” DALY estimate was identified in this literature review.

| **Publication** | **Year** | **Country/ Region** | **Estimate** | **Notes** |
| --- | --- | --- | --- | --- |
| Haesen et al. | 2013 | Worldwide | 1-10 million DALYs | Accounts for premature mortality and disability for surviving noma patients.  Does not account for impacts to household, stigma, psychological effects |

This DALY estimate for noma was also referenced as a secondary source in Srour et al., 2017.

Further analyses of the estimates should be conducted to assess the strength of the evidence but was beyond the scope of this work.

### 11. Risk of Bias

Given the heterogeneity of study design in included studies, The Oxford Centre for Evidence-Based Medicine Levels of Evidence was used to assess the level of evidence and quality of individual studies across this systematic review (Table 33). As specified by the objectives of this systematic review, all study types were included in the planned narrative synthesis.

Evidence assessed as Level 1 represent systematic reviews of randomised controlled trials or systematic reviews of high-quality observational studies with dramatic effect or nested case-control studies. Studies assigned with Level 2 bias represent individual randomised trials or observational studies with dramatic effect. For Level 3 this represents non-randomized controlled cohort or follow-up studies. Level 4 captures case-series, case control studies, or poor-quality prognostic cohorts or historically controlled studies. Level 4 evidence represents most of the literature published on noma. Level 5 evidence is mechanism-based reasoning.

For the level of evidence assigned to each of the included studies refer to the ‘Risk of bias’ file in Supplementary File 2.

Table 33	OCEBM levels of evidence for included noma literature

|  | **Number of studies** |
| --- | --- |
| Level 1 | 0 |
| Level 2 | 1 (0.3%) |
| Level 3 | 24 (6.6%) |
| Level 4 | 341 (93.2%) |
| Level 5 | 0 |

ROB SOURCE: Centre for Evidence-Based Medicine (CEBM), Nuffield Department of Primary Care Health Sciences. Oxford Centre for Evidence-based Medicine – Levels of Evidence (March 2009). Website accessed: 08 January 2020 <https://www.cebm.ox.ac.uk/resources/levels-of-evidence/ocebm-levels-of-evidence>

**Reporting Biases**

Table 34 presents the number of studies and patients for each synthesis assessed for which there was missing results. These reporting biases highlight the recommendation of this systematic review to develop minimum reporting guidance for future noma studies.

Table 34	Missing results across areas of synthesis in the included publications (reporting biases)

| **Area of Synthesis** | **Number of Studies** | **Number of patients** |
| --- | --- | --- |
| Patient age was not reported | 21 (5.7%) | 523 |
| Microbiology was not observed/reported | 253 (69.1%) | 13,998 |
| Risk factors were not observed/reported | 97 (26.5%) | 1,837 |
| Treatment details were not reported | 14 (3.8%) | 8,210 |
| Burden of disease indicators were not observed/reported | 248 (67.8%) | 2,931 |

**Certainty in the body of evidence**

There was highly variability in both the study designs and heterogeneity of methods across the included literature. Additionally, this review does not address specific clinical questions or meta-analyse treatment outcomes. Accordingly the utility and accuracy of any assessment of the strength of evidence should be cautioned. Nonetheless, based on the OCEBM levels of evidence assigned to each study, we have visually categorised the certainty of evidence by comparing the randomised and non-randomised studies included in this review. For the 6 randomised studies, the average OECBM level of evidence was 3 (Moderate), and for the 360 non-randomised studies, the average OECBM level of evidence was 4 (low). Therefore, the totality of evidence is mapped in Table 35 to ‘N’ (with ‘A’ representing the highest certainty and ‘Y’ the lowest certainty of evidence).

This categorisation is presented only to further highlight the results of this review that there are very few high-quality studies which assess the aetiology and treatment modalities for noma. The majority of included studies (n=270; 74%) were case studies, which are automatically regarded as low-quality evidence, heavily weighting any assessment of the certainty in the body of evidence represented in this review. However, given the paucity of information and knowledge gaps related to noma, the inclusion of case studies in the systematic review protocol was intentional. This review aimed to exhaustively synthesise all available evidence. While limited in their generalisability and ability to provide conclusive or applicable evidence, case studies can provide valuable insights and contextual understanding other study designs cannot. Broadening the scope of this review to include all study designs has resulted in a rich and exhaustive list of all potential risk factors, microbiology and treatments documented in the published literature. This work offers a comprehensive baseline knowledge from which to innovately approach future noma research and the design of new high-quality robust studies that are desperately needed to address the many knowledge gaps of noma.

Table 35	Representation of the certainty of evidence for randomised and non-randomised studies included in the evidence synthesis

|  | | Non-Randomised Studies | | | | |
| --- | --- | --- | --- | --- | --- | --- |
| Randomised studies |  | *Very High*  *1* | *High*  *2* | *Moderate*  *3* | *Low*  *4* | *Very Low*  *5* |
|  | *Very High*  *1* | A | B | C | D | E |
|  | *High*  *2* | F | G | H | I | J |
|  | *Moderate*  *3* | K | L | M | N | O |
|  | *Low*  *4* | P | Q | R | S | T |
|  | *Very Low*  *5* | U | V | W | X | Y |

Based on average OECBM level of evidence by study design (Table 33 & Sup File 2_Risk of bias)

Table adapted for the purposes of this study from C.A. Cuello-Garcia et al./Journal of Clinical Epidemiology 142 (2022) 200-208.
